# Single cell multiomics and 3D genome architecture reveal novel pathways of human heart failure

**DOI:** 10.1101/2025.05.08.25327176

**Authors:** Yang Xie, Luca Tucciarone, Elie N. Farah, Lei Chang, Qian Yang, Thirupura S. Shankar, Weston Elison, Shaina Tran, Jovina Djulamsah, Audrey Lie, Timothy Loe, Alyssa R. Holman, Sierra Corban, Justin Buchanan, Sainath Mamde, Haowen Zhou, Ruth M. Elgamal, Eleni Tseliou, Vincent Huang, Zhaoning Wang, Jeffery Chiu, Rebecca Melton, Emily Griffin, Qingquan Zhang, Jacinta Lucero, Sutip Navankasattusas, Daofeng Li, Chanrung Seng, Eugin Destici, Craig H. Selzman, Agnieszka D’Antonio-Chronowska, Ting Wang, Allen Wang, Stavros G. Drakos, Kyle J. Gaulton, Bing Ren, Neil C. Chi

## Abstract

Heart failure is a leading cause of morbidity and mortality; yet gene regulatory mechanisms driving cell type-specific pathologic responses remain undefined. Here, we present the cell type-resolved transcriptomes, chromatin accessibility, histone modifications and chromatin organization of 36 non-failing and failing human hearts profiled from 776,479 cells spanning all cardiac chambers. Integrative analyses revealed dynamic changes in cell type composition, gene regulatory programs and chromatin organization, which expanded the annotation of cardiac *cis*-regulatory sequences by ten-fold and mapped cell type-specific enhancer-gene interactions. Cardiomyocytes and fibroblasts particularly exhibited complex disease-associated cellular states, gene regulatory programs and global chromatin reorganization. Mapping genetic association data onto cell type-specific regulatory programs revealed likely causal genetic contributors to heart failure. Together, these findings provide comprehensive, multimodal gene regulatory maps of the human heart in health and disease, offering a valuable framework for designing precise cell type-targeted therapies for treating heart failure.

## Main Text

Heart failure (HF) remains a global epidemic and one of the major causes of morbidity and mortality (*1*). The etiologies of HF are multifactorial, and can be due to either acquired cardiovascular diseases such as ischemia, diabetes, hypertension, infections, or toxins, as well as genetic causes including monogenic dilated, hypertrophic and restrictive cardiomyopathies (*2*–*4*). Moreover, recent cardiac genome wide association studies (GWAS) have uncovered a substantial number of new HF-associated risk loci, revealing the importance of polygenic background on HF risk and cardiac ventricular traits (*5–8*). While fine-mapping of these loci identified genetic variants within coding regions, the majority of these likely causal genetic variants reside within non-coding regions (>85%) and remain to be functionally annotated (*5–8*). Delineating how HF-associated risk loci within non-coding regions may impact the function of specific cardiac cell types critical for maintaining cardiac performance will provide new opportunities for developing novel therapeutic or disease prevention strategies.

The heart consists of diverse cardiac cell types which coordinately act to sustain cardiac function and circulation (*9*). However, how these cardiac cell types maintain heart performance and how they are disrupted in ischemic and non-ischemic HF remain to be fully defined. While single-cell transcriptomic studies have begun to elucidate specific cell types and cellular interactions in non-failing and failing human hearts (*10*), the gene regulatory sequences and networks that direct their pathologic HF outcomes remain unresolved. Furthermore, since non-coding risk variants of human diseases are likely to act through disruption of transcription factor binding and dysregulation of gene expression, comprehensively investigating gene regulatory programs that direct cardiac cell type-specific responses to HF are paramount to improving our knowledge of the pathogenesis of HF and the development of new strategies to modulate cellular function to prevent and/or treat cardiac disease. Interrogating the chromatin landscape including its epigenomic and structural regulation enables not only the illumination of gene regulatory networks that control cell type-specific function and responses to cardiac stress through multiple layers of genomic organization (*11*) but also annotation of HF-associated genetic variants within non-coding regions that may impact cell type-specific gene regulation. Recent advancements in single-cell technologies provide the opportunity to examine the epigenome and chromatin architecture across distinct cell types in human tissues to address these crucial questions (*12*, *13*).

Toward this end, we performed single-cell multiomic assays to investigate the transcription, chromatin accessibility, histone modification status, and 3D chromatin conformation of individual cells from all cardiac chambers of 36 adult non-failing and failing (including ischemic and non-ischemic) human hearts. Integrative analyses of these multimodal data defined key cell type-specific gene regulatory and transcriptional programs participating during HF, and in combination with genetic association data, identified novel cell type-specific gene regulatory programs enriched with disease associated genetic variants that drive HF and overall cardiovascular risk.

## Results

### Cell type-specific transcriptomic, epigenomic and 3D genome maps of human hearts

To investigate the cell type-specific transcriptomic and epigenomic landscapes of non-failing (non-HF, control) and failing (HF) hearts, 36 donor hearts, which included both ventricular and atrial cardiac chambers, were collected from 23 HF and 13 non-HF patients, and used for our single-cell multimodal studies (**Fig. 1A, B, table S1**). The HF group comprised 13 ischemic cardiomyopathy (ICM) and 10 non-ischemic cardiomyopathy (NICM) patients and displayed an average left ventricular ejection fraction (LVEF) of 20.1 ± 7.3%, compared to 66.8 ± 9.4% in the non-HF group (**table S1**). These donors consisted of 23 males and 13 females, and the mean age of HF and non-HF donors was 57 ± 11.3 and 47.6 ± 15.9 years, respectively. Clinical metadata, including gender as a potential biological variable, were carefully considered and accounted for during data processing and analyses (**see Methods**).

**Fig. 1.**
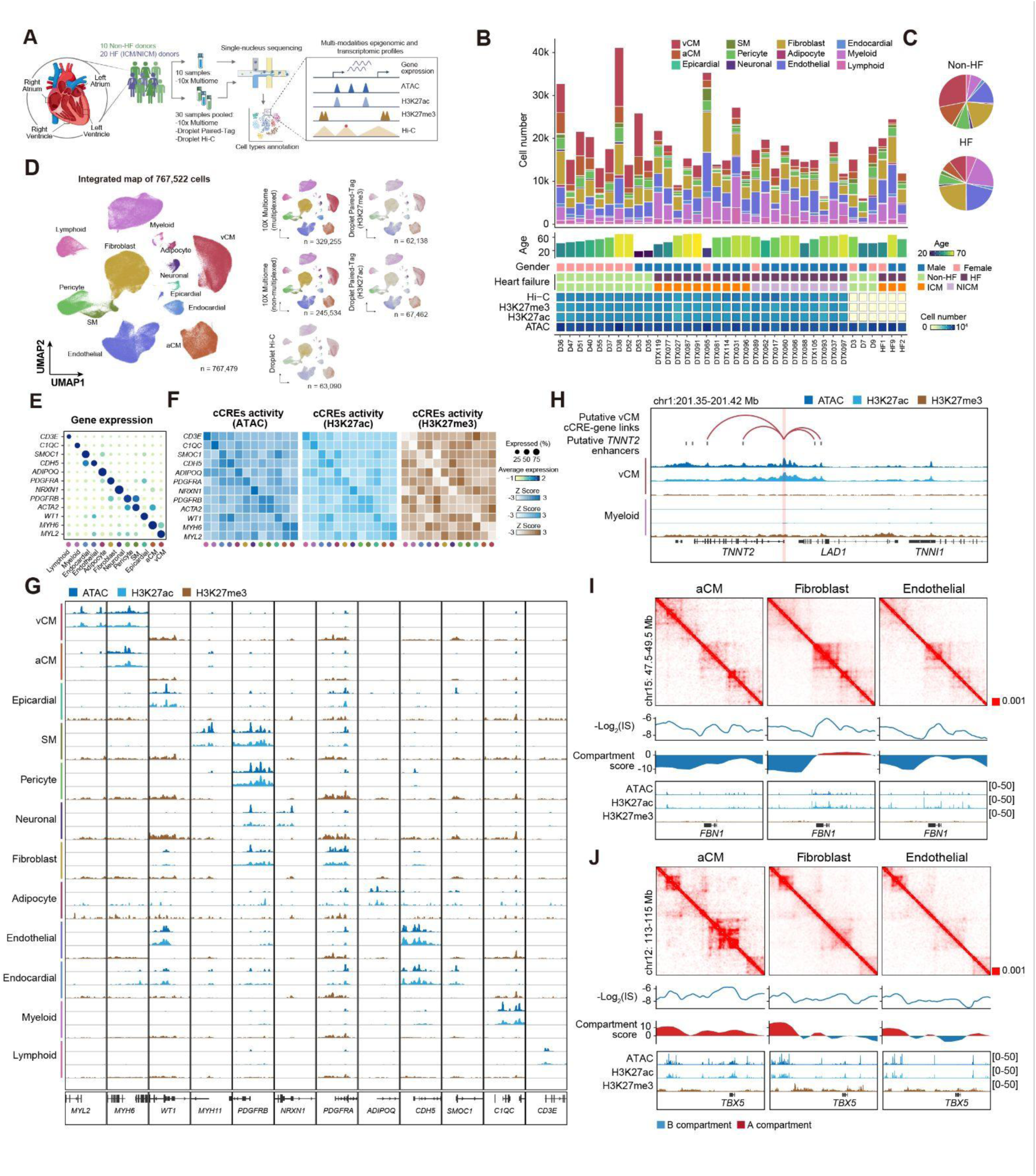
Single-cell multiomic, histone modification and Hi-C analyses uncover cell type-specific transcriptome, epigenome and 3D genome maps of adult human hearts. **(A)** Schematic outlines the workflow of single-cell 10x Multiome, Droplet Paired-Tag, and Droplet Hi-C studies of cardiac chambers from heart failure (HF) and control (Non-HF) donors. HF donors include donors with ischemic cardiomyopathy (ICM) and non-ischemic cardiomyopathy (NICM). **(B)** Stacked bar charts reveal the cardiac cell type proportions for each donor (top). Donor clinical metadata (donor age, gender and condition) along with the number of nuclei profiled for each single-cell sequencing modality is shown below. **(C)** Pie charts display the percentage of cardiac cell types in Non-HF and HF samples. **(D)** Uniform Manifold Approximation and Projection (UMAP) shows the clustering of 767,479 nuclei into major cardiac cell types based on transcriptome or gene activity features from all single-cell sequencing modalities. UMAPs on the right show the nuclei contribution from single-cell sequencing modalities to each major cardiac cell type cluster. **(E)** The expression of selected marker genes is shown in dot plot across the 12 major cardiac cell types. **(F)** Heatmaps display the scaled, averaged epigenetic signals (ATAC, H3K27ac and H3K27me3) for selected marker genes in (E). **(G)** Genome browser tracks reveal aggregated epigenetic profiles for all major cardiac cell types at selected marker gene loci. **(H)** Genome browser track shows an example of cCRE-gene links for *TNNT2* in vCMs (top). Putative *TNNT2* enhancers and epigenomic landscape for vCM versus myeloid lineage are displayed (bottom). **(I, J)** Representative examples of cell type-specific Hi-C contact maps are shown for *FBN1* (I) and *TBX5* (J) loci at 10 kb resolution. –log2 insulation score (IS), compartment score and genome browser tracks for chromatin accessibility and histone modifications are shown below. vCM, ventricular cardiomyocytes; aCM, atrial cardiomyocytes; SM, smooth muscle cell.

A pooled sample strategy was used for our single-cell sequencing studies to cost-efficiently increase assay throughput while reducing potential batch effects (**see Methods, fig. S1A**). To this end, heart tissue samples from 30 donor hearts (10 non-failing, 10 ICM and 10 NICM hearts) were combined into four pools representing each major cardiac chamber for multimodal single-cell sequencing analyses including single-cell multiome (10x chromium single-cell multiome ATAC + Gene Expression), joint single-cell gene expression and histone modification profiling (Droplet Paired-Tag) and single-cell chromatin conformation capture (Droplet Hi-C) (**Fig. 1A-D**) (*12*, *13*). Cells for each donor were identified from pooled samples using genome-wide genotyping information. Individual cells that displayed donor-specific genotype barcodes were observed from all donors in these pooled samples and subsequently processed for downstream analysis (**fig. S1B**). Furthermore, single-cell multiome data was independently generated from the cardiac chambers of 10 additional hearts from both non-HF and HF hearts, including four also used in the pooled single-cell multimodal studies. Comparison with the pooled data confirmed that the pooled sample strategy yielded uniform representation of cells from each donor while maintaining similar data quality, despite yielding a higher number of recovered nuclei per reaction (**fig. S1C-F, table S2**). Finally, correlation analysis of donor profiles confirmed that the pooled sample strategy did not introduce bias in gene expression nor chromatin accessibility data (**fig. S1G**).

To identify the cell types composing the cardiac chambers of these adult human hearts, we initially clustered and annotated 329,255 nuclei from the pooled 10x Multiome data. 12 major cell types were discovered based on the expression of known marker genes (*10*, *14*, *15*) and consisted of atrial and ventricular cardiomyocytes (aCMs and vCMs), fibroblasts, endothelial cells, myeloid, pericyte, lymphoid, endocardial cells, smooth muscle cells (SM), neuronal cells, epicardial cells, and adipocytes (**fig. S2A, B**). Further sub-clustering of these cell types revealed 77 cell subpopulations that ranged from more refined cell subtypes such as capillary, venous and arterial endothelial cells to distinct pathologic cell states for some cell types such as activated fibroblasts (**fig. S2C, D)**. Examining the abundance of these major cardiac cell types revealed their differing proportions between donor hearts, which correlated with the patient’s disease status (**Fig. 1B, C**). Notably, failing hearts displayed increased myeloid cells and fibroblasts but decreased aCMs and vCMs compared to non-failing hearts as previously suggested (*10*, *16*) (**Fig. 1C**). To increase the depth and diversity of transcriptional and chromatin accessibility profiles of these cardiac cell types and their subpopulations, 245,534 nuclei from the single-cell multiome studies on individual (non-pooled) hearts were referenced mapped into the pooled single-cell multiome analyses (**Fig. 1D**).

To annotate accessible chromatin regions and uncover their regulatory interactions across all cardiac cell types, we integrated the pooled single-cell Droplet Paired-Tag data (67,462 nuclei for H3K27ac and 62,138 nuclei for H3K27me3) and pooled Droplet Hi-C data (63,090 nuclei) with the above single-cell multiome data using either gene expression (for Droplet Paired-Tag data) or gene activity score (for Droplet Hi-C data) to create a consensus reference map across modalities (**Fig. 1D, fig. S2E-I, fig. S3A**). As a result, these combined data altogether yielded a single-cell multimodal dataset that in total comprises 767,479 nuclei and specifically consists of 69,944 aCM, 135,564 vCM, 163,977 fibroblasts, 138,045 endothelial cells, 117,065 myeloid, 55,490 pericyte, 36,502 lymphoid, 15,395 endocardial cells, 16,660 SM 10,707 neuronal, 3,424 epicardial, and 4,706 adipocytes (**Fig. 1D, fig. S3B, table S3**). The integrative embedding demonstrated a balanced distribution of nuclei across gender, while disease-specific segregation was preserved, suggesting that gender-related biases were effectively minimized during clustering (**fig. S3C-G**). Detailed cell subpopulation annotations were preserved across the integrated multiomic data, indicating high concordance between sequencing modalities and supporting the integration of these datasets (**fig. S3H**). Correlation analysis using gene expression profiles recapitulated the known hierarchy among major cell types and cell subtypes (**fig. S3I, J**). Consistent with the expression of genes in specific cell types, an enrichment of chromatin accessibility and H3K27ac signals was observed surrounding the transcription start site (TSS) of marker genes specific to each cell type, such as *MYL2*, *MYH6* for cardiomyocytes (CM) and *PDGFRA* for fibroblast (**Fig. 1E-G**). Conversely, these TSS regions were commonly marked by broad, repressive H3K27me3 domains in other cell lineages, likely to ensure the cell type-specific expression of these genes (**Fig. 1F, G, fig. S4A**). To map regulatory elements directing these genes, we predicted enhancers for marker genes in each cell type using the Activity-by-Contact (ABC) model by incorporating the Droplet Hi-C and H3K27ac Droplet Paired-Tag data (*17*). Epigenetic signals over the predicted enhancers, including chromatin accessibility and H3K27ac, were highly correlated with gene expression in corresponding cell types, whereas H3K27me3 was depleted from the putative enhancers in other cell types not expressing the gene of interest (**Fig. 1H, fig. S4B, C)**.

Finally, our single-cell Droplet Hi-C data revealed dynamic and cell type-specific chromatin structural remodeling across the major cardiac cell types, which corresponded with respective cell type-specific transcriptomic and epigenomic changes (**fig. S4D-H)**. Specifically, chromatin organization profiles exhibited cell type-specific domain boundaries (**fig. S4D, E**) and compartment scores that correlated with gene expression variation (**fig. S4F-H**). For example, the genomic loci of *FBN1*, which is primarily active in fibroblasts, exhibited increased H3K27ac activity and was located in an active A compartment in fibroblasts that switched to inactive B compartment in CMs or endothelial cells (**Fig. 1I**). On the other hand, the developmental transcription factor *TBX5*, which is expressed in aCMs, resided within its own cell type-specific TAD-like domain that displayed open chromatin accessibility and strong H3K27ac signal in aCMs. Supporting the strict gene regulation of this transcription factor in a cell type-specific manner, this TAD-like domain in fibroblasts and endothelial cells not only was weakened but also showed an increase in H3K27me3 and B compartment strength (**Fig. 1J**). In summary, our single-cell multimodal dataset provides a unique opportunity to explore the gene regulatory programs that control cell type-specific responses during homeostatic and pathologic conditions of the heart.

### Integrative exploration of single-cell epigenomic studies identifies cell type specific gene regulatory programs of adult human hearts

Integrating multiple single-cell epigenomic profiles enabled not only functional annotation of candidate *cis*-regulatory elements (cCREs), but also classification of gene-regulatory programs across cardiac cell types and conditions. To assess these cCREs and annotate their activities, we initially aggregated chromatin accessibility profiles from 10x single-cell multiome data in all major cardiac cell types and applied non-negative matrix factorization (nNMF) to cluster these cCREs into cell type-related cCRE modules for further chromatin state annotation (**Fig. 2A-D, table S4**) (*18*). As a result, we identified 285,873 cCREs, which covered 2.82% of the human genome and expanded the current annotated cCREs from the ENCODE SCREEN database (*14*) and previous snATAC-seq studies of fetal and adult human hearts (*15*) (**fig. S5A**). Supporting the addition of new cardiac-related cCREs, 49,848 (17.4%) cCREs were previously uncharacterized (**fig. S5A**).

**Fig. 2.**
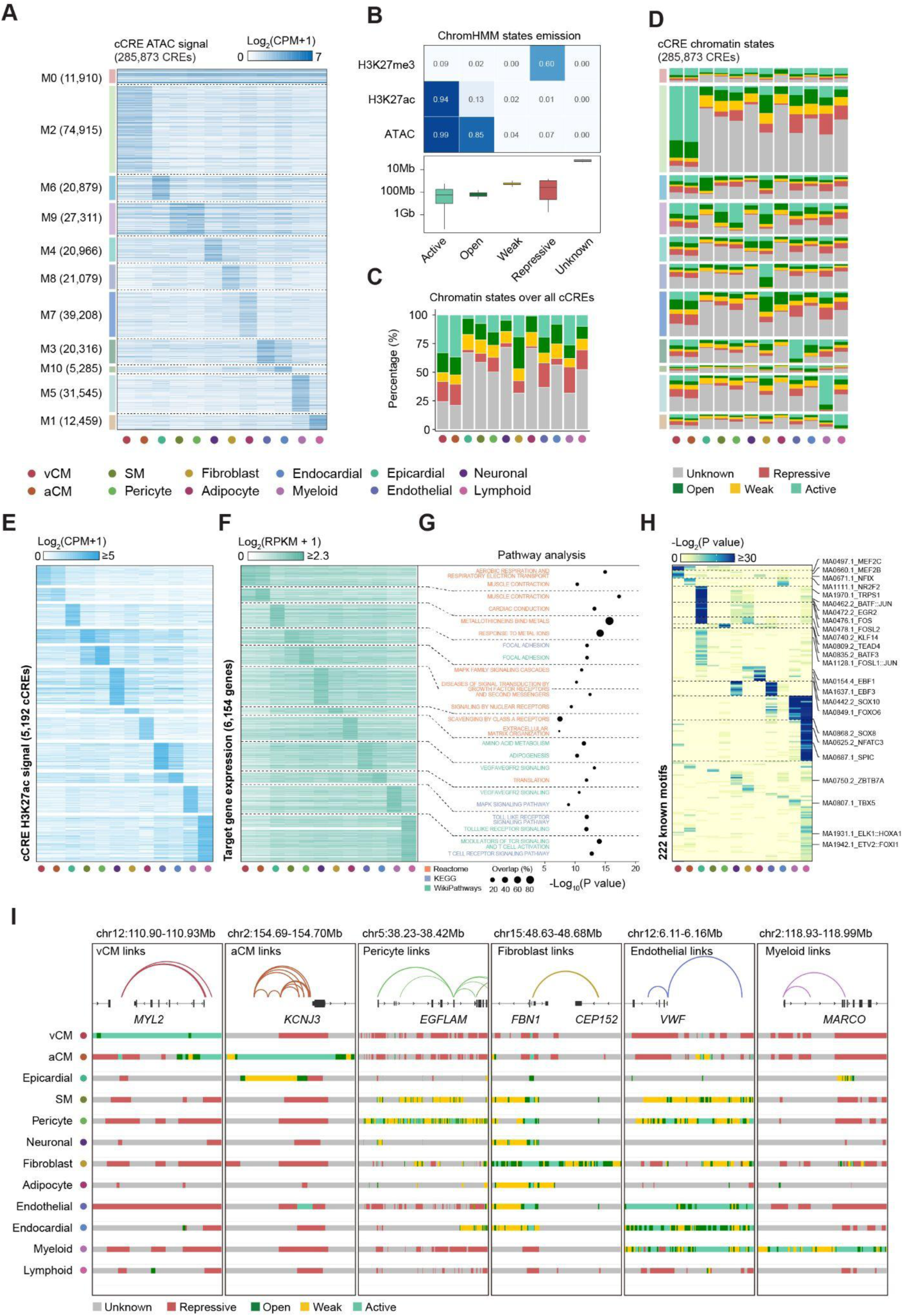
Comprehensive epigenomic profiling of human cardiac cell types reveals their gene regulation. **(A)** Heatmap shows chromatin accessibility signal of cCREs in 11 NMF modules (M0-M10) across 12 major cell types. **(B)** Heatmap shows emission probability of three epigenetic marks across five chromatin states as identified by ChromHMM (top); boxplot shows the genomic coverage of ChromHMM annotated chromatin states (bottom). **(C)** Stacked bar charts show the percentage of cCREs annotated in each chromatin state across all major cell types. **(D)** Stacked bar charts show the proportion of chromatin states over cCREs in each NMF module described in (B) across all major cell types. **(E)** Heatmap shows H3K27ac signal on cCREs in cell type-specific distal cCRE-gene links. **(F)** Heatmap shows gene expression level of target genes associated with cCREs from (E) in cell type-specific distal cCRE-gene links. **(G)** Dot plot shows top enriched GO terms from GSEA analysis of cell type-specific genes targeted by distal cCREs in each cell type. P-values were calculated using a permutation test in GSEA tool. **(H)** 222 HOMER known TF motifs were found enriched in cCREs from cell type-specific distal cCRE-gene links. Example motifs in different cell types are shown. **(I)** Representative genome browser tracks are shown for ChromHMM chromatin states across all major cell types on selected cell type-specific distal cCRE-gene links, including *MYL2* (vCM-specific), *KCNJ3* (aCM-specific), *EGFLAM* (Pericyte-specific), *FBN1* (Fibroblast-specific), *VWF* (Endothelial-specific) and *MARCO* (Myeloid-specific).

Clustering of these cCREs identified 11 distinct gene regulatory modules including 10 modules that displayed cell type-enriched chromatin accessibility patterns, and one module (M0) that exhibited constant chromatin accessibility and higher enrichment for promoter-TSS region (32.7% vs 3.51% in all other modules) across all cell types (**Fig. 2A, fig. S5B**). To further characterize cCRE activity within each module, we applied ChromHMM to derive a five-state model by training on aggregated pseudo-bulk profiles that included chromatin accessibility, H3K27ac and H3K27me3 across the major cardiac cell types (**Fig. 2B, table S5**) (*19*). Based on the emission signals of these chromatin features, we identified five chromatin states including active (high H3K27ac and high ATAC), open (high ATAC and low H3K27ac), weak (low H3K27ac and low ATAC), repressive (high H3K27me3), and an unknown state (**Fig. 2B, fig. S5C, D, table S5**). The unknown state, which displays no detectable signals for all three features, may correspond to different chromatin region types such as unmarked regions or constitutive heterochromatin marked by H3K9me3, which was not included in this study. Although exhibiting fewer chromatin features, the five-state annotations showed close overlap with the 18 chromatin states annotated on adult heart tissues by the ENCODE Epigenome Roadmap (**fig. S5E**) (*20*). Genome-wide annotation using the five-state model revealed that on average 3.75% and 3.02% of the genome were classified as open and active, respectively, compared to on average 0.8% annotated using only cCREs from chromatin accessibility data (**fig. S5C**). When specifically evaluating cCREs rather than the genome, the proportion of open, active and repressive states increased to 14.90%, 15.24% and 11.89% on average, respectively (**Fig. 2C)**. The chromatin states of these cCREs were typically dependent on the cell type, supporting the lineage-specific regulatory roles of cCREs in cardiac cell types. Consistent with this notion, many cCREs displaying active and open states were present specifically in cell type-enriched gene regulatory modules (except for M0); however, these cCREs could also be observed to be reciprocally repressed in other cell type-enriched gene regulatory modules where cCREs were not active or open (**Fig. 2D**). On average, 75.1% of these repressive cCREs were open or active in at least one other cell type (**fig. S5F**), and GREAT analysis revealed that they were mostly associated with cellular functions in distinct cell lineages, thus revealing how gene expression may be actively repressed to ensure tight regulation across cardiac cell types (**fig. S5G**). In summary, our integrative analysis of multimodal single-cell data provides a comprehensive framework for characterizing cCRE dynamics, thus providing the foundation for investigating gene regulatory networks (GRNs) across diverse cardiac cell types and disease conditions.

To identify cCREs specific to each cell type, we performed a likelihood ratio test to identify variable cCREs and then calculated a cCRE specificity score. cCREs were classified as cell type specific if their specificity score was significantly higher than background (empirical FDR < 0.05, see Methods) and can be attributed to only one cell type. This approach identified 20,267 cell type specific cCREs, which further clustered into ten major classes that generally corresponded with specific cardiac cell types (**table S6**). However, two distinct cCRE classes were discovered to be associated with vCMs, aCMs as well as pericytes, SMs, supporting how these cell types may be closely related as reported (*9*) (**fig. S6A, table S6**). Based on H3K27ac and chromatin state annotation, the majority of these cCREs were active in corresponding cell types (**fig. S6A, B**). On average, 86.3% of the cell type-specific cCREs were classified as active across all major cardiac cell types, with a lower ratio in epicardial cells and adipocytes, which may be due their inadequate cell coverage (**fig. S6B**). Motif enrichment analysis identified 333 known transcription factor (TF)-binding motifs with cell type-specific enrichment patterns reflecting known TF activities in each cell type (**fig. S6C, table S7**). For example, we detected enrichment of myocyte enhancer factor MEF2A and cardiac developmental factor MEIS2 in cardiomyocyte-specific cCREs, SOX4 in endocardial-specific cCREs and SPIB in myeloid-specific cCREs (**fig. S6C**). Additionally, gene set enrichment analysis (GSEA) (*21*, *22*) revealed that these cell type-specific cCREs were associated with genes involved in distinct cardiac functions, such as “cardiac muscle contraction” for cardiomyocyte-specific cCREs (K1/2) and “T-cell receptor signaling pathway” for lymphoid-specific cCREs (K10) (**fig. S6D, table S8**). Together, these analyses uncovered cell type-specific cCREs that may serve as enhancers to mediate the expression of essential genes in distinct cardiac cell types.

To link distal cCREs with their putative target genes, we applied the ABC model and identified 66,317 distal cCREs-promoter pairs across all cell types (**fig. S6E-H, table S9**), including 11,938 cell type-specific distal cCRE-gene links based on overlap with at least one cell type-specific distal cCRE (**Fig. 2E, F, table S6)**. These cell type-specific links displayed strong enrichment of chromatin contacts and active chromatin states in respective cell types (**fig. S6B, I**). Genes targeted by these cell type-specific links exhibited concordant expression in cognate cell types. GSEA of these genes further identified pathways closely related to the function of corresponding cell types (**Fig. 2G, table S10**), such as vCM-specific distal cCREs linked to genes involved in muscle contraction. Additional motif analysis of cCREs within these links identified 222 known TF-binding motifs that were associated with known TFs that are critical for the function of each related cell type as similarly observed in cell type-specific cCREs (**Fig. 2H, table S11**). As examples of these distal cCRE-gene links, we observed that the vCM specific gene *MYL2* was linked to a cCRE >10 kb downstream; the aCM-specific gene *KCNJ3* was linked to multiple cCREs spanning 10 kb upstream; and the endothelial gene *VWF* was linked to a cCRE ∼30 kb upstream (**Fig. 2I**). Altogether, these findings highlight how cell type specific epigenetic landscapes orchestrate the precise regulation of gene expression in each cell type.

### Global gene regulatory and chromatin structural changes of cardiac cell types during heart failure

Our multimodal single-cell analyses provide the opportunity to comprehensively interrogate the cell type-specific epigenomic and chromatin architecture changes that underlie the dynamic gene expression programs of distinct cardiac cell types during HF. To this end, we identified in total 10,400 differentially expressed (DE) genes and 52,738 differentially accessible (DA) cCREs in failing versus non-failing hearts using DESeq2 with a FDR cutoff of 0.05 (*23*). These DA cCREs were particularly enriched in cell types including CMs (4,373 in aCM, 28,557 in vCM), fibroblasts (18,292) and myeloid cells (4,121), and correlated with changes in gene expression in these cell types during HF (**Fig. 3A, fig. S7A-D, tables S12, S13**). Further annotation of these cCREs with cell type-specific epigenomic information revealed the complex regulation of cCRE activities. Despite ubiquitous changes in gene expression across different cell types, epigenetic changes were most prominent in CMs, fibroblasts, pericytes, endothelial cells and myeloid cells, supporting the significance of these cell types during HF (**Fig. 3A**). In particular, differential H3K27ac activity typically correlated with ATAC-seq signal changes for many cCREs; however, in vCMs, a substantial number of cCREs with increased H3K27ac activity (52.31%) did not show an increase in chromatin accessibility during HF, suggesting that these cCREs may have been open and primed in non-failing vCMs but become activated in failing vCMs (**fig. S7E, table S14)**. Furthermore, we also identified 37,924 genomic regions (15 kb) with differentially regulated H3K27me3 signals (**Fig. 3A, table S14**). Notably, we found that most cell types except for CMs displayed more down-regulated than upregulated H3K27me3 signals, suggesting that H3K27me3 may globally decrease to lead to derepression of specific genetic programs during HF (**Fig. 3A**).

**Fig. 3.**
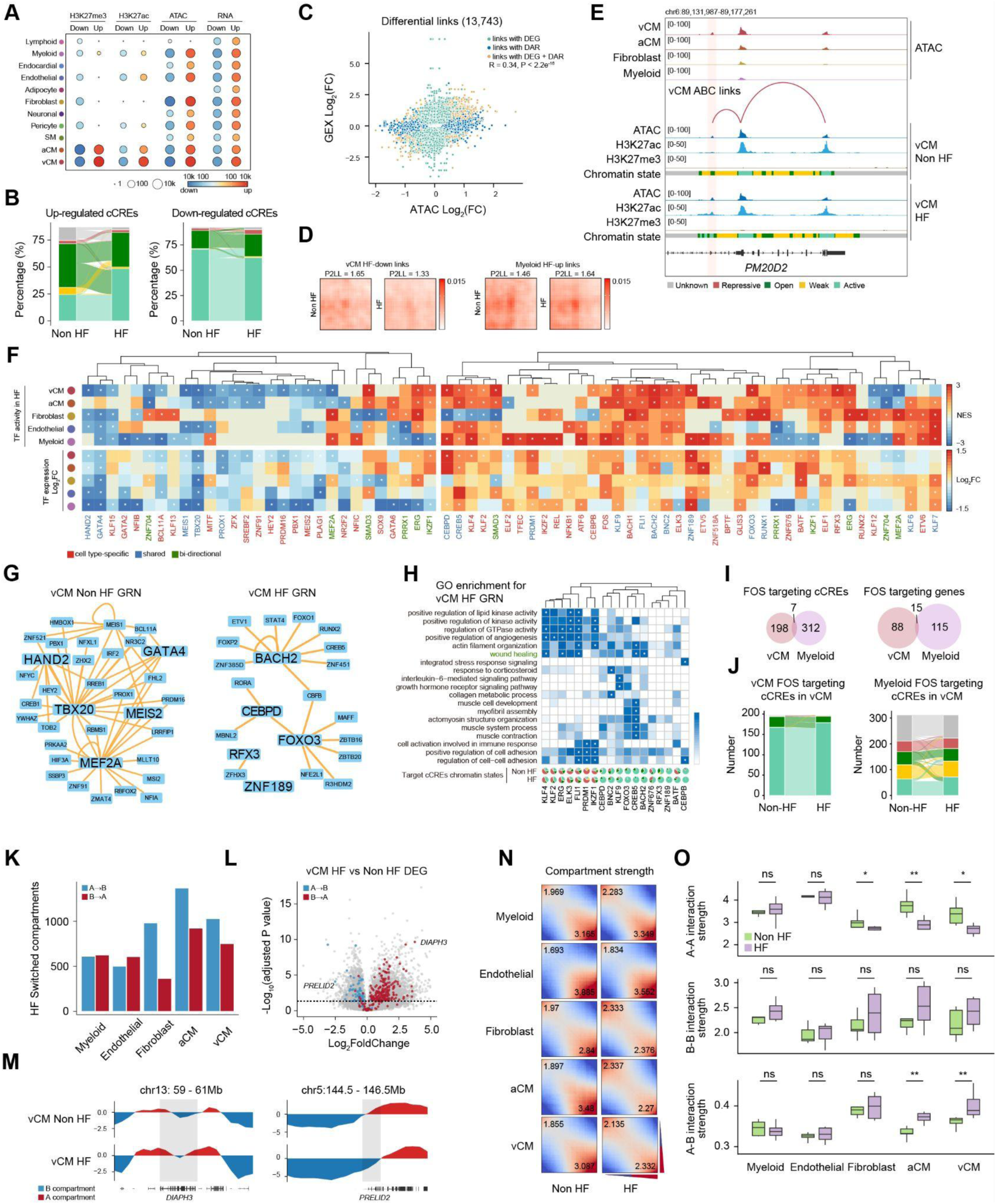
Cell type-specific epigenetic changes direct gene regulatory responses during heart failure. **(A)** Dot plot shows the number of genomic regions and genes identified from differential analysis between heart failure (HF) and non-heart failure (Non-HF) conditions across cell types. **(B)** Sankey plots show the chromatin state dynamics for differentially accessible cCREs in HF and Non-HF hearts. Percentage was calculated as the averaged percentage of chromatin states across cell types. **(C)** Scatter plots compare the log2 fold changes of gene expression (GEX) and chromatin accessibility (ATAC) for genes and cCREs from putative ABC links between HF and Non-HF conditions. Links are colored based on whether the associated gene and/or cCRE is classified as differentially expressed (DEG) or differentially accessible (DAR). **(D)** Normalized aggregate peak analysis (APA) scores of Droplet Hi-C signals in vCM and myeloid cells are shown for vCM down-regulated and myeloid up-regulated ABC links. APA scores are represented as P2LL (peak-to-lower-left) values. **(E)** Genome browser tracks show an example of a differential ABC link targeting *PM20D2* in vCM. Pseudobulk chromatin accessibility tracks across selected cardiac cell types (top) as well as differential epigenetic signals and chromatin states in HF versus Non-HF vCMs (bottom) are shown. **(F)** Heatmap shows the enrichment of HF-or Non-HF-associated transcription factor (TF)-regulated gene regulatory networks (GRNs) calculated from fGSEA across selected cell types (top). Only GRNs with absolute normalized enrichment score (NES) ≥ 2 and with consistent differential TF expression results are shown. Asterisk indicates significant enrichment (adjusted P-value < 0.05), with P-values estimated based on an adaptive multi-level split Monte-Carlo scheme in *fgsea* package. Log2 fold changes of each TF corresponding to the respective GRN are also shown (bottom). Asterisk indicates significant enrichment (adjusted P-value < 0.05), calculated using Wald test in *DESeq2* package and corrected for multiple testing using Benjamini-Hochberg FDR correction. **(G)** Network plots illustrate representative vCM GRNs in Non-HF versus HF conditions. **(H)** Heatmap shows enrichment of gene ontology (GO) terms for HF-associated TF GRN-targeted genes in vCMs (top). Pie charts below show the chromatin state percentage for HF-associated GRNs-targeting cCREs under Non-HF and HF conditions (bottom). Asterisk indicates significant enrichment (adjusted P-value < 0.05). P-values were calculated using hypergeometric test in enrichGO and corrected for multiple testing using Benjamini-Hochberg FDR correction. **(I)** Venn diagrams show the overlap of FOS target genes and cCREs in vCMs versus myeloid. **(J)** Sankey plot shows the dynamic changes of vCM chromatin states of FOS-targeted chromatin regions in HF versus Non-HF conditions for vCMs and myeloid. **(K)** Bar chart indicates the number of switched compartments in HF versus Non-HF for selected major cell types. **(L)** Volcano plot shows the log2 fold changes and adjusted P-value of differentially expressed genes in vCMs between Non-HF and HF conditions. Genes that overlap with switched compartments are colored in blue (A to B) or red (B to A). Example genes shown in (M) are labeled. P-values were calculated and corrected as described in (F). **(M)** Representative genome browser track of compartment scores in HF versus Non-HF vCMs for *DIAPH3* and *PRELID2*. Compartments that shifted are shaded in grey. **(N)** Saddle plots show compartmentalization strength comparison in selected major cell types between Non-HF and HF conditions. Numbers indicate A-A (bottom right) and B-B (top left) compartment interaction strengths. **(O)** Boxplots compare intra-(A-A or B-B) and inter-compartment (A-B) interaction strengths in Non-HF versus HF cell types in young-aged (≤60 years) donors.

The majority of DA cCREs (44,367, 84.13%) were distal to promoter regions (defined as 1 kb upstream and 500 bp downstream of annotated TSSs) of protein-coding and non-coding RNA genes. Interrogating the chromatin states of these distal DA cCREs revealed that their upregulation was typically associated with chromatin states switching from open to active, whereas their downregulation was associated with the decrease of active chromatin states (**Fig. 3B**). Subsequent motif analyses on these distal DA cCREs identified 328 and 374 known motifs enriched in up-regulated and down-regulated distal DA cCREs during HF, respectively (**fig. S7F**). While these motifs were enriched in specific cell types, we also discovered some motifs that were shared across cell types including the stress-response AP-1 family TF-binding motif (JUN, FOS and ATF), thus supporting that a common stress response gene regulatory program may be broadly activated across many cardiac cell types during HF. In contrast, more distinct TF-binding motifs such as MEF2A and ESRRB were enriched in HF-down regulated cCREs in vCMs but not aCMs, suggesting that vCMs may undergo more dynamic gene regulatory changes than aCMs during HF (**Fig. 3A, fig. S7F**).

To discover gene targets for distal DA cCREs, we overlapped our cell type-specific cCRE-gene links with cCREs that differentially exist between non-HF and HF hearts. As a result, we identified target genes for these distal DA cCREs, which included *NKX2-5*, *ADRB2* and *CKM* in CMs, *NPPC* and *FAP* in fibroblasts, and *NLRP3*, *CD163* in myeloid lineages (**table S15**). Supporting their regulation by these distal DA cCREs, putative target genes were more likely to be also differentially expressed between non-HF and HF conditions (**Fig. 3C**). Further interrogation of these distal cCRE-gene links revealed a concordance between the changes in cCRE-gene links and Hi-C signal (**Fig. 3D**). As an example, we observed that the *PM20D2* gene, which is specifically upregulated in CMs during HF, was linked to a distal cCRE located 5 kb upstream, which was altered from an open to an active chromatin state in HF vCMs. Consequently, this interaction was associated with increased H3K27ac and active chromatin at the promoter of *PM20D2* (**Fig. 3E**). Gene ontology analysis was further performed on these HF-associated cCRE-targeted genes to understand their roles in HF. Genes in up-regulated links were enriched in a cell type-specific manner in pathways related to muscle adaptation in vCMs and extracellular matrix and structure organization in fibroblasts, whereas those associated with down-regulated links were related to biological processes such as collagen metabolic process in fibroblasts or heart contraction in vCMs (**fig. S7G**). In summary, these findings reveal how distal cCREs regulate target gene activities that influence the function of specific cell types during HF progression.

Identifying TFs involved in directing cCRE-gene regulation is critical for understanding how gene regulatory programs are established to control cell type specific responses under diverse conditions. The integration of our single-cell multiomic RNA-seq and ATAC-seq data enabled the construction of GRNs to discover key TFs regulating cell type-specific responses during HF (**fig. S7H, tables S16, S17**) (*24*). In particular, combining TF expression levels as well as corresponding cCRE accessibility changes in each cell type identified GRNs with altered activity across each major cell type in non-HF and HF conditions (**Fig. 3F**). Using this strategy we identified 41 cell type-specific TFs as well as 23 shared TFs, including six TFs that exhibit divergent activity depending on the cell types, which we termed “bi-directional” (**Fig. 3F).** For instance, during HF, cell type-specific BACH2*-* and RUNX2-regulated GRNs were specifically activated in CMs and fibroblasts, respectively, whereas the HEY2-regulated GRN was selectively down-regulated in vCMs (**Fig. 3F)**. Despite overall transcriptional similarities between aCM and vCMs (**fig. S3I, J**), we also observed divergences in the GRN landscapes of these CMs during HF, which included differentially activated cCREs within each TF regulon (**fig. S7I**). Notably, for a shared TF like BACH2, over 70% of its target cCREs were distinct between aCM and vCM. These cell type–specific BACH2-targeted cCREs also exhibited different levels of activation in HF (**fig. S7J-L**). We further investigated the bi-directional TF MEF2A, which was associated with HF fibroblasts and non-HF vCMs (**fig. S7M**). Chromatin state analysis revealed that MEF2A-bound cCREs in vCMs were largely active under non-HF conditions but showed reduced activity in HF, while its target cCREs in fibroblasts displayed increased activation in HF (**fig. S7N**). This bi-directional regulatory pattern was corroborated by the expression levels of MEF2A target genes in vCMs and fibroblasts that corresponded with their respective cell type–specific functions (**fig. S7O, P**).

Additional analysis of vCM GRNs illuminated the dynamic changes of GRNs and TF regulatory landscape between non-HF and HF conditions (**Fig. 3G**). Gene ontology analysis of the regulons in the HF-associated GRN in vCMs revealed target genes that shared similar biologically-related pathways including wound healing, regulation of GTPase activity and positive regulation of kinase activity; however, the cCREs targeted by these TFs displayed different activities between non-failing and failing hearts (**Fig. 3H**). In contrast to these cell type specific GRNs, some TFs, such as FOS, displayed elevated GRN activity across multiple cell types, although its expression level was only significantly increased in endothelial cells (**Fig. 3F**). Despite the increased GRN activities across multiple cell types, FOS was predicted to bind to cCREs and regulate target genes in a cell type-specific manner (**Fig. 3I, J, fig. S7Q**). GO term enrichment results showed that the FOS-targeted genes in different cell types were not only associated with stress response signaling but also closely related with cell type-related functions (**fig. S7R**). In line with these findings, chromatin state annotation revealed that FOS-targeted cCREs were largely open or active in vCMs, but displayed more heterogeneous chromatin states in myeloid lineages (**Fig. 3J**). Notably, the open FOS-targeted cCREs in vCMs exhibited further increased activity including active chromatin states during the transition from non-failing to failing heart conditions, suggesting that these cCREs may be primed during non-failing conditions but may become activated upon cardiac stress or HF (**Fig. 3J**). Altogether, these findings uncover key transcriptional programs in HF across many major cardiac cell types including CMs, thus highlighting the distinct gene regulatory changes underlying HF progression.

Chromatin interaction profiles not only facilitated identification of cCRE-gene links but also revealed the global organization pattern of nuclei. Using Droplet Hi-C data, we systematically compared the chromatin architecture heterogeneity between non-HF and HF conditions across the major cardiac cell types. To this end, we investigated the genome-wide chromatin compartment score of each cell type at 100-kb resolution, which correlated with the underlying sequence activity. Across the five most abundant cell types, we identified in total 5,221 genomic bins that underwent compartment switch events (**Fig. 3K**), which showed global enrichment of differential cCREs and genes (**Fig. 3L, fig. S8A**). For example, the vCM-specific gene *PRELID2* was among the top down-regulated genes during HF, and its genomic region switched from A to B compartment in HF vCMs (**Fig. 3M**). On the other hand, the up-regulation of *DIAPH3* gene expression in vCMs correlated with its genomic region switching from B to A compartment. Notably, we also observed a higher proportion of compartments switched from A to B in CMs and fibroblast (**Fig. 3K**). Finally, we observed significantly increased cross-compartment interactions specific to CMs (**Fig. 3N, O, fig. S8B-D)**, possibly indicating a disruption of nuclear architecture in these cell types during HF. These increased cross-compartment interactions were associated with the gain of extreme long-range interactions in HF conditions especially for B compartments (**fig. S8E, F**). We also noticed that the loss of short-range interactions in A compartments is unique to vCMs rather than aCMs, although both cell types exhibited decreased A compartment interaction, indicating that their chromatin organization changes may be different (**Fig. 3O, fig. S8E, F**). The CM– and fibroblast-specific decrease in A-A interaction frequency are consistent with the loss of insulation in these cell types during HF, suggesting that enhancer-promoter interactions are more likely to be rewired in these cell types (**Fig. 3O, fig. S8G**).

Overall, our findings present a comprehensive, disease– and cell type-specific map of gene regulation programs in both failing and non-failing hearts. The genomic profiles categorized by modalities, cell types and disease states will be a valuable resource for the biomedical community.

### Cardiomyocytes display a broad spectrum of cellular states in control and diseased hearts

Consistent with cell types that may be affected during HF, CMs displayed the most heterogeneity among the major cardiac cell types in non-diseased and diseased hearts (**Figs. 4, 5, figs. S9-13**). In particular, we identified 13 vCM and 7 aCM subpopulations, which expressed vCM-specific marker *MYL2* and aCM-specific marker *NR2F2* (*9*, *25*), respectively, within corresponding cardiac chambers of all analyzed hearts (**Fig. 4**, **fig. S9**). While all CM subpopulations were present in non-HF and HF hearts, these CM subpopulations derived from all cardiac conditions in different abundances (**Fig. 4B**, **fig. S9A, B**, **fig. S14A-C**).

**Fig. 4.**
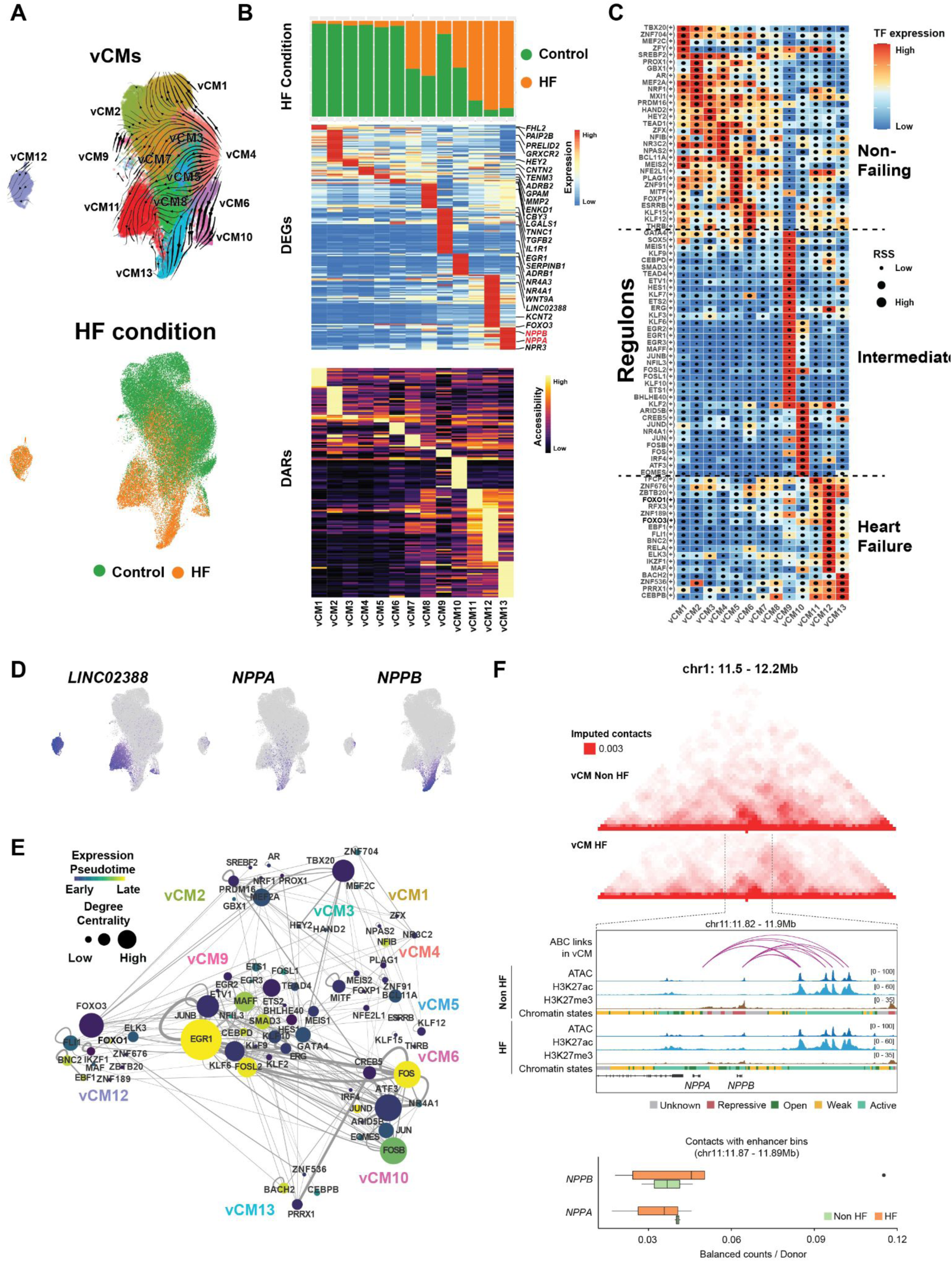
Gene regulatory network analysis of ventricular cardiomyocytes uncovers genetic programs controlling their cellular states. (**A**) UMAPs show vCM cell subpopulations and their RNA velocity derived trajectories (top) and the contribution of HF and non-HF vCMs to each subpopulation (bottom). (**B**) Each vCM cell subpopulation exhibits distinct cellular contributions (top, bar graph) from each heart condition as well as gene expression (middle, heatmap) and chromatin accessibility (bottom, heatmap). (**C**) SCENIC+ TF GRNs for each vCM cell subpopulation is shown in heatmap. The box is colored by transcription factor (TF) expression, and the dot size shows the regulon specificity score (RSS). (**D)** UMAPs display the gene expression of marker genes for distinct vCM disease states. (**E**) Analyzing the TF network within the vCM GRN inferred the interactions between distinct GRNs across non-HF and HF vCM subpopulations. Nodes are colored by pseudotime of expression. Node size is determined by their degree of centrality in the network, and edges represent TF-TF connections. (**F**) Hi-C contact maps (top) and representative genome browser tracks at *NPPA* and *NPPB* locus display pseudobulk epigenetic signals, chromatin states and ABC links associated with *NPPA* and *NPPB* in HF versus non-HF conditions. Box plot of balanced Hi-C contacts in non-HF versus HF conditions reveals decreased enhancer-*NPPA* contacts and increased enhancer-*NPPB* contacts in HF vCMs (bottom).

Focusing on the ventricles, which are the primary cardiac chambers affected during HF, we identified six vCM subpopulations primarily from non-failing ventricles (vCM1-vCM6), three vCM subpopulations mainly from failing ventricles (vCM11, vCM12, vCM13) and four vCM subpopulations containing vCMs appreciably from both non-failing and failing vCMs (vCM7-vCM10) (**Fig. 4A, B, fig. S14A-C**). Furthermore, vCMs from ICM and NICM hearts were proportionately present in HF-related subpopulation vCM7, vCM8, vCM11, vCM12, vCM13, whereas diseased vCMs in vCM9 and vCM10 were from predominantly ICM but not NICM hearts (**fig. S14C**). In line with these findings, vCM9 and vCM10 were transcriptionally similar to vCM subpopulations observed in human myocardial infarction and ischemic hearts, which may represent an intermediate stress/pre-disease state as reported (*26*).

Among non-failing related vCM subpopulations (vCM1-vCM6), many genes and chromatin regions were commonly upregulated and accessible including *MYL2* and *TBX20* genes but genes and accessible chromatin regions present in failing-related vCM states (vCM11-vCM13) were downregulated (**Fig. 4B, C, tables S18, S19**). Of the highly disease-enriched vCM subpopulations (vCM11, vCM12, vCM13), two transcriptionally distinct vCM states were identified based on gene expression and chromatin accessibility patterns. In particular, vCM13 specifically expressed HF-related and inflammatory-related genes, *NPPA*, *NPPB* and *TGFB2, IL1R1*, respectively, as reported (*26*), whereas vCM12 displayed upregulated genes related to DNA repair and metabolism including *FOXO1*, *FOXO3*, as well as *LINC02388*, which has been suggested to promote blood vessel formation in response to hypoxia (*27*) (**Fig 4B, D, table S19**). Additionally, vCM11 displayed genes and accessible chromatin regions that were specific to either vCM12 or vCM13 but at different levels. In contrast, vCM7-vCM10, which consisted of vCMs from both non-diseased and diseased ventricles, broadly displayed genes and accessible chromatin regions that were observed differentially upregulated in either non-disease or disease-enriched vCMs but at lower levels (**Fig. 4B, table S19**), supporting that these vCMs may represent intermediate/transition vCM states during HF as reported for vCM9 and vCM10 (*26*). Notably, we discovered that stress response, wound healing and inflammatory related genes were highly upregulated in vCM9 and vCM10 including *SERPINB1, TGFB2, IL1R1*, and *CCN1* (F**ig. 4B****, tables S19, S20**), suggesting that inflammatory and wound healing pathways regulating activated fibroblasts (*28*, *29*) may also initiate adaptive stress/disease responses in vCMs. In line with this notion, vCM10 highly expressed *NR4A1* and *NR4A3*, which regulate cardiomyocyte hypertrophic responses to cardiac stress and are key regulators of known cardiac neurohormonal factors including the renin-angiotensin-aldosterone system/RAAS (e.g. Angiotensin II) and the sympathetic nervous system (e.g. β-adrenergic signaling) (*30*). Illuminating the disease relationship of these vCM subpopulations, scVelo RNA velocity analysis across all vCM subpopulations revealed major disease state trajectories initiating from non-disease related vCMs (vCM1-6), spanning intermediate vCM disease states (vCM7-10) and ending at vCM11-vCM13 disease states (**Fig. 4A, table S21**).

SCENIC+ (*24*) was applied to the integrated single-cell multiome data for all cardiac cell subpopulations to predict their changes in gene regulatory programs including TF and chromatin accessibility (**Fig. 4C, 5C, fig. S9-13, table S17**). Building upon our cell type-specific HF GRN analyses (**Fig. 3F, fig. S7H**), we further interrogated how gene regulatory programs of identified vCM states may be regulated across conditions including non-failing, intermediate and failing states (**Fig. 4C**). In particular, several known CM-specific TFs including GATA4, HEY2, MEF2A, MEF2C and TBX20 were expressed and displayed high target gene accessibility in non-failing related vCM states, but were downregulated across intermediate and disease-related vCM states, suggesting a potential loss of vCM identity during cardiac disease/stress. Conversely, we identified distinct sets of transcriptional regulators that progressively increased their expression from non-HF to HF-enriched vCM states including FOXO3 (vCM12) and CEBPB (vCM13), a TF reported to regulate adaptive CM hypertrophic and injury responses (*31*, *32*) (**Fig. 4C**). Corresponding to the expression of stress response and wound healing genes in vCM9 and vCM10, JUN, FOS, ATF, EGR and SMAD related TFs were particularly elevated in these intermediate vCM states, supporting that the activation of these stress response gene programs are transient but may initiate a cascade of gene programs that eventually lead to end/chronic disease vCM states (**Fig. 4C**). As such, we discovered that vCM9 and vCM10 displayed large complex GRNs where their TFs displayed high centrality scores indicating their elevated interconnectedness with other TFs within and between the GRNs of vCM subpopulations including those from both non-diseased and diseased vCM states (**Fig. 4E**).

**Fig. 5.**
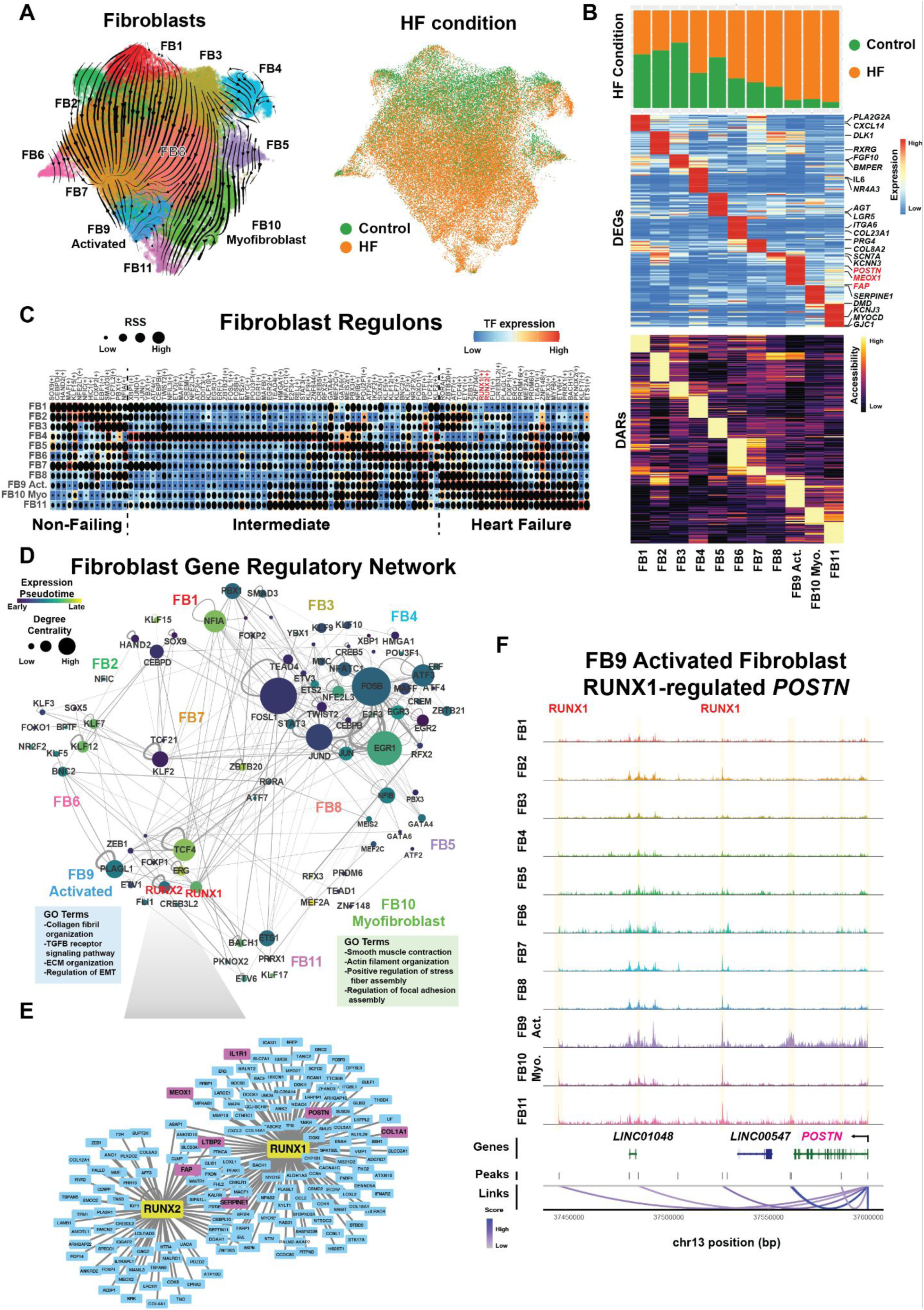
Gene regulatory network analysis of cardiac fibroblasts reveals genetic programs directing their cellular states. (**A**) UMAP shows fibroblast cell (FB) subpopulations and their trajectories as detected by RNA velocity (left). UMAP labeled by HF condition reveals the contribution of non-HF and HF fibroblast cells to each subpopulation (right). (**B**) Each cardiac fibroblast cell subpopulation displays distinct cellular contributions (top, bar graph) from each heart condition as well as gene expression (middle, heatmap) and chromatin accessibility (bottom, heatmap). (**C**) Heatmap shows SCENIC+ predicted TF GRNs for each cardiac fibroblast cell subpopulation. The box is colored by transcription factor expression, and the dot size shows the regulon specificity score (RSS). TFs in red are associated with FB9-Activated Fibroblasts. (**D**) Interrogating the TF network within the cardiac fibroblast GRN identifies interactions between distinct GRNs across cardiac fibroblast states. Nodes are colored by pseudotime of expression. Node size is determined by their degree of centrality in the network, and edges represent TF-TF connections. Gene Ontology (GO) terms for the FB9-Activated Fibroblasts and FB10-Myofibroblasts are shown. (**E**) Network plot of the RUNX1 and RUNX2 regulons shows their target genes including known gene markers of activated fibroblasts (colored in magenta). (**F**) Representative genome browser tracks at *POSTN* locus display aggregated chromatin accessibility profiles of each FB subpopulation and co-accessible peaks linked to *POSTN* (below, loops), including two SCENIC+ predicted peaks for the RUNX1 regulon.

Integrating these vCM-specific gene regulatory programs with corresponding vCM-specific epigenetic signals and chromatin 3D-structure enabled illumination of how pathologic vCM gene programs may become activated during HF (**Fig. 4F, fig. S14D, E**). Notably, we discovered that HF-related genes *NPPA* and *NPPB*, which tandemly reside within the same genomic locus on chromosome 1, were dynamically and reciprocally regulated in non-HF and HF vCMs (**Fig. 4F**). Consistent with the low expression of *NPPA* but negligible expression of *NPPB* in non-failing vCMs (**Fig. 4D, fig. S14D**), the chromatin state of *NPPA* was weak, but *NPPB* was repressed (**Fig. 4F**). ABC analyses and vCM-specific Hi-C contact maps further revealed that a super-enhancer ∼40 kb upstream of *NPPB* interacted with the promoter of *NPPA* to control its activity as previously suggested (*33*) (**Fig. 4F**). While the chromatin state for both genes became active in failing vCMs (**Fig. 4F**), the expression of *NPPA* was lower than that of *NPPB* (**fig. S14D**). This expression pattern corresponded to the altered Hi-C contact maps within the *NPPA-NPPB* locus of failing vCMs, which revealed reciprocal interactions of the super-enhancer with *NPPA* and *NPPB* (**Fig. 4F**).

### Gene regulatory programs activating diseased fibroblasts

Cardiac fibroblasts (FBs) actively participate in the overall maintenance and remodeling of the heart under a wide range of cardiac conditions throughout life. Supporting the dynamic role of cardiac FBs in regulating overall cardiac homeostasis and disease, 11 cardiac FB states were identified and notably contributed across all cardiac conditions (non-HF and HF) (**Fig. 5, fig. S15A-C**). Over 50% of the cardiac FBs from FB4, FB6-FB12 subpopulations were from diseased hearts, especially FB9, FB10 and FB11 subpopulations, suggesting that these diseased-enriched FBs may be involved in stress responses during cardiac disease (**Fig. 5A, B, fig. S15A-C**). In line with this notion, diseased-associated FBs differentially expressed genes involved in inflammation and fibrosis such as TGF-β signaling, ECM, wound healing and stress response genes (**Fig. 5B, tables S19, S20**). In particular, FB9 and FB10 displayed genes including *POSTN, FAP, MEOX1* and *DMD, SERPINE1* that were noted to be specifically upregulated in activated fibroblasts and myofibroblasts, respectively (*28*) (**Fig. 5B**). While we identified several cardiac FB states (e.g., FB9, FB10) that were recently reported (*28*), we observed that FB11, a small subpopulation of fibroblasts that was related to FB10 and localized more in the atria than ventricles, expressed genes related to ion channels including *KCNJ3* and *GJC1* (*CX45)*, supporting a potential electrically-active FB as recently suggested (**Fig. 5B, fig. S15C**) (*34*). Similar to CM subpopulations, we also observed cardiac FB states that expressed genes that were observed in both non-failing– and failing related FB states, suggesting that these FB populations may exhibit intermediate disease states (**Fig. 5B, table S19**). Supporting these findings, scVelo RNA velocity uncovered two major cardiac fibroblast trajectories that specifically directed cardiac FBs to either an activated fibroblast (FB9) or myofibroblast (FB10, FB11) state and included intermediate-disease FB subpopulations expressing related genes to each FB disease state (**Fig. 5A, B, table S21**) as suggested (*28*).

To illuminate the gene regulatory programs that may direct how cardiac FBs may transition to disease-related states, we further examined the changes in TF activity as well as chromatin accessibility across all cardiac FB subpopulations (**Fig. 5C**). We observed that the Wnt-related TF TCF7L2 and its accessibility for chromatin regions were elevated in non failing-associated FBs but gradually decreased along each disease trajectory as FBs progress from intermediate to disease-associated FB subpopulations (**Fig. 5C**) as previously described (*35*). On the other hand, we discovered TFs that specifically increased along specific disease trajectories including EMT-related RUNX1, RUNX2 and FGF-related ETV1 in the FB9 activated fibroblast subpopulation and MEF2A/C, TEAD1 and MYF6 in the FB10 myofibroblast subpopulation (**Fig. 5C, fig. S15D**). Notably, we discovered that FB3 displayed upregulation of the TGF-β related TF SMAD3, and FB4 specifically expressed many stress response TFs such as FOS, JUN and ATF related TFs and may be a key transitory state as cardiac FBs become either activated fibroblasts or myofibroblasts (**Fig. 5C**).

Additional GRN and epigenomic analysis revealed how identified TFs may control the TF network directing the transition of FBs along distinct disease trajectories (**Fig. 5D, fig. S15E**). FOS and JUN related TFs, which were enriched in FB4, were highly connected to many TFs in not only FB4 but also nearly every fibroblast state as determined by GRN centrality scores, further supporting that FB4 may be a key intermediate state for FB disease progression (**Fig. 5C, D**). As such, FB4 exhibited the largest GRN among the different FB states, which included FB4-related TFs that may direct FB4 to either FB9 activated fibroblasts or FB10 myofibroblasts (**Fig. 5C, D**). Consistent with these findings, FB4-as well as FB5-related TFs were identified to regulate RUNX1 and other TFs in FB9 activated fibroblasts and MEF2A/TEAD1 in FB10 myofibroblasts (**Fig. 5D, fig. S15F, G**). Conversely, the TGF-β related TF SMAD3 in FB3 subpopulations was linked to FB4-related TFs, suggesting that FB3 may represent an early pre-stressed FB state which precedes FB4 as the scVelo FB trajectory analysis predicts (**Fig. 5A, C, D**).

Further interrogating the GRNs of FB9 and FB10 illuminated pathologic genetic programs regulating activated fibroblasts and myofibroblasts, respectively. Within the FB9 GRN, we observed that RUNX1 along with RUNX2 may regulate known activated fibroblast-associated genes including *POSTN, MEOX1* and *FAP* (**Fig. 5E, F**) (*28*). While these genes were expressed in FB9 (**Fig. 5B, fig. S15D, table S19**), RUNX1 displayed transcriptional activity and expression in not only FB9 but also FB4 (**Fig. 5C, fig. S15D**). These findings indicate that RUNX1 may serve as a pre-disease transcriptional regulator which translates early cardiac stress responses to the broad initiation of pathologic fibroblast gene programs, as suggested (*36*, *37*); however, *RUNX2* and *MEOX1*, whose expression are more restricted to FB9 (**fig. S15D**), may direct specific expression of genes in activated fibroblasts as reported (*28*, *37*). Supporting this notion, we observed that RUNX1 was regulated by both FB9-related TFs as well as FOS, JUN and other stress-response TFs in FB4, whereas RUNX2, which was downregulated in FB4, was controlled primarily by FB9-related TFs (**fig. S15F**). On the other hand, we discovered that MEF2A and TEAD1 may regulate known myofibroblast-associated genes such as *MYH9*, *TNS1* and *YAP1* in FB10 myofibroblasts (*38–40*) (**fig. S15G, H**). These myofibroblast-related TFs were discovered to be regulated by MEF2C and NFIB in FB5, respectively (**fig. S15I**), suggesting that FB5 may be a precursor state of myofibroblasts. Thus, these findings reveal a hierarchy of gene regulatory networks that direct the transition of cardiac FBs into distinct pathologic FB states during HF.

### Genetic effects on heart cell types and risk of heart failure

To discover how genetic variants may impact distinct cardiac cell types to affect the risk of HF and other cardiovascular traits, we analyzed our cardiac cell type gene regulatory programs with GWAS data from a wide range of cardiovascular traits **(table S22)**. To this end, we tested for enrichment of genetic variants associated with cardiovascular traits in cCREs using LD score regression (*41*). We identified significant (FDR < 0.1) enrichment of trait-associated variants in the cCREs of distinct cell types, which notably grouped based on the type of cardiovascular trait (**Fig. 6A, table S23**). For instance, genetic variants associated with HF, dilated cardiomyopathy (DCM), cardiac arrhythmia, and cardiac trait measurements were strongly enriched in CM-specific cCREs (**Fig. 6A**). In contrast, traits related to hypertension, blood pressure, myocardial infarction and other cardiovascular diseases were associated with cCREs in non-cardiomyocyte cell types including fibroblasts and SMs (**Fig. 6A, table S23**). Notably, non-ischemic HF-associated (HF, DCM) genetic variants were particularly enriched in cardiomyocyte cCREs whereas those in ischemic HF (HF, myocardial infarction) were enriched in fibroblast and SM cCREs, suggesting potentially different pathologic mechanisms for these two forms of HF (**Fig. 6A, fig. S16A, table S23**). To further define these associations, we examined the chromatin states of these cell type cCREs harboring genetic variants. cCREs in active states displayed cell type-specific patterns of enrichment across traits, whereas those in open states exhibited more limited enrichment, supporting that genetic trait enrichments may be primarily driven through active cCRE states (**Fig. 6B, table S24**).

**Fig. 6.**
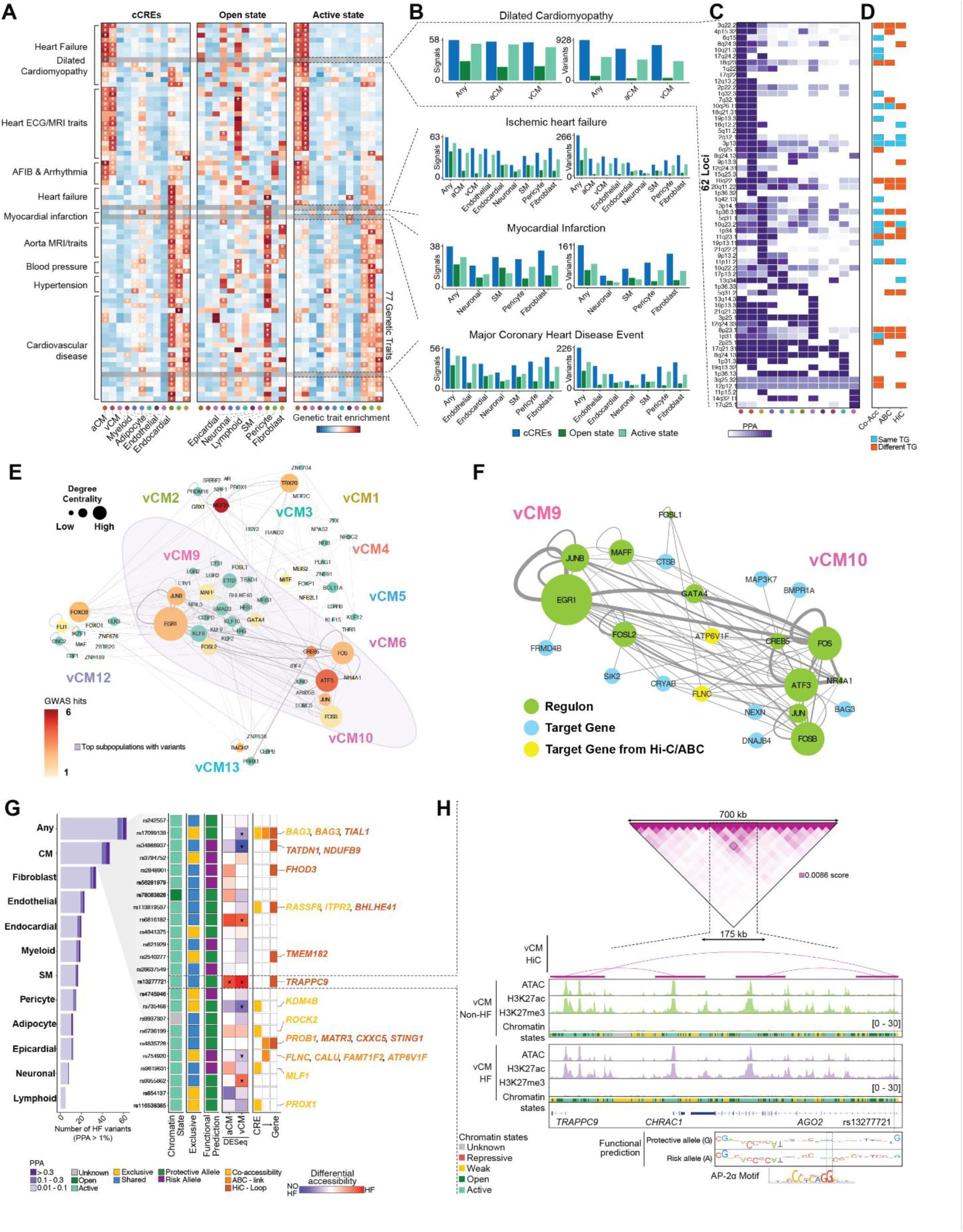
Fine-mapping of genetic loci associated with cardiovascular traits and heart failure. **(A)** Heatmap displays LD score regression (LDSC) enrichment of trait-associated variants in all cCREs as well as cCREs in the open (Open) or active (Active) chromatin state across distinct cardiac cell types. Color represents the level of enrichment. **(B)** Bar plots quantify the number of fine-mapped variants (Variants) and loci (Signals) overlapping all, open and active cCREs in LDSC-enriched cell types. **(C)** Heatmap represents the cumulative posterior probability association (PPA) values of fine-mapped variants associated with dilated cardiomyopathy (DCM) across cardiac cell types. The cytogenetic band of the lead variant was used as the locus identifier. **(D)** Heatmap indicates whether predicted target gene annotations coincide with the prioritized genes from Zheng et al. (*7*). SCENIC+ inferred links (Co-Acc), ABC links, or Hi-C loops were used to assign putative target genes. A target gene annotation was considered different if a method did not find a prioritized target gene from Zheng et al. **(E)** Network plot shows the vCM subpopulation GRN overlap with cCREs harboring fine-mapped variants. Node size represents the centrality score from SCENIC+, while node color intensity indicates the number of variants overlapping GRN-associated cCREs (nodes in teal have no overlapping variants). **(F)** Network plot shows the GRNs and target genes overlapping GWAS variants for the two vCM subpopulations containing the highest number of GWAS variants (vCM9 and vCM10). **(G)** Stacked bar chart represents the overlap of fine-mapped DCM genetic variants with cCREs across cardiac cell types (left). Colors indicate PPA ranges: 0.01–0.1, 0.1–0.3, and >0.3. Heatmaps display the overlap between variants and chromatin state classification, exclusivity of variant localization within CM cCREs, ChromBPNet functional predictions, and a log2 fold-change heatmap of differential chromatin accessibility between HF and non-HF aCMs and vCMs from DESeq2 (FDR < 0.1, right). Additionally, cCRE-to-gene links are visualized based on co-accessibility, ABC, or Hi-C methods. The colorimetric scheme applies to both boxplots and listed target genes, distinguishing each annotation method. **(H)** Hi-C contact map displays the interactions between a cCRE harboring the HF risk variant rs13277721 and the *TRAPPC9* promoter (top). Genome browser tracks show aggregated chromatin accessibility, histone modifications and chromatin state heatmap for non-HF and HF vCMs (middle). Motif plot shows ChromBPNet functional predictions comparing the protective allele (G) and risk allele (A), highlighting disruption of an AP-2α motif by the HF risk allele (bottom).

We then used cardiac cell type cCREs to identify candidate causal genetic variants associated with different cardiovascular traits using fine-mapping data (**Fig. 6B**, **fig. S16A, table S24**). Overall, most known loci for each cardiovascular trait were linked with cCREs in active states for disease-enriched cell types (**Fig. 6B**). Based on a recent GWAS study identifying a large set of DCM risk loci (*7*), we focused specifically on HF due to DCM. As a result, 1,293 fine-mapped DCM genetic variants representing 77.5% (62/80) of known DCM loci were located within a cardiac cell type cCRE of which genetic variants at 94% (58/62) of these 62 loci overlapped with a CM-specific cCRE, supporting the enrichment of DCM-genetic variants in CMs (**Fig. 6C, D**, **fig****. S16B-D**). We then clustered these 62 DCM loci based on the cumulative probability of fine-mapped genetic variants overlapping cCREs in each cell type, which revealed many loci associated with CMs and smaller subsets of loci linked with fibroblasts and other cell types (**Fig. 6C, table S25**). Next, we identified potential target genes of these DCM loci in CMs by combining ABC links, SCENIC+, and chromatin loop information, and discovered that 33 DCM loci were linked to at least one predicted target gene. Of these loci, 22 loci had predicted target genes that differed from those in previous studies (*7*) (**table S26**), including target genes highly distal to risk variants such as *GATA4* and *MYH7B*, highlighting potential novel biological links to HF (**Fig. 6D**, **table S26**).

To understand how genetic risk of HF may impact GRNs controlling CM states, we overlaid fine mapped DCM-associated genetic variants onto our CM GRN maps. As a result, these fine-mapped genetic variants preferentially overlapped GRNs controlled by specific TFs, suggesting convergence of pathways altered in HF in cardiomyocytes (**Fig. 6E, F**). In particular, we identified an enrichment of HF variants residing in GRNs of specific vCM states, particularly the intermediate states (vCM9, vCM10) which are controlled by stress-response related TFs including JUN, FOS and ATF3 (**Fig. 6E, F, table S27**). These GRNs were associated with genes involved in stress response, inflammatory, and disease pathways, suggesting that the genetic variants may impact responses to cardiac stress (**fig. S16E, table S28**). To further understand specific subpopulations/states of CMs that may drive HF risk, we tested for HF enrichment of genetic risk variants in GRNs active in CM subpopulations. We identified enrichment of DCM-associated variants in GRNs for intermediate states (vCM10) (log enrich=4.2) using *fgwas*, and no corresponding evidence for enrichment for other states (all log enrich<0), thus further supporting the critical role of cardiac stress responses in driving HF (**table S29**).

Finally, we explored more closely how fine-mapped genetic variants may impact cCREs and target genes in cardiac cell types to regulate HF/DCM risk. Given the strong enrichment of HF variants for CM cCREs, we focused specifically on variants affecting CMs. We initially prioritized candidate variants in CM-specific cCREs with predicted effects on CM chromatin accessibility using ChromBPNet (**Fig. 6G, table S30**). We then identified target genes of prioritized variants using ABC links, SCENIC+, and chromatin loops. Our results not only provided support for previously implicated target genes of HF variants, such as rs17099139 variant for *BAG3* at the 10q26.11 locus or variant rs754920 for *FLNC* at locus 7q32.1, but also revealed the specific CM cCREs that may be affected to mediate disease risk (**Fig. 6G, fig. S16F, G**). Furthermore, we also identified many target genes of HF variants not identified in previous studies (*7*). For example, candidate HF risk variant rs13277721 at locus 8q24 mapped in an active distal cCRE predicted to regulate *TRAPPC9* over 100 kb upstream away based on chromatin looping in CM Hi-C data (**Fig. 6H**). This candidate HF variant rs13277721 was predicted to alter cCRE activity through reducing chromatin accessibility and disrupting an AP-2α sequence motif (**Fig. 6H**).

Overall, integrating cardiac cell type regulatory programs with human genetic association data revealed cell types, gene networks, cCREs, and target genes of HF and cardiovascular disease risk which may provide new therapeutic targets for disease.

## Discussion

Our multiomic single-cell sequencing studies offer new insights into the coordinated transcriptomic and epigenomic landscape of the distinct cell types comprising non-failing and failing human hearts as well as the chromatin organization/interactions that wire the gene regulatory networks controlling cell type-specific enhancer-promoter interactions. Expanding upon single-cell RNA sequencing studies that have previously identified these cell types (*9*, *10*, *26*, *42–44*), our epigenomic data, including Droplet Paired-Tag, Droplet Hi-C and single-cell multiome, reveal additional layers of complexity for how gene programs are regulated in specific cell types during homeostatic and pathologic heart conditions. In particular, these integrative analyses enabled not only the interrogation of chromatin states for regulatory elements in distinct cell types but also the prediction of distal enhancers for genes of interest and definition of cell type-specific enhancer-promoter interactions. As a result, we identified genes that were dynamically regulated by diverse sets of distal enhancers, which exhibit distinct chromatin states across cell types and disease conditions. These distal enhancers, bound by unique combinations of transcription factors, regulated target genes in a cell type-specific manner, thus enabling the precise regulation of gene expression across distinct cell lineages. Among the affected cell types, CMs and fibroblasts displayed significant changes in not only gene expression and chromatin accessibility, but also global chromatin architecture during HF. Notably, the chromatin compartmentalization in CMs was substantially perturbed, supporting previous reports of CM nuclear remodeling during HF (*45*, *46*). These chromatin architecture changes, reminiscent of those observed in cancer and developmental disorders (*47*, *48*), may reflect HF regulatory mechanisms that alter nuclear organization and function in CMs and other cardiac cell types.

Additionally, deeper interrogation of these cell types uncovered multiple distinct intermediate and pathologic cellular states that may direct the responses of each cell type during HF. Through network analyses of our integrated single-cell multiomic data, we identified key gene regulatory programs and transcription factors that govern the transition of these cell types from non-disease to intermediate– and end-stage disease states. Consequently, these findings provide insights into how pathologic gene programs may be activated and how homeostatic gene programs may be suppressed in a cell type-specific manner during HF. For example, our results highlighted molecular mechanisms driving the reactivation of fetal cardiac genes (e.g., *NPPA*, *NPPB*) during HF, which promotes pathologic cardiac hypertrophy and remodeling, but not reparative cardiomyocyte proliferation as observed in fetal hearts (*49*).

Finally, this comprehensive atlas of human cardiac cell type-specific epigenomes and regulatory interactomes enabled the systematic interrogation of how non-coding variants may influence gene expression, enhancer activity, and disease susceptibility in a cell-type-specific manner. Thus, our findings provide a valuable, explorable resource for prioritizing candidate cardiovascular-disease associated genetic variants discovered from GWAS and whole genome sequencing efforts (*5–8*). This prioritization will aid in the functional examination and identification of causal genetic variants at scale using emerging high-throughput genetic screening assays (*50–52*).

Collectively, these insights offer a potential framework for guiding the development of new cell type-specific strategies aimed at directing specific cardiac cell types from a pathologic to a reparative state. Overall, our studies illuminate the diversity of cell types in failing human hearts and the chromatin dynamics that establish gene regulatory programs contributing to the pathogenesis of HF with reduced ejection fraction (HFrEF). However, they may also offer new insight into potentially other presentations of HF because of the overlap in the pathophysiology of different forms of HF (*53*).

## Data Availability

All data produced in the present study are available upon reasonable request to the authors

## Acknowledgments

The authors would like to thank the University of Utah patients and their families for providing the valuable myocardial tissue donations used in this study. We thank the Chi, Drakos, Gaulton and Ren laboratories for comments, and the University of California San Diego (UCSD) Institute for Genomic Medicine. We also thank JD Hocker and S Preissl for comments and help with single-nuclei isolation from heart tissue.

## Funding

This project was supported by:

Foundation of NIH (KJG, BR, NCC, SGD)

AHA Heart Failure Strategically Focused Research Network, 16SFRN29020000 (SGD) NIH NHLBI R01 HL135121 (SGD)

NIH NHLBI 1R01HL166513 (SGD)

Merit Review Award I01 CX002291 (SGD)

U.S. Department of Veterans Affairs (SGD)

Nora Eccles Treadwell Foundation (SGD)

NIH HG012059 (KJG)

NIH NHLBI T32HL007444 (ENF, ARH)

NIH-1K08HL168315-01 (ET)

Life Sciences Research Foundation (ZW)

## Author contributions

Conceptualization: SGD, KJG, BR, NCC

Methodology: YX, LC, AW

Investigation: YX, ENF, LT, QY, ST, JD, JB, HZ, ED, WE, SC, RME, SM, ZW, JC, RM, CM, EG, JL

Visualization: YX, ENF, LT, ST, HZ, ARH, JB, DL, CS, TW

Data Curation: LC, TSS, SP, AL, TL, QY Funding acquisition: SGD, KJG, BR, NCC

Resources: TSS, ET, VH, QZ, SN, CHS, SGD, NCC

Project administration: YX, ENF, LT, LC, TSS, ADC

Supervision: YX, ENF, LT, AW, SGD, KJG, BR, NCC

Writing – original draft: YX, ENF, LT, KJG, BR, NCC

Writing – review & editing: YX, ENF, LT, LC, ARH, ED, AW, SGD, KJG, BR, NCC

## Supplementary Materials

Materials and Methods

Figs. S1 to S16

References (*54–90*)

