## Supplementary Materials for "Single cell multiomics and 3D genome architecture reveal novel pathways of human heart failure"

### Materials and Methods

#### Study Population and Ethics Statement

Patients with end-stage cardiomyopathy who underwent heart transplantation at the institutions comprising the Utah Transplantation Affiliated Hospitals Cardiac Transplant Program (i.e., University of Utah Health Science Center, Intermountain Medical Center, and the Veterans Administration Salt Lake City Health Care System) were enrolled in this study after obtaining informed consent. Control tissue was acquired from non-failing donors whose heart was not allocated for transplantation due to non-cardiac reasons. Patients with acute forms of HF, defined by symptoms <3 months of duration with no evidence of LV dilation, were excluded. Subjects with hypertrophic or infiltrative cardiomyopathies were also excluded from this study. The study was approved by the institutional review board of the participating institutions (IRB 00030622).

Chronic ischemic cardiomyopathy (ICM) was defined as a left ventricular ejection fraction (LVEF) <40% and any of the following: 1) a history of MI or revascularization; 2) a history of angina or chest pain and evidence of scarring in noninvasive imaging studies corresponding to previous MI; 3) presence of ≥75% stenosis of the left main or proximal left anterior descending artery; or 4) presence of ≥75% stenosis of ≥2 epicardial vessels in a patient with unexplained cardiomyopathy. Patients with a LVEF <40% and nonobstructive coronary artery disease without evidence of prior MI or revascularization were considered to have non-ischemic cardiomyopathy (NICM) as previously described (*54*).

#### Myocardial Tissue Acquisition

Transmural cardiac samples were collected from all 4 chambers of the heart (left ventricle (LV), right ventricle (RV), left atrium (LA), and right atrium (RA)) and snap frozen in liquid nitrogen at the time of heart transplantation and stored at –80 °C. A total of 13 ICM, 10 NICM and 13 non heart failure (HF) donor samples were used for this study.

#### Sample pooling

For each pool, we selected 15 or 30 donors to represent all condition groups and gender. For each donor, approximately 20-30 mg of snap frozen tissue (tissue samples measuring approximately 3 mm X 3 mm X 3mm) was sectioned on dry ice using pre-chilled scalpels and collected in a pre-chilled 5 ml LoBind tube on dry ice. Sectioned tissue pools were stored in the -80°C freezer.

#### Heart nuclei isolation

For single sample and pooled sample Multiome, single-nuclei suspensions were prepared from frozen tissues using gentleMACS™ Dissociator (Miltenyi Biotec) in MACS buffer (5mM CaCl2 (R040, G-Biosciences), 3mM Mg-acetate (M2545, Sigma), 10 mM Tris-HCl pH 8.0 (15568025, Thermo Fisher Scientific), 2mM EDTA (15575020, Thermo Fisher Scientific), 0.6 mM DTT (D9779, Sigma), 1X Roche cOMPLETE Protease Inhibitor EDTA-Free (11873580001, Sigma), 1.5 U µl− RNasin® Ribonuclease Inhibitor (N2515, Promega). The nuclei suspension was then filtered through 100-µm and 30-µm CellTrics™ Filters to remove debris and centrifuged for 5 min at 500g at 4°C. Nuclei pellet was resuspended in iodixanol buffer (50% and 25% iodixanol (OptiPrep™ Density Gradient Medium (D1556-250ML, Sigma), Dilutent Buffer (120mM Tris-HCl, pH = 8 (15568025, Thermo Fisher Scientific), 150 mM KCl (AM9640G, Invitrogen), 30 mM MgCl2 (194698, Mp Biomedicals Inc), Molecular biology water (46000-CM, Corning)) and centrifugation for 30 min at 4,000g at 4°C. After resuspension in sort buffer, nuclei were stained with 2 µM 7-AAD in sort buffer (1X PBS, 1X protease inhibitor, 1.5 U µl− RNasin® Ribonuclease Inhibitor and 1% BSA) for 10 min on ice and were sorted using SH800 cell sorter for single nuclei. Nuclei were collected in collection buffer (1X PBS, 5X protease inhibitor, 7.5 U µl− RNasin® Ribonuclease Inhibitor and 5% BSA) and centrifuged for 10 min at 500g at 4°C.

For pooled sample Droplet Paired-Tag, single-nucleus suspensions were also prepared using gentleMACS™ Dissociator (Miltenyi Biotec) in MACS buffer as with multiome but with 1 U/µl RNaseOUT and 1 U/µl SUPERaseIn inhibitor. After filtering using 100-µm and 30-µm CellTrics™ Filters and centrifugation,Anti-Nucleus MicroBeads (Miltenyi Biotec) were used for debris removal and enrichment of nuclei. Nuclei suspension was centrifuged for 5 min at 500g at 4 C. After resuspension in sort buffer, nuclei were stained with 0.2mg/ml Hoescht in sort buffer (1X PBS, 1X protease inhibitor, 1 U/µl RNaseOUT and 1 U/µl SUPERaseIn inhibitor) and 1% BSA) for 10 min on ice and were sorted by fluorescence-activated nuclei sorting with an SH800 cell sorter for single nuclei. Nuclei were collected in collection buffer (1X PBS, 5X protease inhibitor, 5 U/µl RNaseOUT, 5 U/µl SUPERaseIn inhibitor, and 5% BSA).

For pooled Droplet Hi-C, nuclei were fixed immediately following dissociation using the gentleMACS™ Dissociator in MACS buffer, as described above.

<u>10x Genomics Epi Multiome ATAC + Gene Expression assays.</u>

The nuclei pellet was resuspended in permeabilization buffer (1mM DTT, 0.2% IGEPAL-CA630, 1X cOmplete EDTA-free protease inhibitor, 1.5 U µl−1 RNasin and 5% BSA in PBS), incubated on ice for 2 min and centrifuged for 5 min at 500g and 4°C and resuspended in 1x nuclei buffer (20x Nuclei buffer (PN 2000207, 10x Genomics), 1 mM DTT (D9779, Sigma) Molecular biology water (46000-CM, Corning). Suspension of 30,000 permeabilized nuclei were each loaded onto 8 lanes and processed using a 10x Genomics Controller following manufacturers recommendations. 10x Genomics Epi Multiome ATAC + Gene Expression assays were processed following manufacturers recommendations. Generated libraries were sequenced at the Institute for Genomic Medicine at UC San Diego using NovaSeq X Plus (Illumina) sequencer to the depth of ~25.6B reads for ATAC-seq and ~26.5B reads for RNA-seq.

<u>Genotyping arrays and imputation</u>

Genomic DNA was extracted from 10–15 mg sections of frozen tissue obtained from each of the 30 donors. DNA isolation was performed using the Monarch Genomic DNA Purification Kit (NEB #T3010L), with elution in 70–100 µL of UltraPure Distilled Water (Invitrogen 10977-015). When required, DNA concentrations were adjusted to 50 ng/µL using a speed vacuum concentrator.

Genotyping was conducted at the UCSD IGM Genomics Center using the Illumina HumanCoreExome-24v1 array. Genotype calling, cluster positioning, and extraction of SNP statistics were performed with GenomeStudio v2.0.5, employing the PLINK input report plugin with an hg38 cluster file for reference. The resulting genotypes were converted into PLINK format within GenomeStudio.

For downstream analysis, genotyping data from different arrays were merged using PLINK v1.9. Quality control filtering was applied as follows: variant missing call rate $\leq 0.05$, minor allele frequency (MAF) $\geq 0.01$, and Hardy-Weinberg equilibrium (HWE) p-value $> 1 \times 10^{-5}$. Prior to imputation, variant positions and annotations were standardized using the HRC-1000Gcheck-bim.pl script (v4.3.0) from the McCarthy Group Tools collection (https://www.chg.ox.ac.uk/~wrayner/tools/).

Droplet Paired-Tag data generation

To obtain joint profiles of histone modifications and transcriptomes from single cells in heart samples, Droplet Paired-Tag was performed as previously described (*12*), with minor modifications. In brief, we pooled cardiac chamber tissues from 30 different donors in equal weights and extracted nuclei using the same protocol as our 10x Genomics single-cell multiome data generation. The nuclei were sorted via fluorescence-activated nuclei sorting using an SH800 cell sorter (Sony) to isolate single nuclei. The isolated nuclei were collected and counted using a cell counter (RWD C100-Pro) with DAPI staining. We aliquoted 500,000 nuclei to initiate reactions in individual tubes for each histone modification mark and replicate. Two replicates were included for each tissue pool and histone modification.

First, nuclei were permeabilized with OMNI buffer and incubated with 2 μL PA-Tn5 (0.4 mg/mL) and 2 μg antibody in MED#1 buffer overnight at 4 °C (*55*). PA-Tn5 and antibodies were then removed by washing with MED#2 buffer. Tagmentation was carried out in MED#2 buffer supplemented with 10 mM $MgCl_2$ (Invitrogen, AM9530G) at 550 r.p.m. and 37 °C for 60 minutes in a ThermoMixer (Eppendorf), and the reaction was terminated by adding 2× stop solution. The nuclei were washed in 1× nuclei buffer and counted again. From each tube, 24,000–30,000 nuclei were aliquoted and used for a single droplet generation reaction with the Chromium Next GEM Single Cell Multiome kit (10x Genomics, 1000283). Finally, DNA and RNA library amplification was performed according to the Chromium Single Cell ATAC Library kit manual, with the following modifications: the starting amount of preamplification product for DNA library amplification was doubled, 13 amplification cycles were used for the DNA library, and a different SPRI bead size selection were applied for the DNA library (50 μL + 105 μL).

For this study, we used antibodies targeting two histone modification marks: H3K27ac (Abcam, ab4729, polyclonal) and H3K27me3 (Abcam, ab192985, recombinant).

Droplet Hi-C data generation

To obtain cell type-specific chromatin architecture, we performed Droplet Hi-C on all the heart samples from different donors, as previously described (*13*). In brief, heart samples from 30 donors were pooled together, and nuclei were extracted within the same reaction. Nuclei were fixed with paraformaldehyde (Electron Microscopy Sciences, 15714) at a final concentration of 2%, and then quenched with glycine solution at a final concentration of 0.4 M. Nuclei were permeabilized with lysis buffer on ice for 30 minutes. Then, the nuclei were treated with 0.5% SDS (Promega, V6551), incubated at 62 °C for 10 minutes on a ThermoMixer, and then quenched with Triton X-100 (Sigma, 93443) at 550 r.p.m. and 37 °C for 15 minutes. Chromatin in the nuclei was sheared using three restriction enzymes: DpnII (NEB, R0543L), MboI (NEB, R0147M), and NlaIII (NEB, R0125L), at 37 °C for 90 minutes on a ThermoMixer at 550 r.p.m. The three enzymes were then deactivated at 65 °C for 20 minutes, and the mixture was cooled to room temperature. The sheared chromatin was further ligated using T4 DNA ligase (NEB, M0202L). All enzymes were removed from the nuclei, which were then suspended in 1% BSA in 1x PBS with 7-AAD (Invitrogen, A1310) and incubated on ice for 10 minutes. The nuclei were sorted into collection buffer (5% BSA in 1xPBS) via fluorescence-activated nuclei sorting using an SH800 cell sorter (Sony) to remove all the debris.

The sorted nuclei were collected by centrifugation and washed twice with 1x nuclei buffer. Approximately 25,000 nuclei were aliquoted into each PCR tube, and one reaction was performed using the Chromium Next GEM Single Cell ATAC Reagent Kits v2 (10x Genomics). The tagmentation incubation time was 60 minutes, and the index PCR elongation time was extended from 20 seconds to 1 minute. Double-sided size selection was adjusted to 1.14x SPRIselect to remove only small fragments. For each cardiac chamber pool, we performed 2-3 independent reactions.

Preprocessing of Droplet Paired-Tag data

Fastq files from Droplet Paired-Tag were demultiplexed using the 'mkfastq' command in cellranger-arc (v2.0.0). After demultiplexing, histone modification and transcriptome modalities were aligned to reference genome (hg38) separately using the 'count' command in cellranger-atac (v2.0.0) and cellranger (v6.1.2), respectively. To select high-quality nuclei, we adopted a three-step filtering strategy: First, histone modification data were aggregated at the sample level, and peak calling were performed to identify narrow peaks (for H3K27ac) or broad peaks (for H3K27me3) using MACS2 (v2.1.2) (*56*). Per-cell fragment number and the Fraction of Reads in Peaks (FRiP) were used to filter and retain barcodes with high-quality histone modification profiles. For RNA, barcodes were filtered based on UMI counts per cell. Next, nuclei were selected by pairing the pass-filtered barcodes from both modalities. Lastly, based on donor genotype phasing results, only pass-filtered nuclei confidently assigned to a single donor were retained for downstream analysis. Ambient RNA contamination was estimated and removed using SoupX (*57*) for each library separately before clustering.

Genome track visualization

Cell type- or condition-specific alignments for histone modification data were extracted from cellranger bam files using custom scripts. BigWig files were generated with the 'bamCoverage' function in deepTools (v3.5.1) (*58*) using the normalization method 'RPGC', with chrY excluded for normalization.

For chromatin accessibility data, valid alignments were extracted, split into single-end reads, and corrected for Tn5 insertions: reads aligned to the positive strand were shifted +4 bp, and reads aligned to the negative strand were shifted -5 bp. Each alignment was further shifted -100 bp (for the positive strand) or +100 bp (for the negative strand) and then extended to a length of 200 bp to capture the Tn5 insertion points. BigWig files were generated using the 'bamCoverage' function with the same normalization method and exclusion criteria as described above for the histone modification data.

Integration of Droplet Paired-Tag data with 10x Multiome

We used the RNA modality from both multiomic assays (10x Multiome and Droplet Paired-Tag) as anchors to perform integration and annotate cell type identities using Seurat (v5.1.0) (*59*). In brief, datasets were first normalized using SCTransform (*60*). The top 3,000 integration features shared between the datasets were selected and used to identify integration anchors via canonical correlation analysis (CCA) with the 'FindIntegrationAnchors' function. The joint integrated embedding was calculated using the 'IntegrateData' function, followed by principal component analysis (PCA). The first 30 principal components were then used for UMAP visualization and Leiden clustering.

Following integration, cell identity annotation for Droplet Paired-Tag RNA data was carried out using a custom script similar to the 'TransferData' function in Seurat. At the major cell type level, we identified the 25 nearest neighbors for each query cell in the Droplet Paired-Tag dataset based on the 10x Multiome RNA modality using RANN (v2.6.2) (10.32614/CRAN.package.RANN). Distances were scaled, standardized, and converted into weights. For each query cell, the prediction score for each major cell type label was calculated by multiplying a binary classification matrix by the weights. The identity of each query cell was determined as the label with the highest prediction score. At the cell subpopulation level, a similar process was applied, except only 5 nearest neighbors were used. For downstream analysis at the major cell type level, cells with a prediction score < 0.9 were excluded. For analysis at the cell subpopulation level, cells with a prediction score < 0.8 were removed.

Preprocessing of Droplet Hi-C data

Droplet Hi-C data were processed as previously described (*13*). Fastq files were demultiplexed using the 'mkfastq' command in cellranger-atac (v2.0.0). After demultiplexing, the cellular barcode sequences were extracted, aligned to the whitelist using Bowtie (v1.3.0), and appended to the start of each read name to record the cellular identity (*61*). Potential sequencing adapters were trimmed using Trim-Galore (v0.6.10), and the reads were mapped to the reference genome (hg38) using BWA-MEM (v0.7.17) with the arguments '-SP5M' specified (*62*) (https://doi.org/10.5281/zenodo.7598955). Contact pairs were parsed, sorted, and deduplicated using Pairtools (v0.3.0) (*63*), with barcode information stored as separate columns in the pairs file.

Before downstream analysis, high-quality nuclei were selected based on the number of per-cell unique contacts in each library and then processed for donor phasing using genotype information. Nuclei confidently assigned to a single donor were retained and split into individual pairs files, with each file containing all contact information from a single nucleus. Imputation was performed using scHiCluster (v1.3.5) (*64*) for individual cells at three different resolutions: 100 kb (for compartment analysis), 25 kb (for domain analysis), and 10 kb (for cell embedding and loop analysis).

Reference mapping of Droplet Hi-C data

We adopted a previously described method to reference-map and annotate Droplet Hi-C profiles with gene expression data generated from the same tissue and samples (*65*). In brief, single-cell gene associating domain (scGAD) score Rij is calculated as the total contact number across the gene body for gene i in cell j at 10 kb resolution. The chromatin interaction profiles can then be represented as a cell-by-gene scGAD score matrix, which has been demonstrated to be correlated with gene activity. This matrix will serve as the input for reference mapping with snRNA-seq data.

To perform reference mapping, The RNA modalities from standard 10x Multiome and Droplet Paired-Tag were integrated and used. The integrated features from the reference dataset were selected and subjected to dimension reduction by PCA. The derived PCA model was then applied to transform the scGAD score matrix with the same features normalized and scaled. Subsequent integration of scGAD score with reference dataset was conducted using canonical correlation analysis. The integrated embedding was then transformed and visualized with the pre-calculated UMAP embedding model from the reference dataset.

To annotate cell identities within using scGAD score, we identified the 15 nearest neighbors in reference RNA data for each Droplet Hi-C cell via PyNNDescent (v0.5.6) (*66*) using Euclidean distance. These distances were scaled and standardized, and neighbors with the shortest distance would be assigned the highest score. Identity of each Droplet Hi-C cell was inferred by nominating the cell type label that garnered the highest standardized score among its top nearest neighbors.

<u>Predicting enhancer–gene pairs with the Activity-by-contact (ABC) model</u>

We used the ABC model (*17*) to infer putative enhancer-gene links for each condition. The ABC model took imputed contact matrices at 10 kb resolution from Hi-C data, along with H3K27ac signal over putative enhancers (in our case consensus peaks identified in the corresponding cell type from ATAC modalities). Predictions with an ABC score greater or equal to 0.02 were considered positive and retained for downstream analysis.

<u>Annotation of genome using ChromHMM</u>

We used ChromHMM (*19*) to integrate and summarize all three epigenetic modalities (ATAC, H3K27ac and H3K27me3) and annotate the genome. To generate the ChromHMM model, we first split the fragments files from cellranger atac output by cell type and disease conditions. The resulting bed files were then binarized using the 'binarizeBed' function of ChromHMM, with the `-center` flag specified. A model with states number set to 5 was trained across all conditions using the LearnModel function with default parameters. Chromatin states were manually annotated based on prior knowledge and the likelihood of epigenetic marks associated with each state. It should be noticed that the states identified here may differ from those in other studies due to the exclusion of additional histone marks.

<u>ChromVar</u>

Motif accessibility z-scores were calculated from snATAC-seq data using chromVAR (v1.22.1) (*67*). The fixed peak sparse count matrix was converted to a SummarizedExperiment object, and GC content bias was estimated using chromVAR's internal method. Human transcription factor motifs were obtained from the JASPAR2022 Bioconductor package and mapped to peaks using motifmatchr (v1.22.0). The motif annotations and SummarizedExperiment were then used as input for chromVAR's computeDeviations function to generate GC bias–corrected motif accessibility Z-scores.

<u>ChromBPNet</u>

We trained a ChromBPNet (v0.1.7) model using the default preprocessing parameters outlined in the ChromBPNet tutorial (cite 39829783). To account for Tn5 bias, we trained a separate model using our own 10x multiome pseudobulk data. Two bias-factorized models were trained on human heart Ventricular and Atrial Cardiomyocytes from non-HF individuals.

For training, we used MACS2 narrowPeak regions with a P value threshold of <0.05, called on pseudobulk ATAC fragments from all ventricular cardiomyocytes. Chromosomes 1, 3, and 6 were assigned for training, while chromosomes 8 and 20 were reserved for validation. The model was trained using default parameters. ChromBPNet models have two output heads:

1) Profile head – predicts the shape of the chromatin accessibility profile.
2) Counts head – estimates the total read counts within a profile.

For analysis, we used chrombpnet_nobias.h5 models and computed sequence contribution scores for one or both output heads. Additionally, we applied ChromBPNet for variant prediction to assess the impact of genetic variants on chromatin accessibility within our dataset.

ChromBPNet plots

Contribution scores and predicted counts for reference and alternate alleles were generated using the chromBPNet_nobias model. Alternate alleles were introduced into the reference genome using the consensus command from bcftools (v1.10.2) (*68*) to create allele-specific genome sequences. The contribs_bw and pred_bw commands from chromBPNet were then applied separately to the reference and alternate genome versions to compute contribution scores and predicted signal tracks.

motifbreakR

To identify motifs disrupted by alleles of interest, we ran motifbreakR (v2.8.0) using human transcription factors from JASPAR2022 and HOCOMOCO_v11 accessed via MotifDb (v1.36.0) (*69*).

Single cell deconvolution

Multiomics, DPT, Droplet Hi-C data were demultiplexed using demuxlet (v2) (*70*) (https://github.com/statgen/popscle). Genotype VCFs for demultiplexing were generated from TOPMed R3 imputed data, filtered for imputation quality ($R^2 > 0.9$) and minor allele frequency (>1%) using bcftools. For Droplet Paired Tag data, the filtered VCF and the DNA BAM file (from 10x Multiome or Droplet Paired-Tag, processed with CellRanger) were provided as input to demuxlet. For Droplet Hi-C, cell barcodes were appended to BAM records as "CB" tags to match 10x formatting, and BAM files were sorted prior to demultiplexing using the same filtered VCF.

Single cell multiome clustering and annotation

Single cell Multiomics samples were processed using Cellranger (v6.0.1) with the reference genome hg38. Individual samples were processed and analyzed in Seurat (v4.3) for quality control (*71*). Cells with > 500 expressed genes, > 1000 ATAC fragments, ≤ 5% mitochondrial genes, ≥ 2 TSS enrichment were kept. Ambient RNA contamination was accounted for by running SoupX (v1.6.2) (*57*) on each sample using its automated algorithm to estimate contamination rates. Gene expression count matrices were adjusted for predicted contamination and rounded to integers (adjustCounts(sc,roundToInt=TRUE)); these SoupX adjusted counts were used for both clustering and downstream analysis. Then, all samples were merged; mitochondrial genes were removed; and a first round clustering on the WNN space was performed using 5kb windows. The data was then batch-corrected using harmony (v1.2.0) (*72*) by sample, and major cell types and cell compartments were then annotated. We then further cleaned the data by calculating Silhouette scores between any combinations of cellular compartments and removed all cells that didn't pass a threshold of -0.5 (1,953 cells), and by performing one round of manual cleaning to reach a final number of cells of 329,255 cells. After cleaning, the data was re-clustered and re-harmonized by sample and gender, and peaks were called by major cell types using Signac (v1.12.0) (*73*) with default parameters. We first limited peak size for all called peaks to 300 bp by centering any peaks larger than 300 bp at their summit and extending coordinates 150 bp in either direction. We then grouped peaks based on overlap to create clusters of peaks using bedops v2.4.41 (*74*). Within each cluster, the peak with the highest read count at its summit was identified as the reference peak for the region. We then generated a list of peaks that did not overlap any of the reference peaks and began the process of clustering and identifying reference peaks again until no peaks remained. Finally, peaks were filtered using quantile thresholding (Q2) with both pileup and variability scores. For the CAREHF dataset: Single cell Multiomics samples were processed as described above. Doublets were detected and removed from each sample using Scrublet 0.2.3 (*75*) with an automatic threshold, considering an expected doublet rate of 6% for all samples.

<u>Differential analysis</u>

Differential expression analysis was performed using DESeq2 1.42.0 on pseudo-bulked RNA, ATAC, and PairedTag counts for each cell type (*23*). To account for potential confounders, gender and 10x library batch effects were included as covariates in the model. The analysis aimed to identify statistically significant differences between heart failure (HF) and non-heart failure (Non-HF) samples.

For each dataset, features were filtered to include only those with a minimum of 10 counts across all samples, ensuring sufficient statistical power and reducing noise from low-abundance features. DESeq2's default normalization and shrinkage methods were applied to control for variance, and statistical significance was assessed using Benjamini-Hochberg false discovery rate (FDR) correction to account for multiple testing.

<u>GSEA analysis</u>

Gene set enrichment analysis (GSEA) (*76*) was performed using FGSEA (v1.26.0) on differentially expressed genes identified from DESeq2 results (*77*). For each dataset, pseudo-bulk differential expression results were processed by first removing ribosomal genes and ranking genes based on their statistical score (stat column), ensuring non-mapped genes were excluded. The ranked gene lists were sorted in descending order and used as input for FGSEA, which was conducted against a curated gene set database, considering pathways containing between 10 and 500 genes. Significantly enriched pathways were identified based on false discovery rate (FDR) < 0.1. To remove redundancy, a hierarchical pathway reduction approach was applied, generating a final list of non-redundant enriched pathways ranked by Normalized Enrichment Score (NES).

<u>Motif enrichment and GSEA analysis on Target Genes</u>

Candidate cis-regulatory elements (cCREs) for each set of cell type-specific cCRE or ABC links were analyzed using HOMER (v4.11.1) with function 'findMotifsGenome.pl' (*22*), with peaks from all ABC link candidates serving as the background. Additionally, up to 500 coding target genes from each set of cell type-specific ABC links were analyzed using Gene Set Enrichment Analysis (GSEA) (MSigDB). The analysis was conducted using a false discovery rate (FDR) q-value threshold of <0.05, with gene sets ranging from 10 to 500 genes, and enrichment was evaluated against all pathways within the Canonical Pathways (CP) category. Gene ontology analysis of genes located near cell type–specific cCREs was performed using HOMER's annotatePeaks.pl script (*22*) using the `-go` flag.

<u>Defining heterogeneity within cell types</u>

For each of the 12 major cell types, further filtering and clustering analyses of the multiome cells were performed using the Seurat package. Nearest neighbor graphs were calculated utilizing the 'FindMultiModalNeighbors' function with the 1:50 components for the RNA assay, and the 2:50 components for the ATAC assay. The generated nearest neighbor graph was then used for graph-based, semi-unsupervised clustering (function 'FindClusters', resolution of 0.2) and UMAP to project the cells into two dimensions (function 'wnnUMAP'). Cell identities were assigned to the clusters by cross-referencing their marker genes with known cardiac cell type markers from both human and mouse studies, in addition to in situ hybridization data from the literature. On occasion, a cell cluster would emerge that expressed marker genes representing multiple populations, as well as containing cells with low UMI and gene counts that escaped the first filtering step. These cells were removed from downstream analyses.

<u>Subpopulation differential gene and CRE analyses</u>

Differentially expressed genes (DEGs) and differentially accessible regions (DARs) were identified using DESeq2 (v1.38.3) for RNA and ATAC, respectively. To mitigate unwanted variation, the design model accounted for gender and donor covariates. To address inflated log2 fold-change (log2FC) values, the ashr method was applied for shrinkage (*78*). We considered genes and cCREs with a fold change > 0.58 and 0, respectively, and an adjusted p-value < 0.05 after Benjamini-Hochberg correction as subpopulation specific.

To identify biological processes associated with the differential genes, GO Biological Terms were calculated with EnrichR (v3.1) using significant DEGs per subpopulation and further filtered for significant GO Terms (adjusted p-value < 0.05) (*79*).

<u>Subpopulation RNA velocity analysis</u>

RNA velocity analysis was conducted using scVelo (v0.2.5) (*80*). Spliced and unspliced read counts were computed with velocyto (v0.17.17) from raw sequencing data, using the gex-GRCh38-2020-A reference genome. The spliced and unspliced count matrices were filtered to exclude cells with fewer than 20 shared reads, retaining only the top 20000 variable genes. The data were normalized, and moments were computed using 'scvelo.pp.moments', with the top 30 PCs and 50 nearest neighbors. Dynamics were recovered using 'scv.tl.recover_dynamics'. To refine the dynamics recovery, marker genes representing HF related gene expression changes were selected. Finally, RNA velocities were calculated using scvelo.tl.velocity, and velocity graphs were constructed with 'scvelo.tl.velocity_graph' for latent time inference and visualization.

<u>Gene regulatory network analysis</u>

To identify genetic programs regulating heart failure (HF) related changes in cardiac cells, we applied the SCENIC+ (v1.0.1.dev4+ge4bdd9f) gene regulatory network (GRN) tool to each major cell type. To this end, the multiome dataset was split by cell type, and SCENIC+ analysis was performed to identify cell-subpopulation-specific regulons following the standard pipeline (https://scenicplus.readthedocs.io/en/latest/) (*24*). In brief, SCENIC+ is a three-step workflow that involves identifying candidate enhancers, identifying enriched TF-binding motifs, and linking TFs to candidate enhancers and target genes. For each major cell type, up to 10k cells were used for the GRN construction. The cCREs were set to the multiome defined peak set. To identify the top regulon for each subpopulation, regulons were filtered based on their regulatory specificity scores (RSS) and visualized for each subpopulation by degree centrality. GRNs were visualized in Cytoscape for the top regulons by RSS and direct target genes. Pseudotime trajectories per gene were calculated from scvelo results and were incorporated as the color scheme in the GRN visualizations.

Genome-wide association study enrichments

For 77 genetic traits derived from the Million Veteran Program (MVP) (*81*), CardioVascular Disease Knowledge Portal (CVDKP) (*82*), published studies (*7*, *8*), and FinnGen (*83*), we performed summary statistics munging using the LDSC 1.0 (Linkage Disequilibrium Score Regression LDSC) framework (*41*). Summary statistics were first processed with munge_sumstats.py to ensure standardized formatting and compatibility with LDSC analysis. During this step, SNP alleles were harmonized, and filtering was applied based on a minor allele frequency (MAF) threshold of 0.01. Each dataset was aligned to the HapMap3 reference panel (*84*) to retain high-confidence variants. Key parameters included the specification of effect allele (EA), non-effect allele (NEA), p-value (pvalue), effect size (beta), allele frequency (af_alt), and sample size (TotalSampleSize), with signed sumstats based on effect size directionality. The processed summary statistics were then analyzed using partitioned heritability estimation, leveraging cell type annotations to determine the contribution of specific genomic features to overall trait heritability. First, liftover was applied to convert hg38 peak coordinates to hg19 for each cell type. The resulting hg19 bed files were used to ensure compatibility with reference datasets in subsequent LDSC analyses. Next, annotation files were created for each cell type by mapping the lifted-over peak coordinates to SNPs from the 1000 Genomes Project (Phase 3, European population) (*85*). This was done using make_annot.py, generating one annotation file per chromosome for each cell type. Once annotations were generated, LD scores were computed using ldsc.py with a window size of 1 centimorgan (cM). This step involved thinning the annotation files and calculating LD scores for each SNP while ensuring overlap with baseline LD scores from the 1000 Genomes reference panel. Finally, heritability enrichment analysis was performed using ldsc.py, incorporating cell type annotations alongside baseline LD models. This step was executed for all the processed summary statistics. The analysis estimated the heritability enrichment and statistical significance of each annotation for each trait while controlling for allele frequencies and LD structure.

LDSC heritability enrichment results were aggregated across all cell types and filtered using FDR correction (0.1) to retain significant associations. Non-negative matrix factorization (NMF) (*86*) was applied to decompose the LDSC enrichment matrix into six latent factors (rank = 6), capturing shared heritability signals across cell types and traits. The analysis was run 50 times to ensure stability, generating a basis matrix (W) for cell type contributions and a coefficient matrix (H) for trait associations. Features and traits were then sorted by their dominant factor assignments to enhance interpretability. A heatmap was generated using pheatmap (cite https://cran.r-project.org/package=pheatmap) to visualize enrichment patterns, with row-wise scaling applied to highlight relative differences. At this step additional manual sorting was performed to refine biological clustering. Statistical significance was overlaid, and clustering was manually disabled to preserve the factor-based ordering. To assess the contribution of chromatin state to trait heritability, LDSC was re-run using the same parameters, but with annotations stratified into open and active chromatin states. This allowed for the evaluation of heritability enrichment specifically within functionally distinct regulatory regions. To ensure comparability across analyses, the same column and row order from the initial NMF-based visualization was retained, enabling a direct comparison of how different chromatin accessibility states contribute to genetic trait heritability across cell types.

We performed enrichment of sub-type GRNs using genome-wide association study data for dilated cardiomyopathy (DCM). We retained variants with minor allele frequency (MAF) > 0.05 and calculated Bayes Factors (BF) for each variant using the approach of Wakefield (*87*). For each cardiomyocyte sub-type, we obtained the set of cCREs in GRNs with subpopulation specific activity, and used liftOver to convert coordinates to hg19. We then tested for enrichment of sub-type GRN cCREs for DCM-associated variants using fgwas (v0.3.6) with a window size of 2,000 variants.

Fine Mapping

Credible sets were generated for a publicly available genome-wide association study (GWAS) for Dilated Cardiomyopathy (DCM) (*7*). Where available, causal variants from multi-trait genome-wide association summary statistics (MTAG) were included. First, 1 MB windows were determined from reported causal variants identified from single causal variant fine-mapping. Next, approximate Bayes factors were calculated using the Wakefield calculation (corrcoverage v1.2.0), which takes into account variant p-values and a prior for the standard deviation of the effect size (*87*). Finally, posterior probabilities were calculated using variant z-scores, effect size variances, and the prior for the standard deviation of the effect size. Credible sets with a 95% coverage were produced by determining the minimum number of variants required to reach a posterior probability sum of 0.95.

Fine Mapping Integration

For each cell type, genomic intersections between fine-mapped variants and chromatin accessibility peaks were identified using the findOverlaps function from the GenomicRanges package (*88*). Overlapping variants were extracted, and peak information was assigned to each variant overlap event. The total number of overlapping variants, independent signals, and peaks intersecting variants were computed for each cell type across different chromatin states.

Hi-C loops, ABC links, ChromBPNet predictions and differential accessibility features for each cis-regulatory element (CRE) were integrated using dplyr (*89*) join functions in R. All visualizations were generated using ggplot2 (*90*). The risk/protective allele information was annotated based on the GWAS results.

FORA from the fgsea (v1.26.0) package was used to calculate over-represented canonical pathways in vCM GRNs using all human genes as the background and the following parameters: minSize = 10, maxSize = 500 against all canonical pathways (https://www.gsea-msigdb.org/gsea/msigdb/download_file.jsp?filePath=/msigdb/release/2024.1.Hs/c4.all.v2024.1.Hs.symbols.gmt).

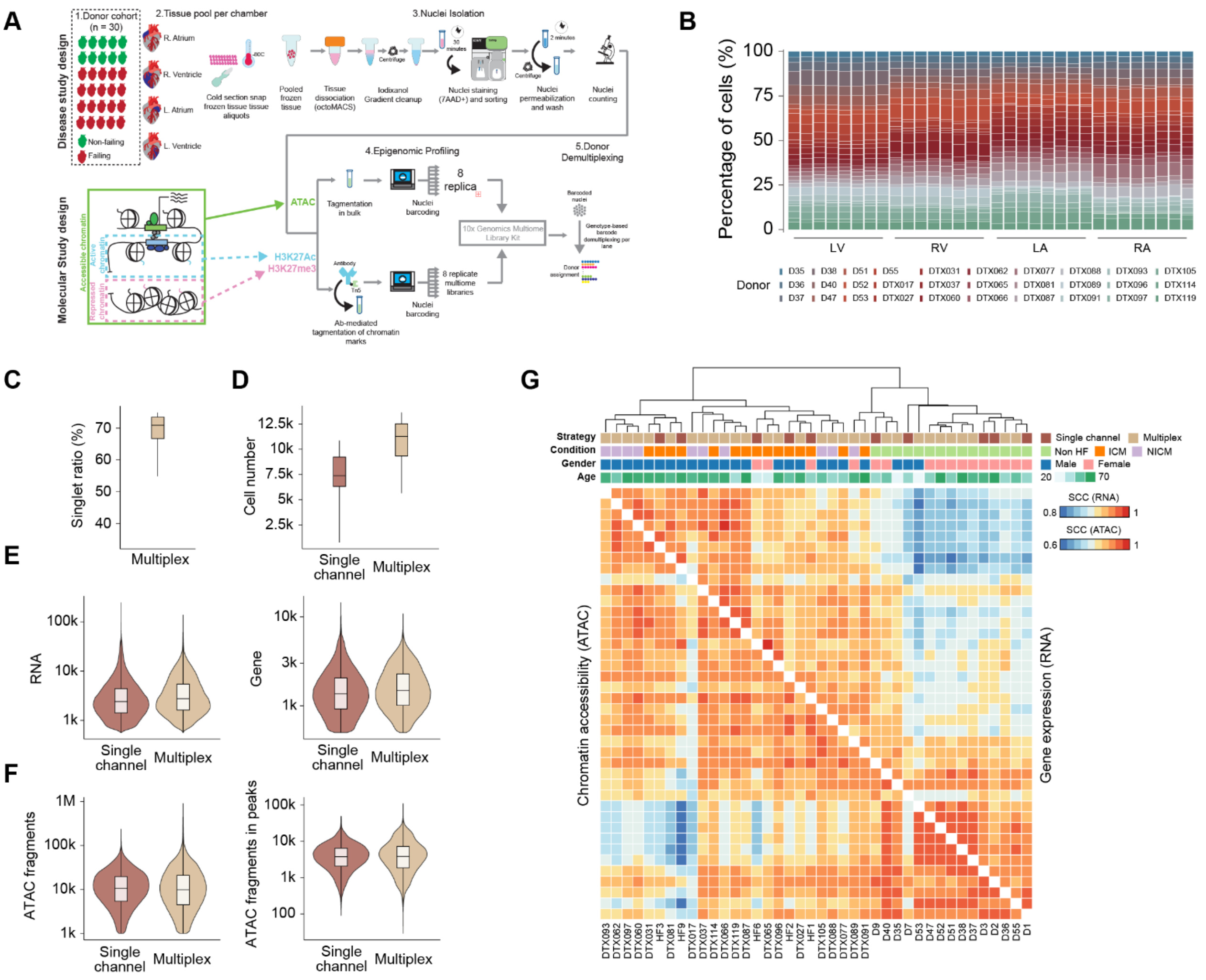


**Fig. S1. Quality control of 10x Multiome data generated using pooled sample-multiplex strategy. (A)** Schematic diagram illustrates the workflow of pooled sample-multiplex strategy. **(B)** Bar plot shows the proportion of nuclei from each donor across all pooled sample-multiplexed 10x Multiome reactions. Samples from the same cardiac chamber were pooled together. **(C)** Boxplot shows the percentage of non-doublet nuclei across all multiplexed 10x Multiome reactions. **(D)** Box plot shows the number of recovered nuclei between pooled sample-multiplexed and conventional (single channel) 10x Multiome reactions. **(E)** Violin plot shows the distribution of the UMI counts (left) and unique gene number (right) per nucleus in pooled sample-multiplexed versus single channel 10x Multiome reactions. **(F)** Violin plot shows the distribution of the unique ATAC fragment number (left) and signal enrichment level (fraction of reads in peaks, right) per nucleus in pooled sample-multiplexed versus conventional 10x Multiome reactions. **(G)** Heatmaps show Spearman's rank correlation coefficients (SCC) between gene expression profiles (top) or chromatin accessibility profiles (bottom), aggregated by donor from pooled multiplexed and single channel 10x Multiome reactions. Hierarchical clustering demonstrates that donors are grouped primarily by heart failure condition rather than by gender or multiplexing strategies.

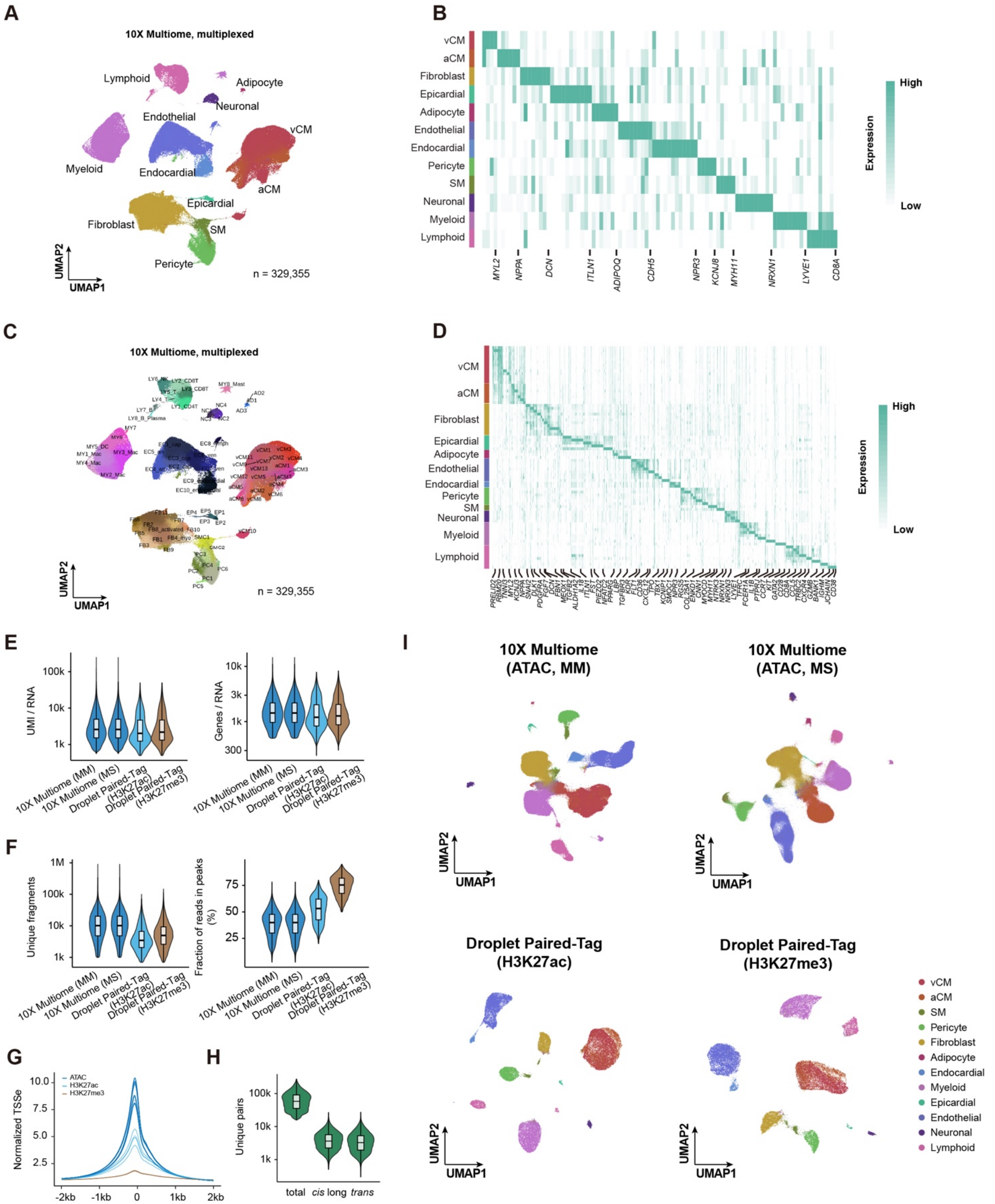
A
10X Multiome, multiplexed
Lymphoid
Adipocyte
Neuronal
Endothelial
vCM
Myeloid
Endocardial
aCM
Epicardial
Fibroblast
SM
Pericyte
UMAP2
UMAP1
n = 329,355
B
vCM
aCM
Fibroblast
Epicardial
Adipocyte
Endothelial
Endocardial
Pericyte
SM
Neuronal
Myeloid
Lymphoid
MYL2
NPPA
DCN
ITLN1
ADIPOQ
CDH5
NPR3
KCNJ8
MYH11
NRXN1
LYVE1
CD8A
High
Expression
Low
C
10X Multiome, multiplexed
UMAP2
UMAP1
n = 329,355
D
vCM
aCM
Fibroblast
Epicardial
Adipocyte
Endothelial
Endocardial
Pericyte
SM
Neuronal
Myeloid
Lymphoid
High
Expression
Low
E
UMI / RNA
Genes / RNA
10X Multiome (MM)
10X Multiome (MS)
Droplet Paired-Tag (H3K27ac)
Droplet Paired-Tag (H3K27me3)
F
Unique fragments
Fraction of reads in peaks (%)
G
Normalized TSSe
ATAC
H3K27ac
H3K27me3
-2kb
-1kb
0
1kb
2kb
H
Unique pairs
total
cis long
trans
I
10X Multiome (ATAC, MM)
10X Multiome (ATAC, MS)
Droplet Paired-Tag (H3K27ac)
Droplet Paired-Tag (H3K27me3)
UMAP2
UMAP1
vCM
aCM
SM
Pericyte
Fibroblast
Adipocyte
Endocardial
Myeloid
Epicardial
Endothelial
Neuronal
Lymphoid

**Fig. S2. Quality control of different single-cell profiles generated from human cardiac tissue**. **(A)** Uniform manifold approximation and projection (UMAP) embedding visualizes sample pooled-multiplexed 10x Multiome data, clustered and annotated based on transcriptomic profiles. Nuclei are colored and labeled by major cell types. **(B)** Heatmap shows scaled expression of canonical marker genes for each major cell type. **(C)** UMAP embedding from (A) but colored and labeled by cell subpopulation. **(D)** Heatmap shows scaled expression of canonical marker genes for each cell subpopulation. **(E)** Violin plot shows the distribution of the UMI counts (left) and unique gene number (right) per nucleus across different multiomic methods, including pooled sample-multiplexed 10x Multiome (MM), conventional 10x Multiome (MS) and Droplet Paired-Tag (targeting H3K27ac or H3K27me3). **(F)** Violin plot shows the distribution of the unique ATAC fragment counts (left) and signal enrichment level (fraction of reads in peaks, right) per nucleus across different multiomic methods. **(G)** Line graph shows the normalized signal enrichment at transcription start site (TSS) for each sequencing library across different epigenetic marks including ATAC, H3K27ac and H3K27me3. **(H)** Violin plot shows the distribution of total, *cis*-long or *trans* unique contact pairs per nucleus from Droplet Hi-C data. **(I)** UMAP embedding of different epigenetic modalities from multi-omic methods, including chromatin accessibility from sample-multiplexed 10x Multiome (MM) and conventional 10x Multiome (MS), H3K27ac and H3K27me3 from Droplet Paired-Tag. Nuclei are colored and labeled by major cell types identified from the corresponding paired single-nucleus transcriptomic data.

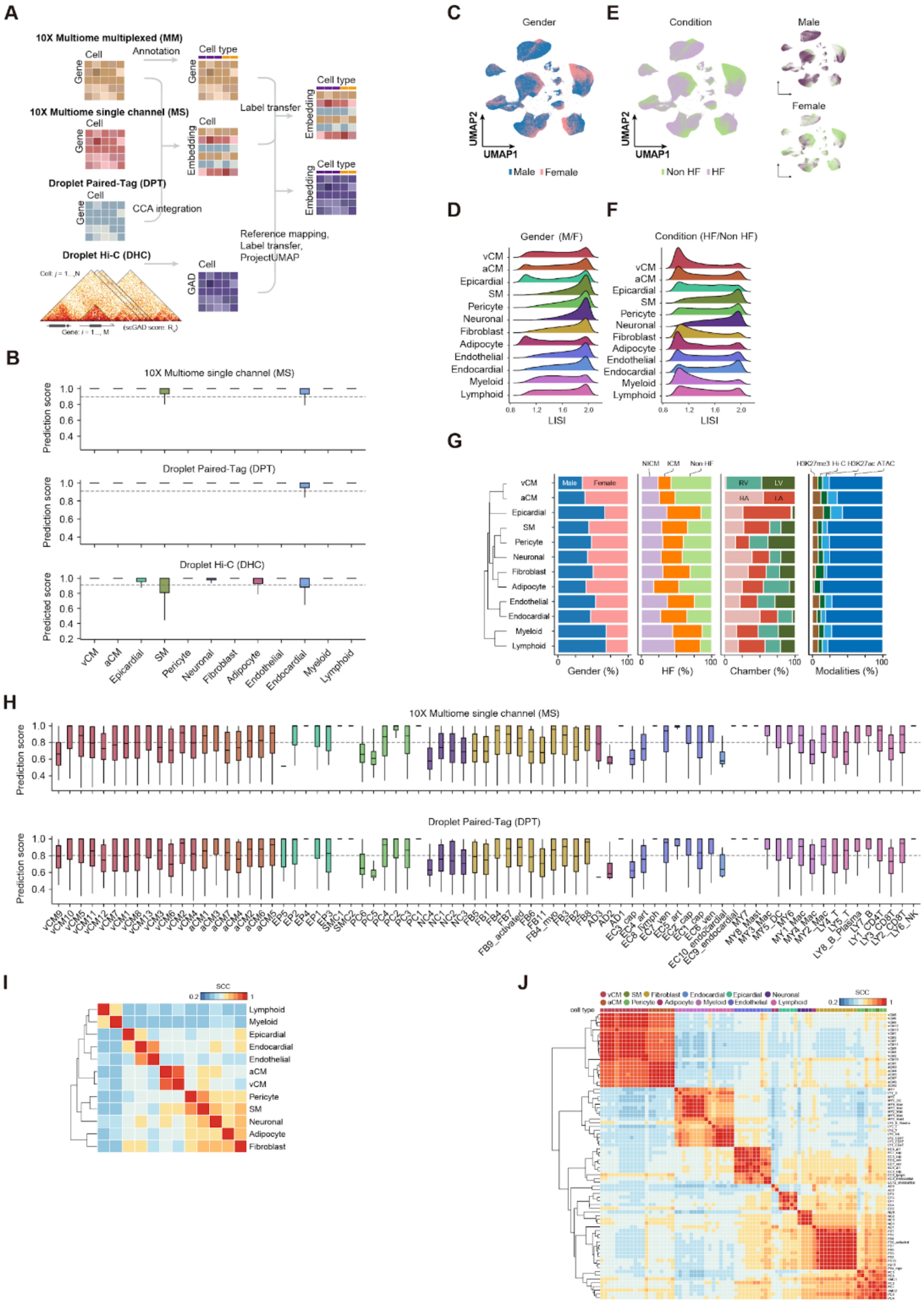
A
10X Multiome multiplexed (MM)
Annotation
Cell type
Cell
Gene
10X Multiome single channel (MS)
Label transfer
Embedding
Droplet Paired-Tag (DPT)
CCA integration
Reference mapping,
Label transfer,
ProjectUMAP
Droplet Hi-C (DHC)
GAD
B
10X Multiome single channel (MS)
Prediction score
Droplet Paired-Tag (DPT)
Droplet Hi-C (DHC)
Predicted score
vCM
aCM
Epicardial
SM
Pericyte
Neuronal
Fibroblast
Adipocyte
Endothelial
Endocardial
Myeloid
Lymphoid
C
Gender
UMAP2
UMAP1
Male
Female
E
Condition
Non HF
HF
D
Gender (M/F)
LISI
F
Condition (HF/Non HF)
G
NICM
ICM
Non HF
RV
LV
RA
LA
H3K27me3
Hi C
H3K27ac
ATAC
Gender (%)
HF (%)
Chamber (%)
Modalities (%)
H
10X Multiome single channel (MS)
Droplet Paired-Tag (DPT)
I
SCC
J
cell type

**Fig. S3. Identifying cardiac cell types from different single-cell profiles by integrative analysis. (A)** Schematic diagram illustrates the integrative analysis workflows across different single-cell datasets. **(B)** Boxplots show the prediction score for individual nuclei in different single-cell datasets, using major cell type labels from sample pooled-multiplexed 10x Multiome data as reference. The dash line at 0.9 indicates the cutoff used to filter out low-confidence predictions. Prediction scores were obtained from Seurat "LabelTransfer" method; Predicted scores were calculated using in-house nearest-neighbor-based cell type prediction approach. **(C)** Integrative UMAP embedding visualizes all valid nuclei, colored and labeled by donor gender. **(D)** Distribution of the diversity score (local inverse Simpson's index, LISI) by gender for each major cell type. **(E)** Same UMAP embedding as (C) but colored and labeled by heart failure (HF) condition. UMAP embeddings split by gender (male or female) are shown to demonstrate that the separation of nuclei by HF conditions is unlikely to be confounded by gender. **(F)** Distribution of the diversity score (LISI) by heart failure conditions for each major cell type. **(G)** Hierarchical organization of major cardiac cell types based on transcriptome profiles (left). Bar charts showing the proportion of nuclei by variables including gender, HF conditions, cardiac chambers and epigenetic modalities for each major cardiac cell type (right). **(H)** Boxplots show the prediction score for individual nuclei in conventional 10x Multiome (top) and Droplet Paired-Tag (bottom) dataset using cell subpopulation labels from pooled sample-multiplexed 10x Multiome data as reference. Dash line at 0.8 indicates cutoff used to filter out low-confidence predictions. (**I**) Heatmap shows Spearman's rank correlation coefficients (SCC) between gene expression profiles of major cell types. (**J**) Heatmap shows SCC between gene expression profiles of cell subpopulations.

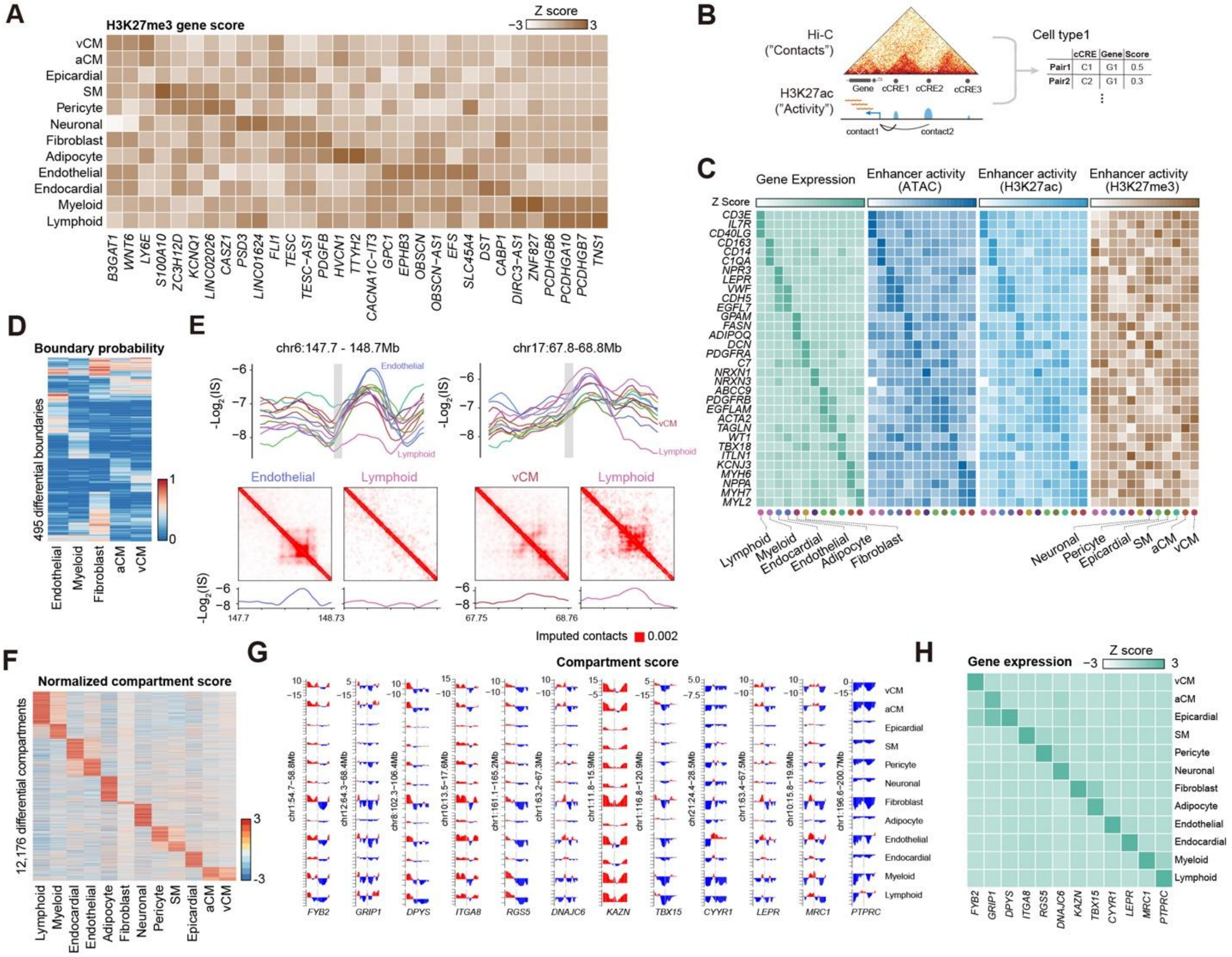


**Fig. S4. Characterization of epigenetic heterogeneity in major cardiac cell types. (A)** Heatmap shows selected genes with scaled H3K27me3 gene score across major cell types. Gene scores were calculated based on epigenetic signals over the promoter region (2 kb upstream the transcription start site) and gene body for a given gene. **(B)** Schematic diagram illustrates the calculation of Activity-by-contact (ABC) score using H3K27ac and Hi-C modalities. **(C)** Heatmaps show, for each major cardiac cell type, the scaled gene expression and enhancer activity of marker genes. Enhancer activity was calculated as the averaged epigenetic signal over putative enhancers (cCREs) identified from the ABC model. **(D)** Heatmap shows boundary probability for differential boundaries identified across selected major cell types. Probabilities were calculated across donors. **(E)** Log2 insulation score profiles are shown for major cell types over representative genomic windows (Top). Imputed contact maps and corresponding log2 insulation score profiles from representative cell types are displayed in the same genomic window (Bottom). Insulation scores were calculated on imputed contact maps. **(F)** Heatmap shows normalized compartment scores at differential compartments across major cell types. Normalized compartment scores were calculated from aggregated imputed contact maps and normalized by *dchic*. **(G)** Compartment scores are shown across genomic windows for representative marker genes in each major cell type. The gray shading indicates the position of the 100 kb bin overlapping the selected gene. **(H)** Heatmap shows the scale gene expression of selected marker genes from (G) across all major cell types.

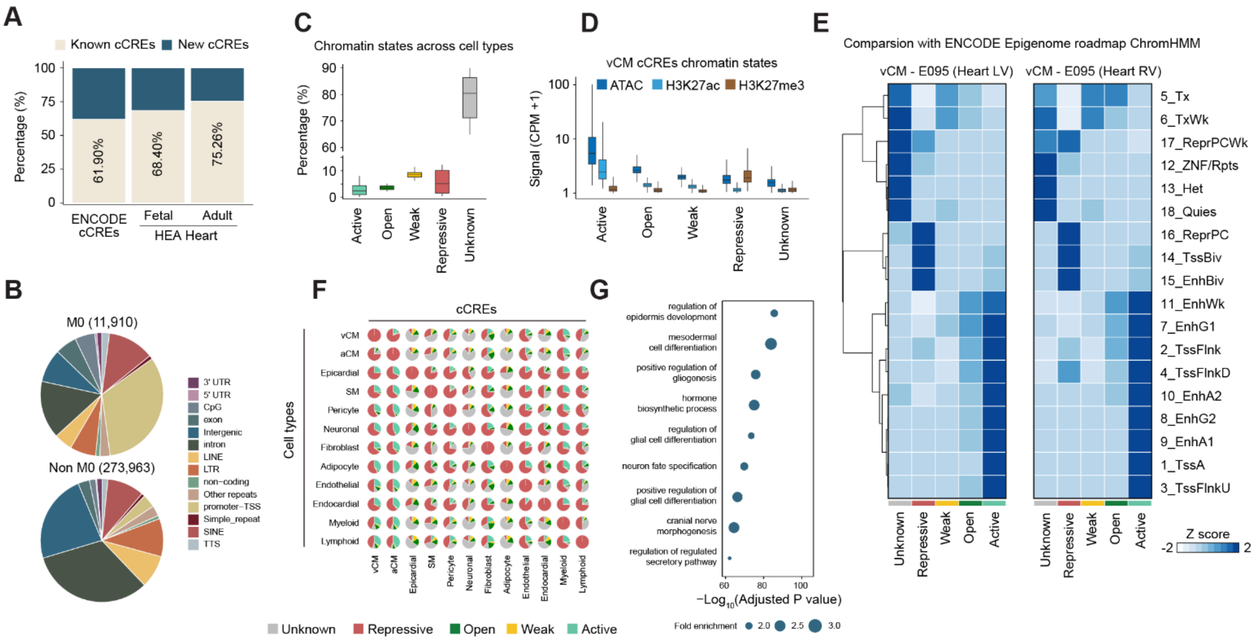


**Fig. S5. Characterization of cCREs and chromatin states across cardiac cell types. (A)** Bar charts show the percentage of cCREs overlapping with publicly available datasets, including cCREs from ENCODE SCREEN database and cCREs from adult and fetal human heart enhancer atlases. **(B)** Pie charts show the fraction of cCREs that overlap with different classes of annotated sequences in module M0 compared to other modules. **(C)** Boxplots show the percentage of genome annotated for each chromatin state across all major cell types. **(D)** Boxplots show the signal of epigenetic marks over cCREs for each chromatin state in vCMs. **(E)** Heatmaps show the scaled fraction of the chromatin state annotations in vCMs from our current 5-state model overlapping with the 18-state model annotation from the ENCODE Epigenome Roadmap project. **(F)** Pie charts show the proportion of cCREs annotated as repressive in each major cell type, along with the chromatin states of these repressive cCREs in other cell types. **(G)** Dot plot shows examples of gene ontology enrichment results from GREAT analysis of the vCM repressive cCREs. P-values were calculated using hypergeometric test in enrichGO.

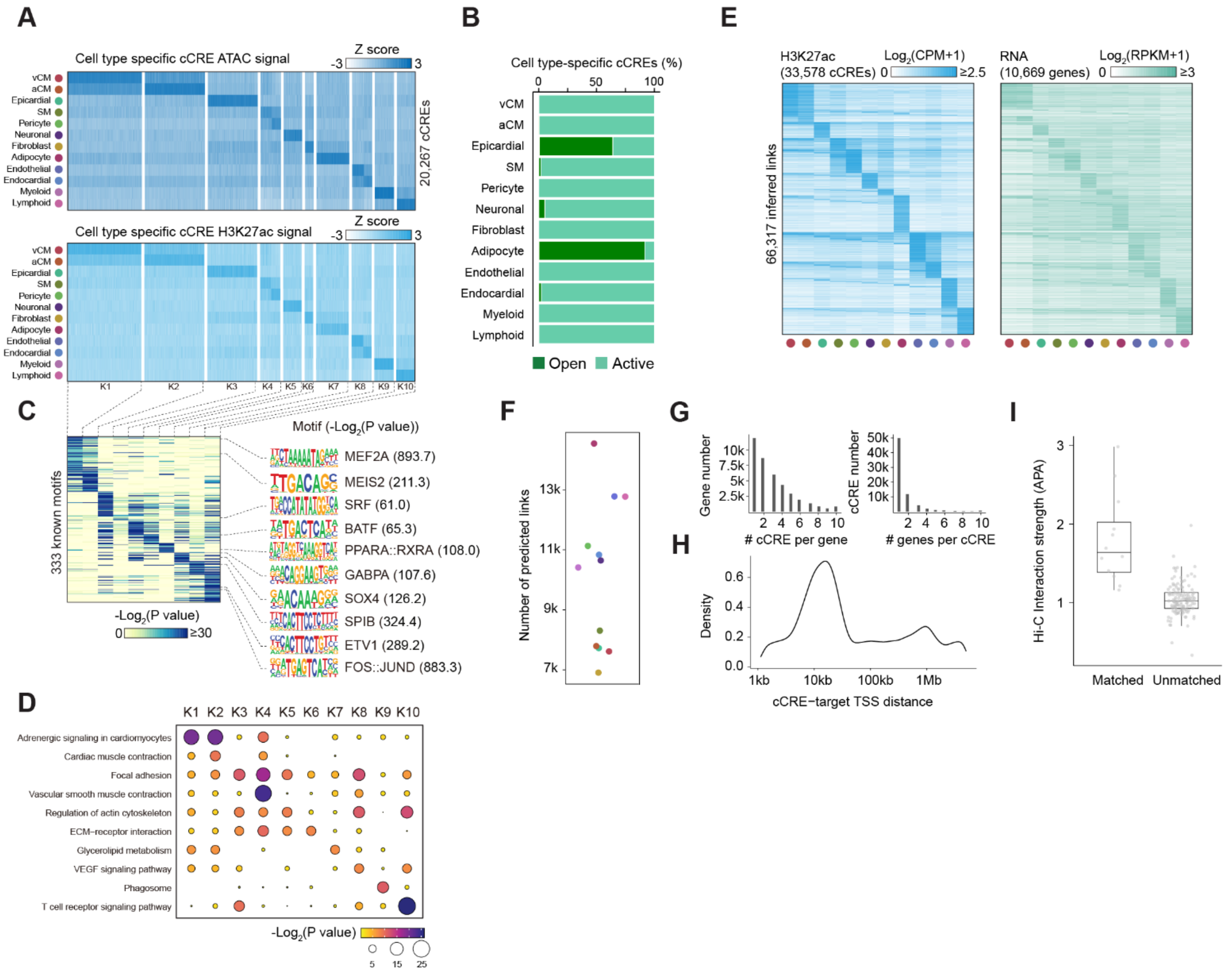


**Fig. S6. Cell type-specific cCREs and cCRE-gene links. (A)** Heatmaps show scaled chromatin accessibility and H3K27ac signal of 20,267 cell type-specific cCREs. **(B)** Bar charts show the fraction of cell type-specific cCREs classified by chromatin state. **(C)** A total of 333 known transcription factor (TF) motifs were found enriched across different cell type-specific cCREs modules. Examples of enriched motifs along with their logos and significance level are shown. P-values were calculated using binomial test by HOMER. **(D)** Gene ontology enrichment results from HOMER are shown for different cell type-specific cCRE modules. P-values were calculated using binomial test by HOMER. **(E)** Heatmaps show the scaled H3K27ac signal (left) and cognate target gene expression level (right) for 66,310 putative cCRE-gene links identified by the ABC model across all cell types. **(F)** Scatter plot shows the number of putative cCRE-gene links identified in each major cell type. **(G)** Bar graphs show the distribution of the number of cCREs targeting per gene (left) and the number of genes targeted per cCRE (right). **(H)** Distribution of genomic distance between cCREs and their putative target genes. **(I)** Boxplots show the aggregated peak analysis (APA) score, representing the averaged chromatin interaction strength at cell type-specific cCRE-gene links in corresponding cell types (matched) versus in other cell types (unmatched).

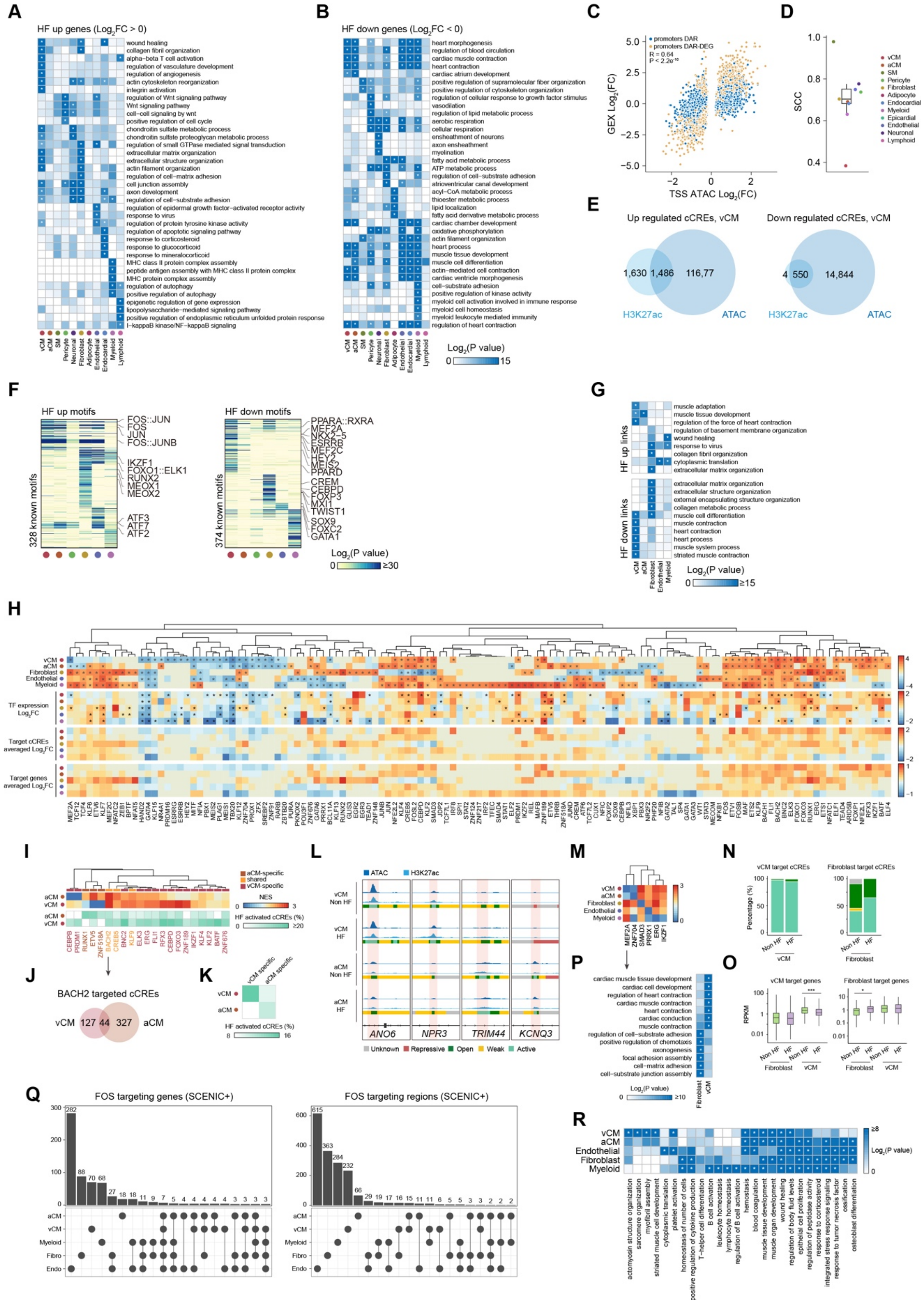
A
HF up genes (Log2FC > 0)
wound healing
collagen fibril organization
alpha-beta T cell activation
regulation of vasculature development
regulation of angiogenesis
actin cytoskeleton reorganization
integrin activation
regulation of Wnt signaling pathway
Wnt signaling pathway
cell-cell signaling by wnt
positive regulation of cell cycle
chondroitin sulfate metabolic process
chondroitin sulfate proteoglycan metabolic process
regulation of small GTPase mediated signal transduction
extracellular matrix organization
extracellular structure organization
actin filament organization
regulation of cell-matrix adhesion
cell junction assembly
axon development
regulation of cell-substrate adhesion
regulation of epidermal growth factor-activated receptor activity
response to virus
regulation of protein tyrosine kinase activity
regulation of apoptotic signaling pathway
response to corticosteroid
response to glucocorticoid
response to mineralocorticoid
MHC class II protein complex assembly
peptide antigen assembly with MHC class II protein complex
MHC protein complex assembly
regulation of autophagy
positive regulation of autophagy
epigenetic regulation of gene expression
lipopolysaccharide-mediated signaling pathway
positive regulation of endoplasmic reticulum unfolded protein response
I-kappaB kinase/NF-kappaB signaling
vCM aCM SM Pericyte Neuronal Fibroblast Adipocyte Endothelial Endocardial Myeloid Lymphoid
B
HF down genes (Log2FC < 0)
heart morphogenesis
regulation of blood circulation
cardiac muscle contraction
heart contraction
cardiac atrium development
positive regulation of supramolecular fiber organization
positive regulation of cytoskeleton organization
regulation of cellular response to growth factor stimulus
vasodilation
regulation of lipid metabolic process
aerobic respiration
ensheathment of neurons
axon ensheathment
myelination
fatty acid metabolic process
ATP metabolic process
regulation of cell-substrate adhesion
atrioventricular canal development
acyl-CoA metabolic process
thioester metabolic process
lipid localization
fatty acid derivative metabolic process
cardiac chamber development
oxidative phosphorylation
actin filament organization
heart process
muscle tissue development
muscle cell differentiation
actin-mediated cell contraction
cardiac ventricle morphogenesis
cell-substrate adhesion
positive regulation of kinase activity
myeloid cell activation involved in immune response
myeloid cell homeostasis
myeloid leukocyte mediated immunity
regulation of heart contraction
vCM aCM SM Pericyte Neuronal Fibroblast Adipocyte Endothelial Endocardial Myeloid Lymphoid
Log2(P value)
0 15
C
promoters DAR
promoters DAR-DEG
R = 0.64
P < 2.2e-16
GEX Log2(FC)
TSS ATAC Log2(FC)
D
SCC
vCM
aCM
SM
Pericyte
Fibroblast
Adipocyte
Endocardial
Myeloid
Epicardial
Endothelial
Neuronal
Lymphoid
E
Up regulated cCREs, vCM
1,630 1,486 116,77
H3K27ac ATAC
Down regulated cCREs, vCM
4 550 14,844
H3K27ac ATAC
F
HF up motifs
328 known motifs
FOS::JUN
FOS
JUN
FOS::JUNB
IKZF1
FOXO1::ELK1
RUNX2
MEOX1
MEOX2
ATF3
ATF7
ATF2
HF down motifs
374 known motifs
PPARA::RXRA
MEF2A
NKX2-5
ESRRB
MEF2C
HEY2
MEIS2
PPARD
CREM
CEBPD
FOXP3
MXI1
TWIST1
SOX9
FOXC2
GATA1
Log2(P value)
0 ≥30
G
HF up links
muscle adaptation
muscle tissue development
regulation of the force of heart contraction
regulation of basement membrane organization
wound healing
response to virus
collagen fibril organization
cytoplasmic translation
extracellular matrix organization
HF down links
extracellular matrix organization
extracellular structure organization
external encapsulating structure organization
collagen metabolic process
muscle cell differentiation
muscle contraction
heart contraction
heart process
muscle system process
striated muscle contraction
vCM aCM Fibroblast Endothelial Myeloid
Log2(P value)
0 ≥15
H
vCM
aCM
Fibroblast
Endothelial
Myeloid
TF expression Log2FC
Target cCREs averaged Log2FC
Target genes averaged Log2FC
I
aCM-specific
shared
vCM-specific
NES
0 3
HF activated cCREs (%)
0 ≥20
CEBPB PRDM1 RUNX1 ETV6 ZNF518A BACH2 CREB5 KLF9 ELK3 ERG FLI1 RFX3 CEBPD FOXO3 ZNF189 IKZF1 KLF4 KLF2 BATF ZNF676
J
BACH2 targeted cCREs
vCM 127 44 327 aCM
K
vCM specific
aCM specific
HF activated cCREs (%)
8 16
L
ATAC
H3K27ac
vCM Non HF
vCM HF
aCM Non HF
aCM HF
ANO6
NPR3
TRIM44
KCNQ3
Unknown
Repressive
Open
Weak
Active
M
vCM
aCM
Fibroblast
Endothelial
Myeloid
MEF2A ZNF704 SMAD3 PRRX1 ERG IKZF1
N
vCM target cCREs
Fibroblast target cCREs
Percentage (%)
Non HF HF
vCM
Non HF HF
Fibroblast
P
cardiac muscle tissue development
cardiac cell development
regulation of heart contraction
cardiac muscle contraction
heart contraction
cardiac conduction
muscle contraction
regulation of cell-substrate adhesion
positive regulation of chemotaxis
axonogenesis
focal adhesion assembly
cell-matrix adhesion
cell-substrate junction assembly
Log2(P value)
0 ≥10
Fibroblast
vCM
O
vCM target genes
Fibroblast target genes
RPKM
Non HF HF Non HF HF
Fibroblast vCM
Q
FOS targeting genes (SCENIC+)
282 88 70 68 27 18 18 11 9 7 5 4 4 4 4 3 3 3 3
FOS targeting regions (SCENIC+)
615 363 284 232 66 29 19 17 16 15 11 11 6 5 5 3 3 2 2 2
aCM
vCM
Myeloid
Fibro
Endo
R
vCM
aCM
Endothelial
Fibroblast
Myeloid
Log2(P value)
0 ≥8
actomyosin structure organization
sarcomere organization
myofibril assembly
striated muscle cell development
cytoplasmic translation
platelet activation
homeostasis of number of cells
positive regulation of cytokine production
T-helper cell differentiation
B cell activation
leukocyte homeostasis
lymphocyte homeostasis
regulation of B cell activation
hemostasis
blood coagulation
muscle tissue development
muscle organ development
wound healing
regulation of body fluid levels
epithelial cell proliferation
regulation of peptidase activity
response to corticosteroid
integrated stress response signaling
response to tumor necrosis factor
ossification
osteoblast differentiation

**Fig. S7. Cell type specific epigenetic changes and gene regulatory networks in heart failure.** **(A-B)** Heatmaps show enrichment of gene ontology terms for differentially up-regulated **(A)** or down-regulated **(B)** genes during heart failure in each cell type. Asterisk indicates significant enrichment (adjusted P-value < 0.05). P-values were calculated using hypergeometric test in enrichGO and corrected for multiple testing using Benjamini-Hochberg FDR correction. **(C)** Scatter plot shows the log2 fold changes of promoters classified as differentially accessible regions (DAR) versus log2 fold changes of differential gene expression (DEG) for associated genes between heart failure (HF) versus non-heart failure (Non-HF) samples. Each point is colored according to whether the associated genes are classified as differentially expressed. P-value is calculated by t-test. **(D)** Boxplot shows the Spearman's rank correlation coefficients (SCC) between the log2 fold changes of differentially accessible promoter peaks and the log2 fold changes of their associated genes across cell types. **(E)** Venn diagrams show the overlap of differentially accessible cCREs classified as up-regulated or down-regulated based on ATAC or H3K27ac signals in ventricular cardiomyocytes (vCMs). **(F)** A total of 328 and 374 known TF motifs are enriched in heart failure (HF) up-regulated (left) and down-regulated (right) cCREs, respectively, across selected cell types. Example motifs with activity changes in specific cell types are labeled. **(G)** Heatmap shows enrichment of gene ontology terms for genes within differential cCRE-gene links across selected cell types. Asterisk indicates significant enrichment (adjusted P-value < 0.05). P-values and FDR correction procedure is the same as described in (A). **(H)** Top heatmap shows heart failure-associated transcription factor (TF) gene regulatory networks (GRNs) across selected cell types. Only GRNs with absolute fGSEA normalized enrichment scores (NES) $\geq 2$ in at least one cell type are shown. Asterisk indicates significant enrichment (P-value < 0.05). P-values were estimated based on an adaptive multi-level split Monte-Carlo scheme in *fgsea* package. Middle heatmap shows log2 fold changes of transcription factors in each cell type. Asterisk indicates significant enrichment (adjusted P-value < 0.05). P-values were calculated using Wald test in *DESeq2* package and corrected for multiple testing using Benjamini-Hochberg FDR correction. Bottom two heatmaps show the averaged log2 fold changes of all cCREs and genes by each TF GRN across selected cell types. **(I)** Top heatmap shows heart failure-associated TF GRNs in atrial (aCM) and ventricular (vCM) cardiomyocytes. GRNs were classified as aCM-specific, vCM-specific, or shared based on overlap between cell types. Bottom panel shows the percentage of cCREs activated under heart failure conditions in each GRN, based on chromatin state annotations. The activation trend aligns with GRN classification. **(J)** Venn diagrams show the overlap between BACH2-targeted cCREs between vCM and aCM. **(K)** Heatmap shows the percentage of non-overlapping BACH2-targeted cCREs that are activated under heart failure conditions in vCM and aCM. **(L)** Representative genome browser tracks show vCM-specific and aCM-specific BACH2 binding sites, highlighted by shaded regions. **(M)** Heatmap shows bi-directional heart failure-associated TF GRNs across selected cell types. **(N)** Bar charts show the distribution of chromatin states for MEF2A-targeted cCREs in vCM and fibroblasts across conditions. **(O)** Boxplots show expression levels of MEF2A-targeted genes in vCM and fibroblasts across HF conditions. Asterisk indicates significance (P-value < 0.05, Wilcoxon rank-sum test). **(P)** Gene ontology enrichment results for MEF2A-targeted genes in vCM and fibroblasts are shown. Asterisk indicates significant enrichment (adjusted P-value < 0.05). P-values and FDR correction procedure is the same as described in (A). **(Q)** Upset plot shows the overlap of FOS-targeted genes and regions (cCREs) across selected cell types. **(R)** Gene ontology enrichment results for FOS-targeted genes across five major cell types. Asterisk indicates significant enrichment (adjusted P-value < 0.05). P-values and FDR correction procedure is the same as described in (A).

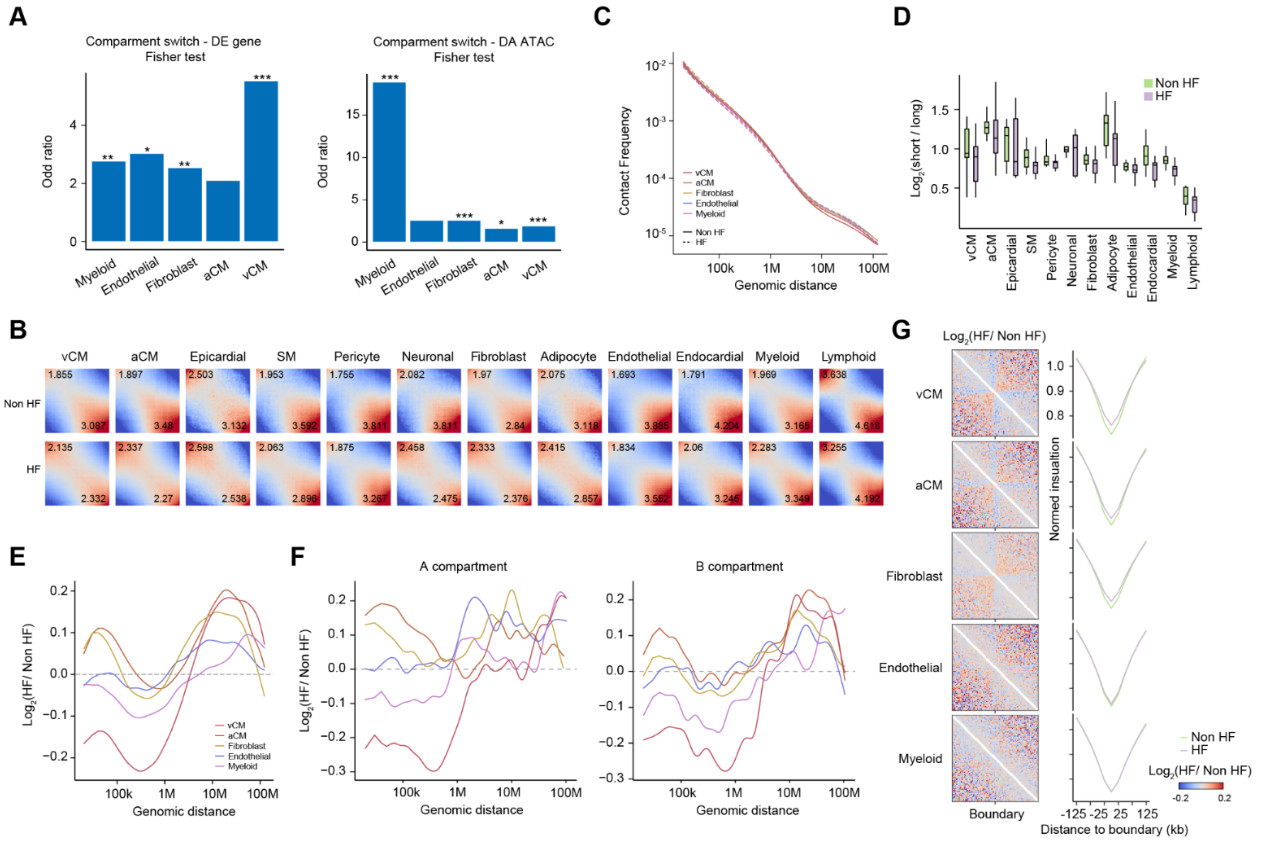


**Fig. S8. Cell type-specific changes in chromatin organization during heart failure.**
**(A)** Bar charts show the enrichment of differentially expressed (DE) genes (left) or differentially accessible (DA) ATAC cCREs (right) within switched chromatin compartments across selected cell types. Asterisk indicates significant enrichment (*, $P < 0.05$; **, $P < 0.005$; ***, $P < 0.0005$). P-value is calculated by Fisher's exact test. **(B)** Saddle plots show compartment strengths during heart failure across all cell types. Number indicates B-B compartment interaction strength (top-left) and A-A interaction strength (bottom-right). **(C)** Comparison of contact frequency by genomic distance for selected cell types in heart failure (HF) versus non-HF (control). Only chromatin interaction profiles from donors ≤ 60 years old are included for analysis. **(D)** Boxplots show the distribution of log2 ratio of long-range (20Mb-50Mb) to short-range (200kb-2Mb) interactions per donor between HF and non-HF across different cell types. **(E, F)** Log2 ratio of contact frequency by genomic distance in HF versus non-HF samples, shown for all compartments (**E**) or stratified by specific compartments (**F**) across selected cell types. **(G)** Log2 ratio of contact pileup in HF versus non-HF surrounding domain boundaries identified from pseudobulk profiles (left). Normalized insulation score in HF versus non-HF for selected cell types at domain boundaries are also shown (right).

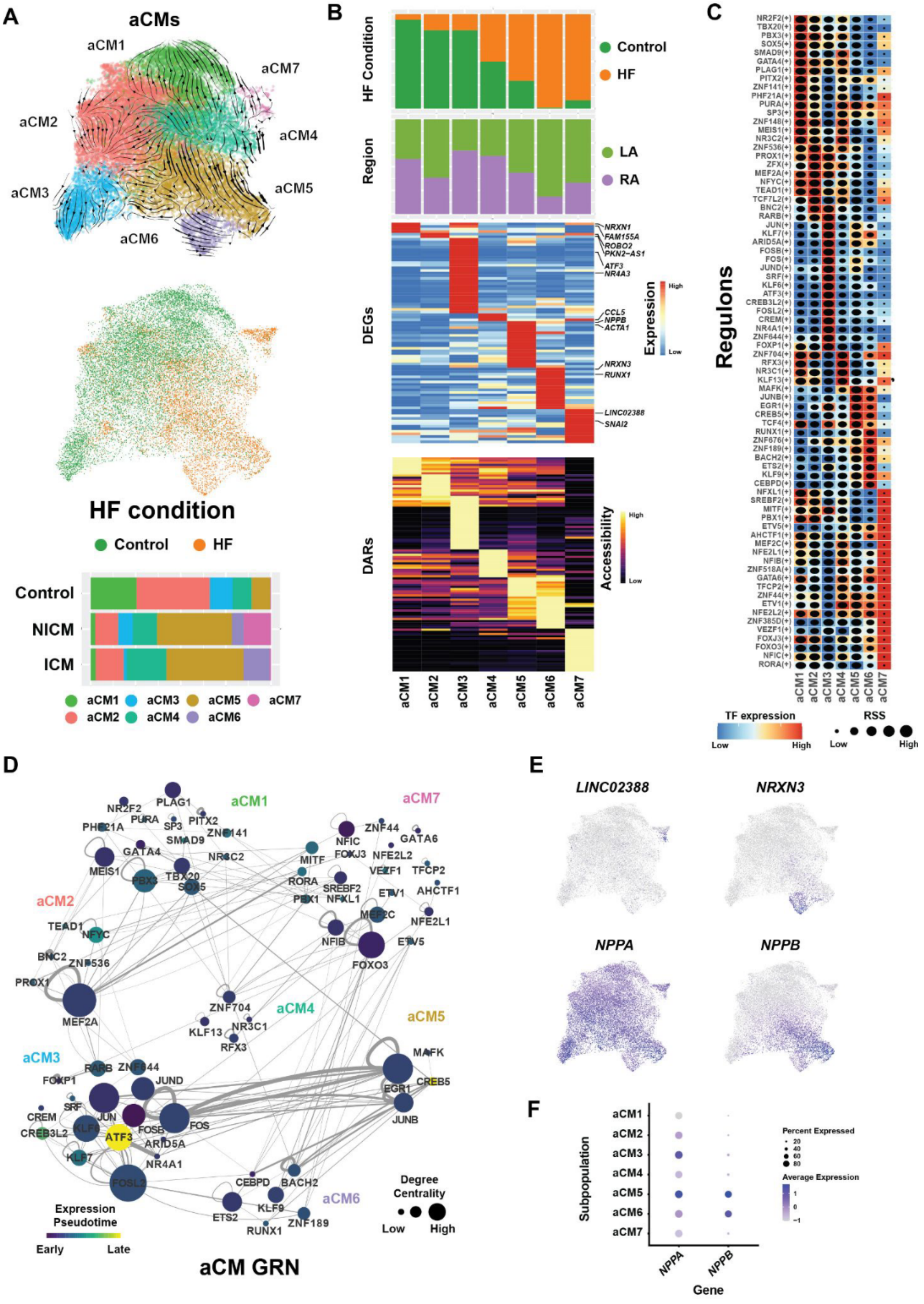
A
aCMs
aCM1
aCM2
aCM3
aCM4
aCM5
aCM6
aCM7
HF condition
Control
HF
Control
NICM
ICM
B
HF Condition
Control
HF
Region
LA
RA
DEGs
NRXN1
FAM155A
ROBO2
PKN2-AS1
ATF3
NR4A3
CCL5
NPPB
ACTA1
NRXN3
RUNX1
LINC02388
SNAI2
Expression
High
Low
DARs
Accessibility
High
Low
C
Regulons
TF expression
Low
High
RSS
Low
High
D
aCM GRN
Expression Pseudotime
Early
Late
Degree Centrality
Low
High
E
LINC02388
NRXN3
NPPA
NPPB
F
Subpopulation
Percent Expressed
20
40
60
80
Average Expression
1
0
−1
NPPA
NPPB
Gene

**Fig. S9. Gene regulatory network analysis of atrial cardiomyocytes reveals genetic programs directing their cellular states.** (**A**) UMAP embedding shows atrial cardiomyocyte (aCM) subpopulations and their RNA velocity-derived trajectories (top). UMAP embedding of aCMs labeled by heart failure (HF) condition reveals the contribution of non-HF and HF aCMs to each subpopulation (middle). Bar graph shows the aCM subpopulation composition for each cardiac condition (bottom). (**B**) Each aCM cell subpopulation exhibits distinct cellular contributions from each heart condition and cardiac chamber (top, bar graph) as well as gene expression (middle, heatmap) and chromatin accessibility profiles (bottom, heatmap). (**C**) Heatmap shows SCENIC+ predicted regulons for each aCM cell subpopulation. Box color indicates transcription factor expression, and the dot size represents the regulon specificity score (RSS). (**D**) Analyzing the TF network within the aCM GRN reveals the relationship among distinct regulons across aCM non-HF and HF subpopulations. Nodes are colored by pseudotime of expression. Node size is determined by their degree of centrality within the network, and edges represent TF-TF connections. (**E**) UMAP embeddings show the expression pattern of *LINC02388*, *NRXN3*, *NPPA*, and *NPPB* across aCMs. (**F**) Dot plot displays the expression level of *NPPA* and *NPPB* across the aCM subpopulations. LA, left atrium; RA, right atrium; ICM, ischemic cardiomyopathy; NICM, non-ischemic cardiomyopathy.

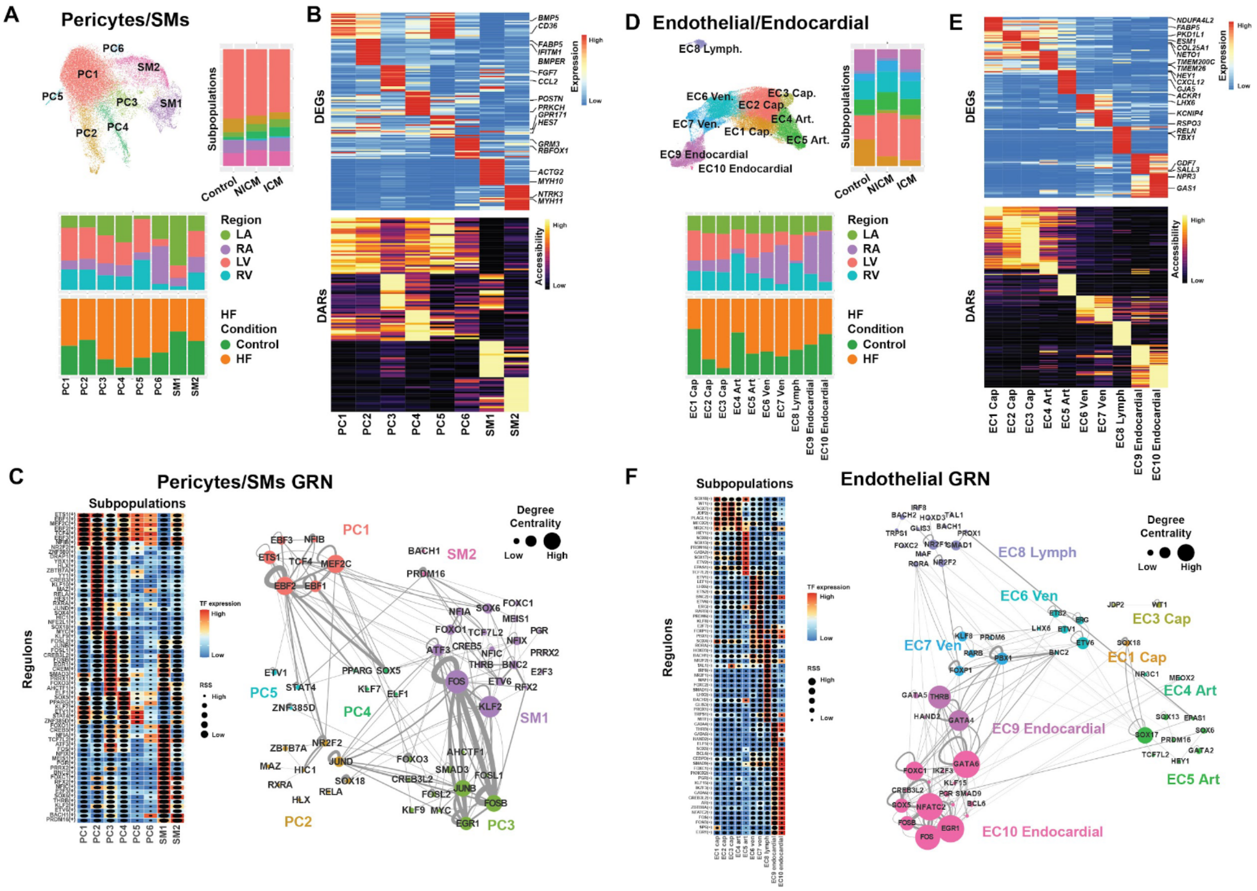


**Fig. S10. Gene regulatory network analysis of cardiac vascular cells reveals genetic programs directing their cellular states.** (**A, D**) UMAP embeddings show pericyte/smooth muscle cells (SM) (A), and endothelial/endocardial (D) cell (EC) subpopulations (top, left), respectively. Bar graphs to the right of UMAPs show the cell subpopulation composition for each cardiac condition. Bar graphs below UMAPs reveal the contribution of each cardiac chamber (top) and heart failure conditions (non-HF versus HF) to specific cell subpopulations (bottom). (**B, E**) Each pericyte/SM (B) and endothelial/endocardial (E) cell subpopulation displays distinct gene expression (top heatmap) and chromatin accessibility profiles (bottom heatmap). (**C, F**) SCENIC+ predicted regulons are shown for pericyte/SM (C) and endothelial/endocardial (F) cell subpopulations (left). Box color represents transcription factor expression, and dot size reflects the regulon specificity score (RSS). Interrogating the TF network within the pericyte/SM (C) and endothelial/endocardial (F) GRN identified the relationship of distinct regulons across pericyte/SM and endothelial/endocardial subpopulations (right). Nodes are colored by cell subpopulation. Node size is determined by their degree of centrality in the network, and edges represent TF-TF connections. LV - left ventricle, RV - right ventricle, LA- left atrium, RA - right atrium. ICM - ischemic cardiomyopathy, NICM - non-ischemic cardiomyopathy.

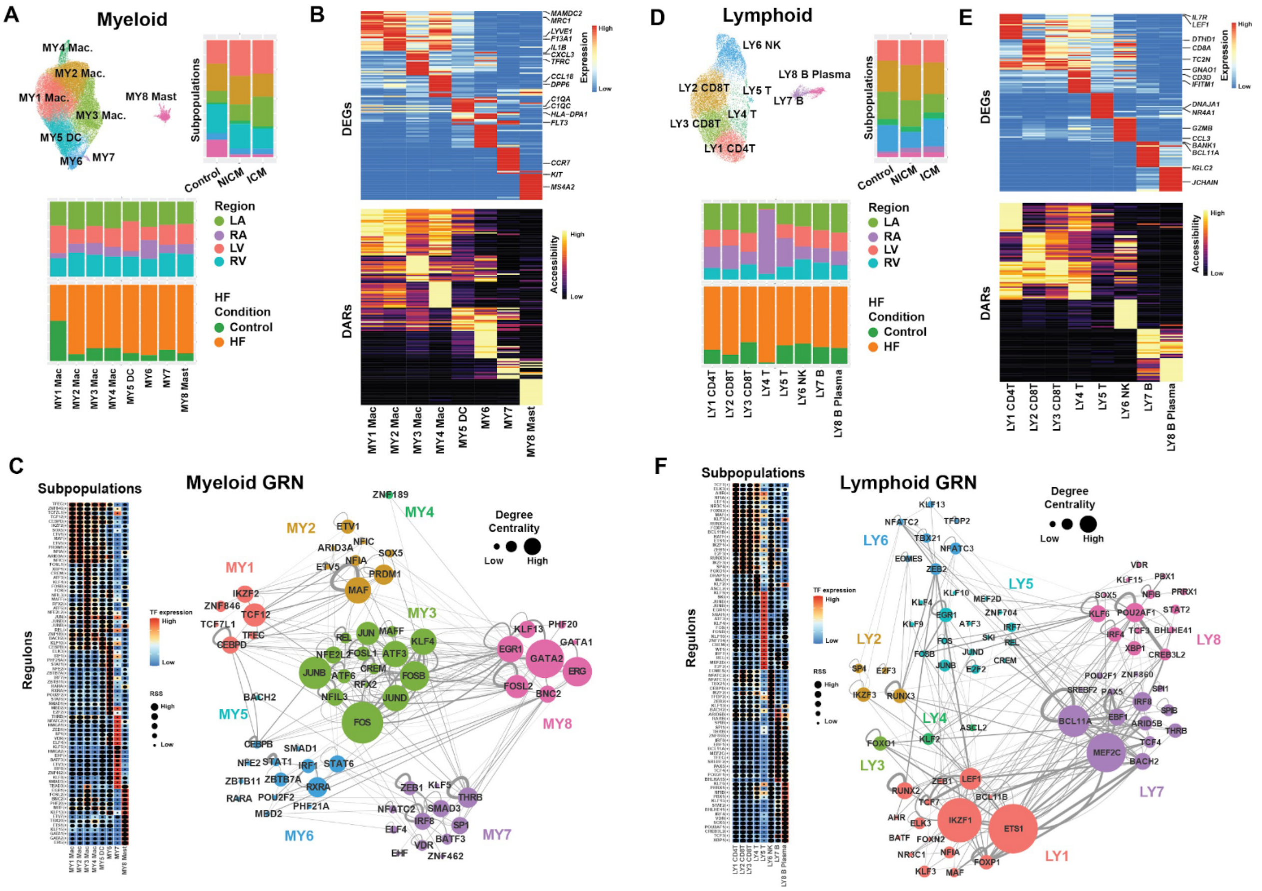


**Fig. S11. Gene regulatory network analysis of immune cells reveals genetic programs directing their cellular states.** (**A, D**) UMAP embeddings show myeloid/MY (MY, in A) and lymphoid/LY (LY, in D) cell subpopulations (top), respectively. Bar graphs to the right of UMAPs show the cell subpopulation composition across heart failure conditions. Bar graphs below UMAPs reveal the contribution of each cardiac chamber (top) and heart failure condition (non-HF versus HF) to specific cell subpopulations (bottom). (**B, E**) Each myeloid (B) and lymphoid (E) cell subpopulation displays distinct gene expression profiles (top heatmap) and chromatin accessibility pattern (bottom heatmap). (**C, F**) SCENIC+ predicted regulons are shown for myeloid (C) and lymphoid (F) cell subpopulations in respective heatmaps (left). Each box is colored by transcription factor expression, and the dot size indicates the regulon specificity score (RSS). Interrogating the TF network within the myeloid (C) and lymphoid (F) cell GRN identified the relationship of distinct regulons across myeloid and lymphoid subpopulations (right). Nodes are colored by cell subpopulation. Node size is determined by their degree of centrality in the network, and edges represent TF-TF connections. LV - left ventricle, RV - right ventricle, LA- left atrium, RA - right atrium. ICM - ischemic cardiomyopathy, NICM - non-ischemic cardiomyopathy.

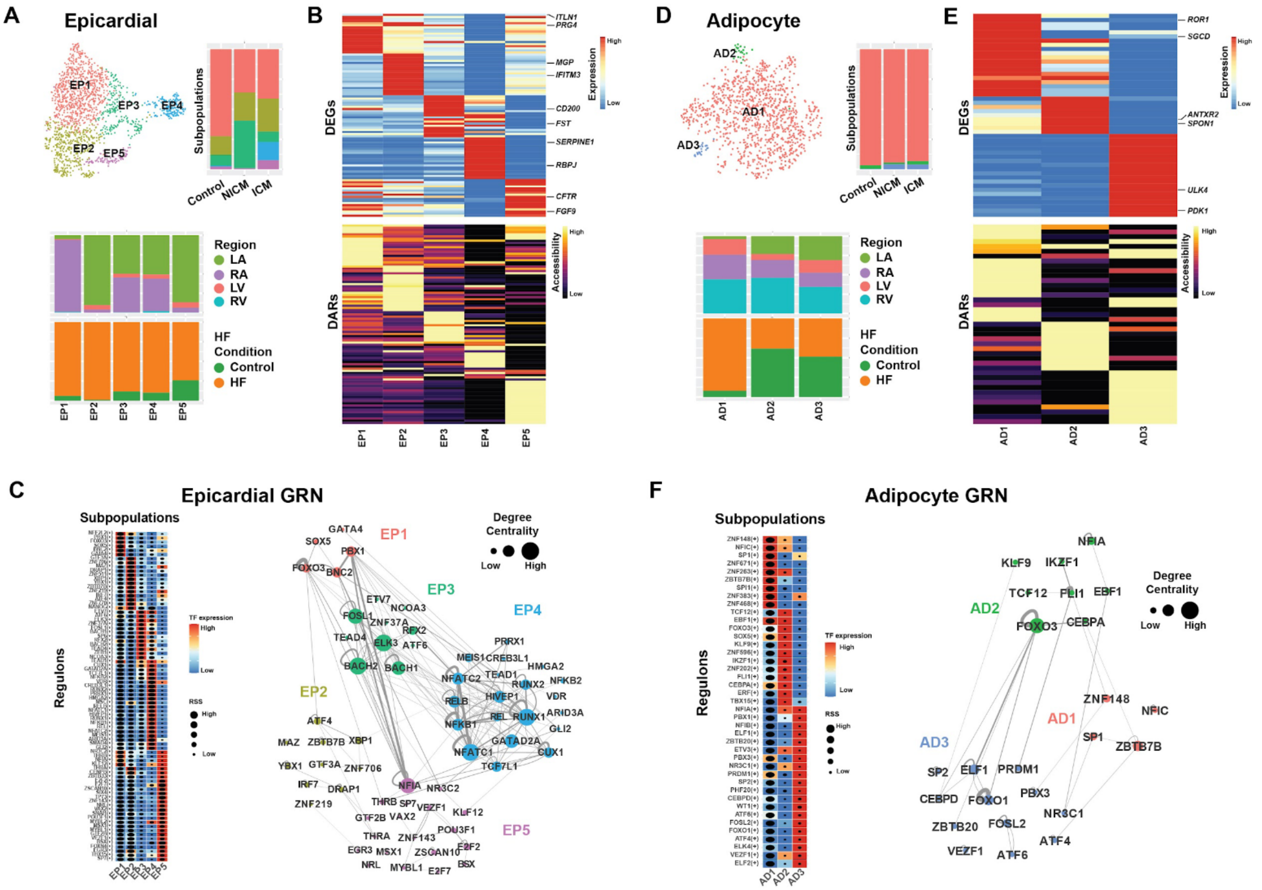


**Fig. S12. Gene regulatory network analysis of epicardial and adipocyte cells reveals genetic programs directing their cellular states.** (**A, D**) UMAP embeddings show epicardial/EP (EP, in A) and adipocyte/AD (AD, in D) cell subpopulations, respectively. Bar graphs to the right of UMAPs show the cell subpopulation composition across heart failure conditions. Bar graphs below UMAPs reveal the contribution of each cardiac chamber (top) and heart failure conditions (non-HF versus HF) to specific cell subpopulations (bottom). (**B, E**) Each epicardial (B) and adipocyte (E) cell subpopulation displays distinct gene expression profiles (top heatmap) and chromatin accessibility patterns (bottom heatmap). (**C, F**) SCENIC+ predicted regulons are shown in respective heatmaps for epicardial (C) and adipocyte (F) cell subpopulations (left). Box color indicates transcription factor expression, and dot size represents the regulon specificity score (RSS). Interrogating the TF network within the epicardial cell and adipocyte GRN identified the relationship of distinct regulons across epicardial and adipocyte subpopulations (right). Nodes are colored by cell subpopulation. Node size is determined by their degree of centrality in the network, and edges represent TF-TF connections. LV - left ventricle, RV - right ventricle, LA- left atrium, RA - right atrium. ICM - ischemic cardiomyopathy, NICM - non-ischemic cardiomyopathy.

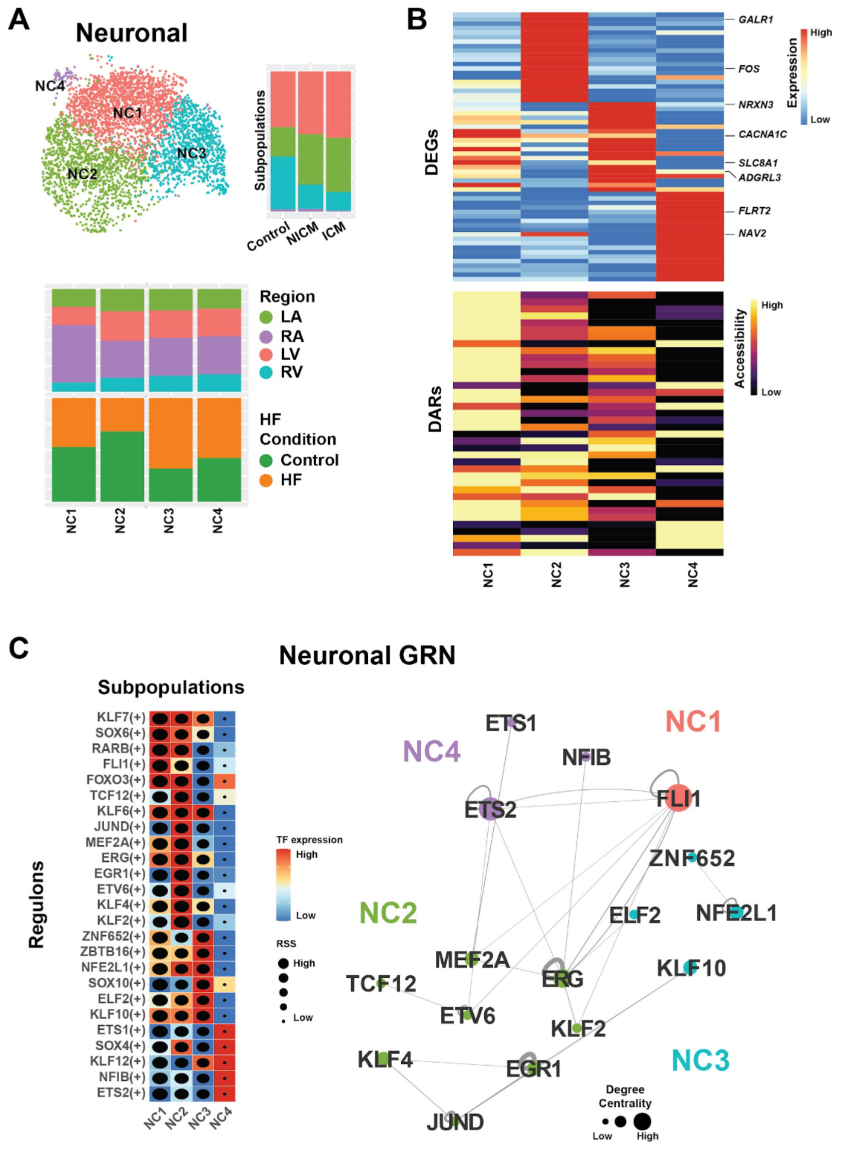


**Fig. S13. Gene regulatory network analysis of neuronal cells reveals genetic programs directing their cellular states.** (**A**) Neuronal cell (NC) subpopulations are displayed in UMAP embedding (top). Bar graphs to the right of UMAP show the cell subpopulation composition for each cardiac condition. Bar graphs below UMAP reveal the contribution of each cardiac chamber (top) and heart failure conditions (non-HF versus HF) to specific cell subpopulations (bottom). (**B**) Each neuronal cell subpopulation displays distinct gene expression profiles (top heatmap) and chromatin accessibility pattern (bottom heatmap). (**C**) Heatmap shows SCENIC+ predicted regulons for each neuronal cell subpopulation (left). Boxes are colored by transcription factor expression, and the dot size shows the regulon specificity score (RSS). Interrogating the TF network within the neuronal GRN identified the relationship of distinct regulons across neuronal states (right). Nodes are colored by cell subpopulation. Node size is determined by their degree of centrality in the network, and edges represent TF-TF connections. LV - left ventricle, RV - right ventricle, LA- left atrium, RA - right atrium. ICM - ischemic cardiomyopathy, NICM - non-ischemic cardiomyopathy.

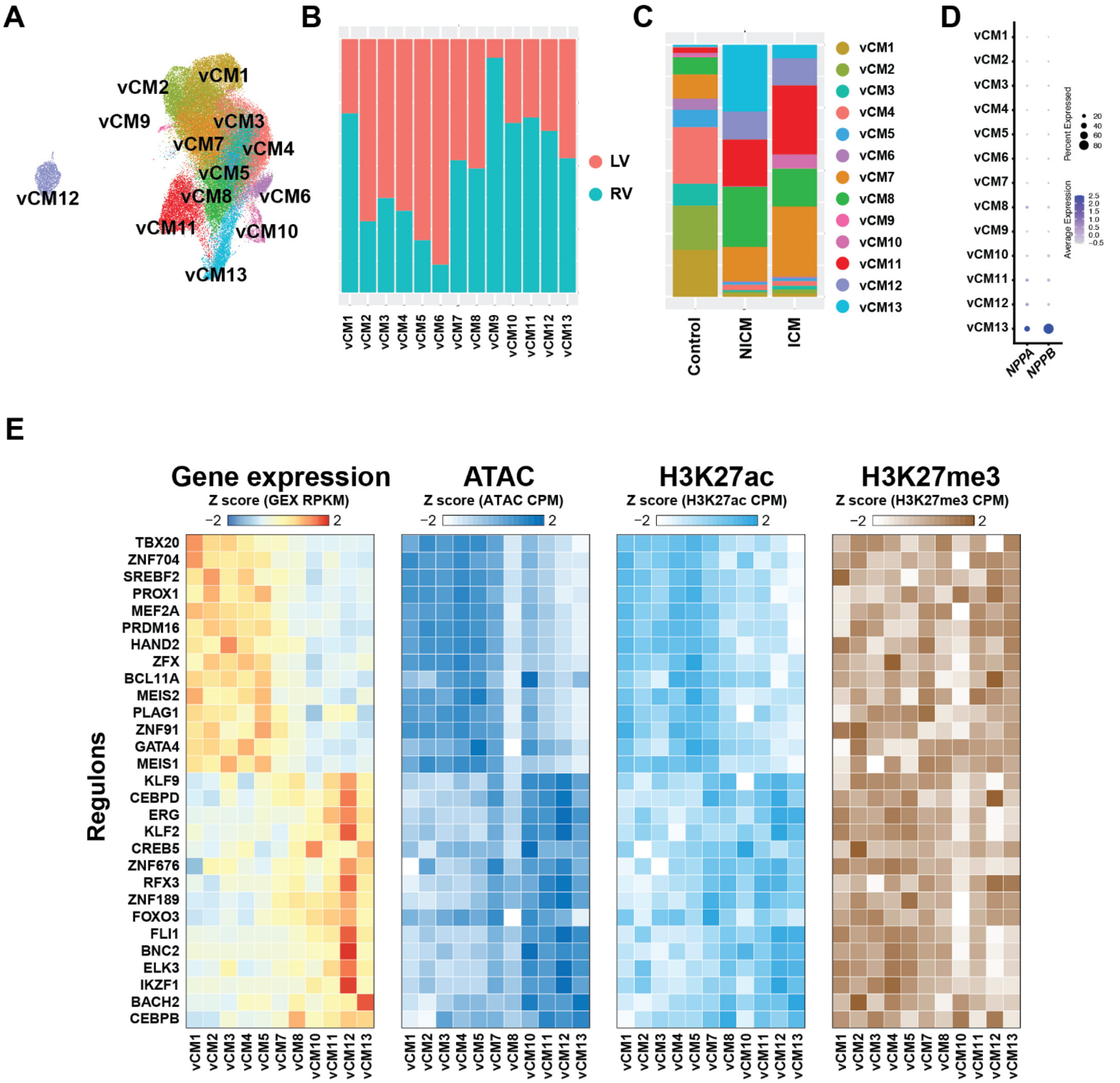


**Fig. S14. Analyzing ventricular cardiomyocyte states and their GRNs revealed genetic programs controlling their disease states.** (**A**) Subclustering of ventricular cardiomyocytes (vCMs) identified 13 distinct cell subpopulations, visualized by UMAP embedding. (**B**) Bar graph shows the contribution of cardiomyocytes from the cardiac chambers to each vCM subpopulation. (**C**) Bar graph shows the vCM subpopulation composition for each cardiac condition. (**D**) Dot plot displays the expression level of *NPPA* and *NPPB* across vCM subpopulations. (**E**) Heatmaps show the TF expression level as well as ATAC, H3K27ac, and H3K27me3 signal for example regulons across vCM subpopulations. LV - left ventricle, RV - right ventricle. ICM - ischemic cardiomyopathy, NICM - non-ischemic cardiomyopathy.

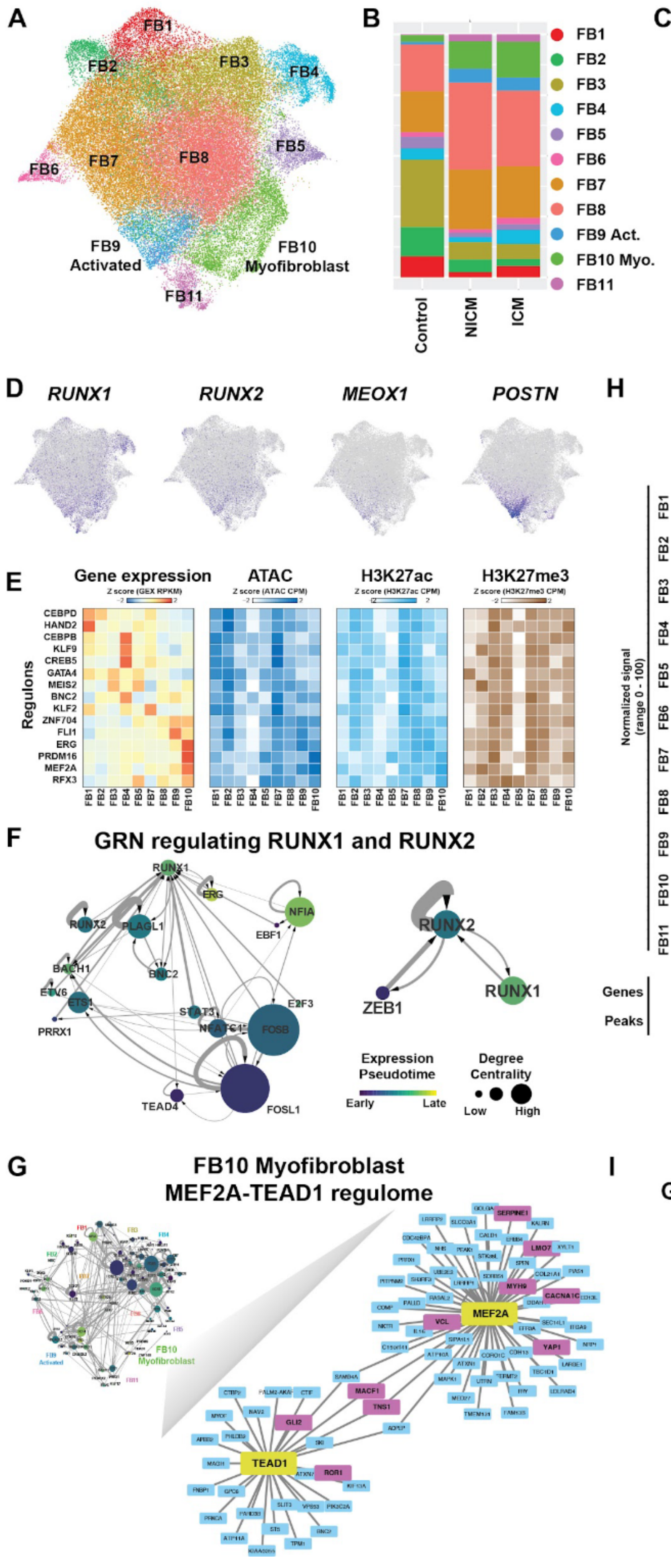
A
FB1
FB2
FB3
FB4
FB5
FB6
FB7
FB8
FB9 Activated
FB10 Myofibroblast
FB11
B
Control
NICM
ICM
FB9 Act.
FB10 Myo.
C
LA
RA
LV
RV
D
RUNX1
RUNX2
MEOX1
POSTN
E
Gene expression
ATAC
H3K27ac
H3K27me3
Regulons
CEBPD
HAND2
CEBPB
KLF9
CREB5
GATA4
MEIS2
BNC2
KLF2
ZNF704
FLI1
ERG
PRDM16
MEF2A
RFX3
F
GRN regulating RUNX1 and RUNX2
RUNX1
ERG
NFIA
EBF1
RUNX2
PLAGL1
BACH1
BNC2
ETV6
ETS1
PRRX1
STAT3
E2F3
FOSB
TEAD4
FOSL1
ZEB1
Expression Pseudotime
Early
Late
Degree Centrality
Low
High
G
FB10 Myofibroblast
MEF2A-TEAD1 regulome
MEF2A
TEAD1
SERPINE1
LMO7
MYH9
CACNA1C
YAP1
VCL
MACF1
TNS1
GLI2
ROR1
H
FB10 Myofibroblast
MEF2A-regulated YAP1
MEF2A
Normalized signal (range 0 - 100)
FB9 Act.
FB10 Myo.
YAP1
Genes
Peaks
RP11-732A21.3
102150000
102200000
chr11 position (bp)
I
GRN regulating MEF2A and TEAD1
MEF2A
TEAD1
KLF7
KLF12
MEF2C
NFIB
Expression Pseudotime
Early
Late
Degree Centrality
Low
High

**Fig. S15. Analyzing cardiac fibroblast states and their GRNs identified the genetic programs directing activated- and myo-fibroblasts.** (**A**) Fibroblast cells (FB) are subdivided into 11 distinct cell subpopulations, visualized by UMAP embedding. (**B**) Bar graph shows the cardiac fibroblast subpopulation composition for each cardiac condition. (**C**) Bar graph shows the contribution of fibroblasts from the cardiac chambers to each cardiac fibroblast subpopulation. (**D**) UMAPs reveal the expression of *RUNX1, RUNX2, MEOX1* and *POSTN* across cardiac fibroblast subpopulations. (**E**) Heatmaps show the TF expression level as well as ATAC, H3K27ac, and H3K27me3 signal of example regulons across fibroblast subpopulations. (**F**) Examining the gene regulatory networks for RUNX1 and RUNX2 identified potential transcription factors (TF) controlling their expression. Nodes are colored by pseudotime of expression, node size reflects degree centrality within the network, and edges represent TF-TF connections. **(G)** Network plot of the MEF2A and TEAD1 regulomes shows that their target genes include known gene markers of myofibroblasts (highlighted in magenta). (**H**) Representative genome browser track plot shows aggregated chromatin accessibility profiles across FB subpopulations for *YAP1* and a cCRE that is predicted by SCENIC+ to regulate *YAP1* by the MEF2A regulon. (**I**) GRN analysis for MEF2A and TEAD1 predicts upstream transcription factors regulating their expression. Nodes are colored by expression pseudotime, node size reflects degree centrality, and edges indicate TF-TF connections.

**A**

| Phenotype | Likely Causal Variant | Number of signals | Top Enriched Cell types | | Total Variants in Active Chromatin | Total signals in active chromatin | Total active chromatin peaks w/ variants |
|---|---|---|---|---|---|---|---|
| Non-ischemic cardiomyopathy (Strict) | 119 | 7 | aCM | vCM | 12 | 3 | 13 |
| Atrial fibrillation and flutter | 2929 | 107 | aCM | vCM | 161 | 64 | 237 |
| Ischaemic heart disease, wide definition | 2083 | 85 | Fibroblast | Pericyte | 184 | 54 | 326 |
| Myocardial infarction, strict | 1633 | 55 | Neuronal | Pericyte | 87 | 28 | 103 |
| Hypertension | 8658 | 278 | Pericyte | SM | 579 | 161 | 952 |
| Aortic aneurysm | 1048 | 22 | Fibroblast | Pericyte | 39 | 13 | 40 |
| Major coronary heart disease event | 2280 | 78 | Fibroblast | SM | 147 | 46 | 228 |

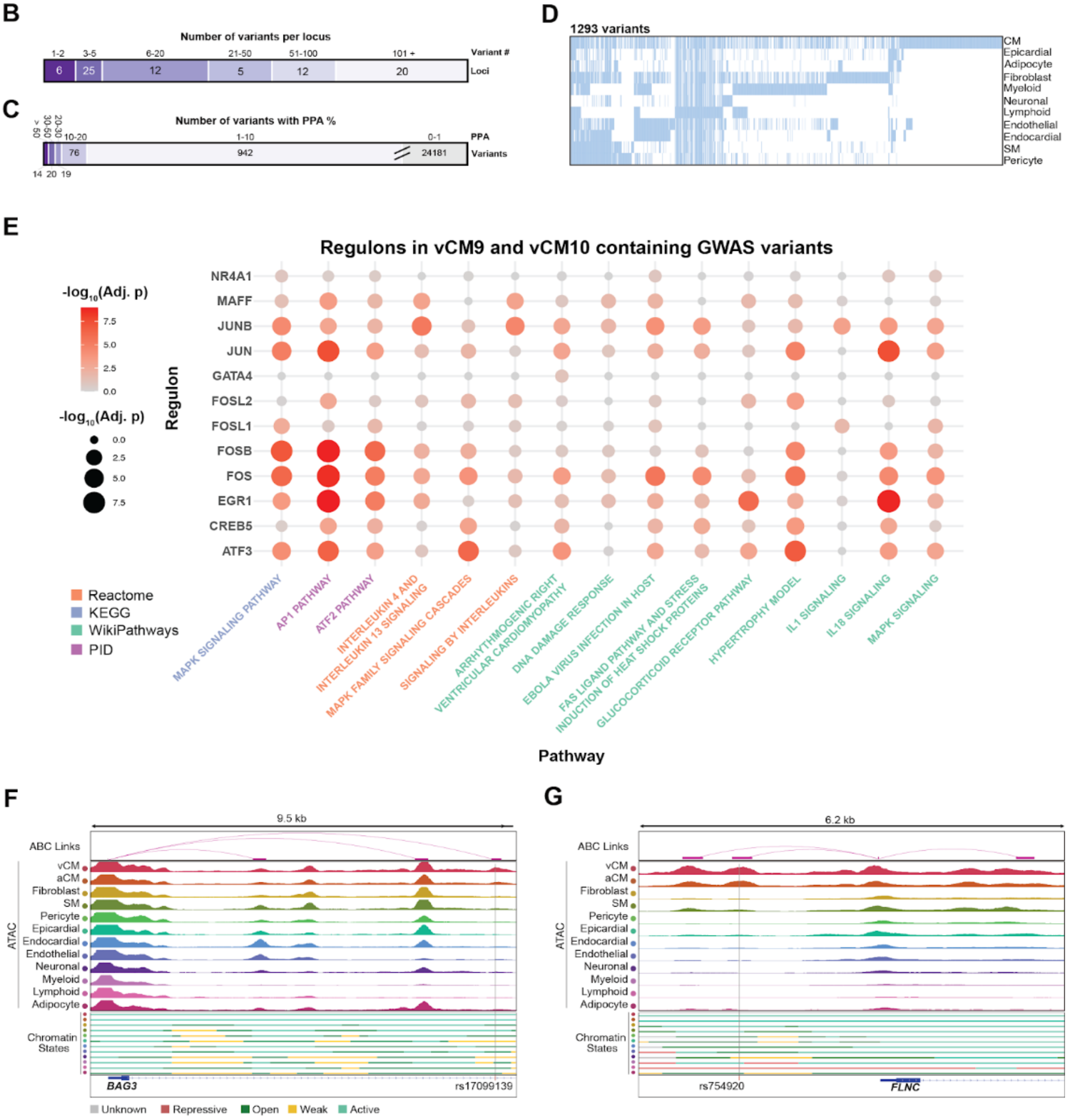

**Fig. S16. Fine-mapping of genetic loci associated with cardiovascular traits and heart failure.** **(A)** Table enumerates the number of fine-mapped variants, signals, and peaks harboring cardiovascular (CV)-associated variants (including heart failure/non-ischemic cardiomyopathy) for the top two cardiac cell types identified by LD Score Regression (LDSC). **(B)** Bar graph displays the number of fine-mapped variants per locus from the GWAS study by Zheng et al. (*7*), providing an overview of variant distribution across genetic loci. **(C)** Bar graph shows the number of fine-mapped variants stratified by different posterior probability (PPA) thresholds from the GWAS study by Zheng et al., reflecting the confidence levels in variant assignments. **(D)** Heatmap illustrates the cCRE peaks containing fine mapped genetic variants across cardiac cell types from Zheng et al. **(E)** Dot plot displays enriched pathways for target genes of the GRNs in vCM9 and vCM10 that contain GWAS variants. **(F)** ABC links reveal the interaction between a cCRE harboring the HF risk variant rs17099139 and the *BAG3* promoter (top). Aggregated chromatin accessibility profiles and chromatin states across all major cardiac cell types are shown below. **(G)** ABC links reveal the interaction between a cCRE harboring the HF risk variant rs754920 and the *FLNC* promoter (top). Aggregated chromatin accessibility profiles and chromatin state across all major cardiac cell types are shown below.

**<u>Tables S1 to S30</u>**

**S1: Clinical metadata for donors involved in study.**
**S2: Comparison of multiplex pooled vs single channel multiomic single-cell data.**
**S3: Metadata for nuclei integrated from all modalities.**
**S4: nNMF module and ATAC signal for cCREs across all major cell types.**
**S5: ChromHMM annotations of cCREs.**
**S6: Catalog of cell type-specific cCREs.**
**S7: Motif enrichment for cell type-specific cCREs.**
**S8: Gene Ontology analysis for cell type-specific cCREs.**
**S9: ABC distal cCRE-gene links across all cardiac cell types.**
**S10: GSEA for target genes in cell type-specific distal cCRE-gene links.**
**S11: Motif enrichment for cCREs in cell type-specific distal cCRE-gene links.**
**S12: Differentially expressed genes (DEGs) in HF.**
**S13: Differentially accessible regions (DARs) in HF.**
**S14: Differentially marked regions for H3K27ac and H3K27me3 in HF.**
**S15: Differential distal cCRE-gene links in HF.**
**S16: Cell type-specific HF-associated GRNs.**
**S17: Cardiac cell subpopulation GRNs.**
**S18: Cardiac cell subpopulation DARs.**
**S19: Cardiac cell subpopulation DEGs.**
**S20: Cardiac cell subpopulation GSEA.**
**S21: Cardiac cell subpopulation latent time.**
**S22: GWAS studies used for examining how cardiovascular-trait associated genetic variants may impact distinct cardiac cell types.**
**S23: LDSC results for all cell types stratified by cCREs chromatin states.**
**S24: FinnGen fine-mapped data intersection with cell type cCREs.**
**S25: Fine mapped genetic variants residing in cell type cCREs.**
**S26: Comparison between target genes identified in this study versus genes prioritized from Zheng et al. 2024.**
**S27: vCM GRNs containing fine mapped genetic variants.**
**S28: GSEA results for all regulons containing fine mapped genetic variants in intermediate states (vCM9 and vCM10).**
**S29: fgwas results reveal enrichment of DCM-associated variants in GRNs for intermediate states (vCM10).**
**S30: ChromBPnet results integrated with the GWAS summary stats.**

*** Supplementary tables are available from the authors upon request prior to publication.**
